# A Randomized Double-Blind Phase 2 Clinical Trial Treating Cervical Intraepithelial Neoplasia 2/3 with PepCan or *Candida*

**DOI:** 10.1101/2025.01.18.25320725

**Authors:** Mayumi Nakagawa, Teresa Evans, Milan Bimali, Hannah Coleman, Jasmine Crane, Nadia Darwish, Jennifer L. Faulkner, Amy Jones, Haley Kelly, Benjamin J. Lieblong, Yong-Chen Lu, Keanna Marsh, Intawat Nookaew, Charles M. Quick, David Ussery, Michael Robeson, Sumit Shah, Takeo Shibata, Heather R. Williams, William Greenfield

**Author notes:** CORRESPONDING AUTHOR, Mayumi Nakagawa, M.D., PhD., Department of Pathology, College of Medicine, University of Arkansas for Medical Sciences, 4301 West Markham Street Slot 502, Little Rock, AR 72205, U.S.A. **ABBREVIATIONS** CI, confidence interval; CIN, cervical intraepithelial neoplasia; DLT, dose-limiting toxicity; ELISPOT, enzyme-linked immunospot; HPV, human papillomavirus; LEEP, loop electrical excision procedure; LT-α, lymphotoxin-α; MDSC, myeloid-derived suppressor cells; PBMC, peripheral blood mononuclear cell; PDGF-ββ, platelet-derived growth factor-ββ; TCR, T cell receptor; Tregs, regulatory T cells; and TGF-β; tumor growth factor-β1.

## Abstract

**PURPOSE:** A non-surgical alternative for treating cervical intraepithelial neoplasia (CIN) 2/3 is an unmet need due to a risk of cervical incompetency.

**METHODS:** PepCan consists of four human papillomavirus (HPV) type 16 E6 peptides and a *Candida* skin testing reagent (adjuvant). In this randomized, double-blind Phase 2 study, women with biopsy-confirmed CIN2/3 were treated with PepCan or *Candida* at one to one ratio. Four intradermal injections were given every 3 weeks, with observation visits at 6 and 12 months post-vaccination. Quadrant biopsies were performed at the 12-month visit, and those whose lesions regressed to no CIN were considered to be complete responders. Regression rates of each treatment group were compared to that of a historical placebo group.

**RESULTS:** With the intention-to-treat analysis, PepCan (n=39) showed 30.8% efficacy (95% confidence interval [CI], 17 to 47.6; *p*=0.25) while *Candida* (n=42) demonstrated 47.6% efficacy (95% CI, 32 to 63.6; *p*<0.001). Likewise, with the per-protocol analysis, PepCan (n=24) showed 45.8% efficacy (95% CI, 25.6 to 67.2; *p*=0.08) and *Candida* (n=29) showed 62.1% efficacy (95% CI, 42.3 to 79.3; *p*<0.001). There was no difference between efficacy of PepCan and *Candida*. No dose-limiting toxicity was observed. HPV-specific T cell responses were elicited in both groups. Vaccine-induced HPV-specific CD4 and CD8 T cells were present in cervix regardless of histological response. Single-cell RNA-seq revealed increased expression of granzymes, CCR5, and EOMES in HPV-specific CD8-positive T cells of a histological responder, compared to non-responders. Six cytokines (CCL4, CCL5, interleukin-9, lymphotoxin-α, platelet-derived growth factor-ββ, tumor growth factor-β1) were significantly decreased in both *Candida* recipients and histological responders suggesting that *Candida* may possibly exert its anti-tumor effects through these systemic mediators.

**CONCLUSIONS:** *Candida* may be effective in inducing histological regression. PepCan and *Candida* treatments are safe. *Candida* should be evaluated in a Phase 3 trial as a potential new treatment for CIN2/3.

## BACKGROUND

Cervical intraepithelial neoplasia (CIN) 2/3 is a precursor of cervical cancer which is the fourth most common cancer among women globally, with annual global incidence of 604,127 cases and mortality of 341,831 cases.^1,2^ It is mostly caused by human papillomavirus (HPV), which is estimated to be responsible for 5.2% of cancer cases in the world.^3,4^

Standard surgical treatments of CIN2/3, such as loop electrical excision procedure (LEEP), are effective but may cause cervical incompetency resulting in increased preterm delivery rates,^5–8^ increased premature rupture of membrane,^9^ and lower birth weight.^9^ Therefore, current treatment guidelines recommend 1–2 years of close observation in women with CIN2, who are less than 25 years in age or who plan to have children. For CIN3, treatment is recommended.^6,10^ Non-surgical alternatives which would leave the cervix anatomically intact, such as therapeutic vaccines, are needed. As the World Health Organization has declared a call to action to accelerate elimination of cervical cancer through increasing prevention, detection, and treatment by 2030,^11^ the need for HPV therapeutic vaccines is great.

HPV transformation of squamous epithelium to a malignant phenotype is mediated by two early gene products, E6 and E7,^12^ and their expression is necessary for HPV 16 transformation of human cells.^13,14^ T cell responses to HPV 16 E6 protein have been associated with favorable clinical outcomes such as viral clearance^15^ and regression of cervical lesions.^16,17^ Therefore, the E6 protein is an attractive target for immunotherapy.

Traditionally, recall antigens, which typically include a panel of *Candida*, mumps, and *Trichophyton,* were used as controls to indicate intact cellular immunity when patients were being tested for *Tuberculosis* by placement of purified protein derivative (PPD) intradermally. A number of studies demonstrated that recall antigen injections can be used to treat common warts, and several have shown that treating warts with recall antigens is effective not only for injected warts but also distant, untreated warts.^18,19^ Furthermore, *in vitro* studies have demonstrated that *Candida* can induce interleukin-12 expression and T cell proliferation.^20,21^ Therefore, we utilized *Candida* for the first time as a vaccine adjuvant in a Phase 1 clinical trial (NCT01653249) of our group’s HPV therapeutic vaccine, PepCan, which is comprised of four synthetic peptides covering HPV 16 E6 protein and *Candida*. PepCan’s safety was demonstrated, and the lowest dose was most efficacious.^22,23^ Furthermore, a systemic increase of Th1 cells and higher clearance rates of non-HPV 16 types in comparison to HPV 16 suggested that the adjuvant, *Candida*, exerted notable immune enhancement.^22,23^ This provided the rationale for testing *Candida* by itself along with PepCan in this Phase 2 clinical trial.

Therefore, patients were randomized in a double-blind manner to PepCan or *Candida* (NCT02481414). The hypothesis was that PepCan would induce HPV-specific immune responses resulting in high-grade squamous intraepithelial regression when the T cells reach the disease sites. While *Candida* is expected to have some immune enhancement effects, it was expected to be less effective than PepCan. Here, we report the unexpected outcome of *Candida* alone appearing to be effective in regressing CIN2/3 in comparison to a historical placebo group.^2^

## METHODS

### PATIENTS

Patients (n=99) were screened between November 2015 and June 2021 at the University of Arkansas for Medical Sciences (UAMS) Medical Center, and those with biopsy-proven CIN2/3 (n=81) were eligible and were randomly assigned to the PepCan or *Candida* arm (Figure 1).

**Figure 1.**
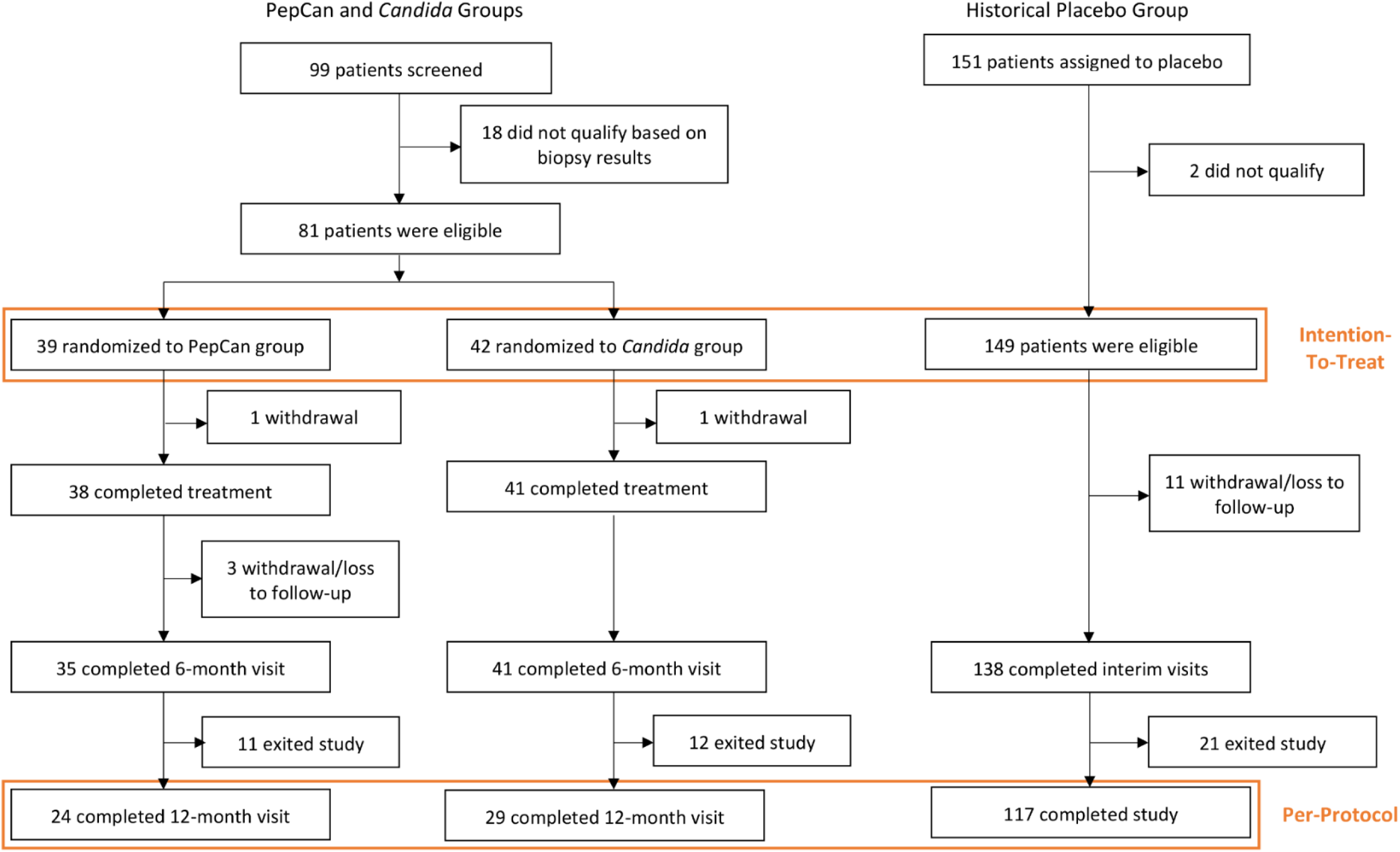
CONSORT diagram.

Exclusion criteria included immunosuppression, pregnancy, breast feeding, allergy to *Candida*, severe asthma, and history of invasive squamous cell carcinoma of the cervix. A full description of eligibility criteria are in the trial protocol submitted with this manuscript.

### VACCINE COMPOSITION AND DELIVERY

The vaccine consisted of four current good manufacturing production-grade synthetic peptides covering the HPV 16 E6 protein [amino acid (aa)1-45, 46-80, 81-115, and 116-158] (CPC Scientific, San Jose, CA; Curia, Albany, NY).^21^ Lyophilized peptides (50 μg per peptide) were reconstituted with sterile water and were mixed in a syringe with 0.3 mL of *Candida* (Candin®, Nielsen Biosciences, San Diego, CA). This is an extract of *Candida*, and is not heat-killed nor fixed. The PepCan mixture was administered intradermally in any limb, but most often in the anterior forearm. In the *Candida* arm, 0.3 mL of *Candida* was also used.

### TRIAL DESIGN AND END POINTS

This clinical trial (UAMS IRB 202790) was a randomized, double-blind, single-center, Phase 2 study in which patients were assigned to PepCan or *Candida* at a one-to-one ratio using a computer-generated randomization scheme created by the study statistician. The patients and all study personnel were blinded to which vaccine the patients received, except for the research pharmacists. Four injections were given 3 weeks apart, and the patients returned for observational visits 6 and 12 months after the last injection (Supplementary Figure 1). A liquid-based cytology sample (Thin-Prep, Hologic, Marlborough, MA) was collected for HPV-DNA testing for 37 individual HPV types (Linear Array HPV Genotyping Test, Roche Molecular Diagnostics, Pleasanton, CA) at the screening visit, 6-month visit, and 12-month visit. HPV 16 viral load was also determined^22,24^ for subjects who were HPV 16-positive at the screening visit.

The primary endpoint was histological regression, which was assessed using four quadrant biopsies collected at the 12-month visit. At least one biopsy was taken from each quadrant with visible lesion under acetic acid application. Random biopsies were taken from quadrants without visible lesions. Endocervical curettage was performed per standard of care. Biopsies were examined by two pathologists who were blinded to each other’s diagnosis. For patients who exited the study after the 6-month visit, the results of the biopsy (performed if colposcopy showed possible progression with acetic acid application) and/or LEEP were used to assess histological responses. The secondary endpoint was safety, which was assessed from the time informed consent was obtained until the 6- or 12-month visit according to the National Cancer Institute Common Terminology Criteria for Adverse Events, Version 4.03. A patient with no dysplasia was considered a complete responder, a patient with CIN1 was considered a partial responder, and a patient with CIN2/3 was considered a non-responder. Only patients with no CIN were considered histological responders with a stringent criterion, and those with no CIN or CIN1 were considered to be responders with a lenient criterion.

Additional endpoints involved examination of peripheral immune responses by examining T cell activity to 10 regions (aa1-25, 16-40, 31-55, 46-70, 61-85, 76-100, 91-115, 106-130, 121-145, and 136-158) of the HPV 16 E6 protein using an interferon-γ enzyme-linked immunospot (ELISPOT) assay (n=71). Epitope spreading to HPV 16 E7 protein, HPV 18 E6 protein, HPV 35 E6 protein, or HPV 52 E6 protein was also examined (n=32) for patients testing positive for respective HPV type at entry. In patients who completed the 12-month visit and demonstrated sustained T cell response to the HPV 16 E6 protein, bulk T cell receptor (TCR) β deep sequencing of peripheral blood mononuclear cells (PBMCs) and liquid-based cervical cytology samples was performed (n=15). If post-vaccination responses were statistically significant, single-cell RNA-seq and TCR sequencing were also performed (n=5) to identify vaccine-induced HPV-specific T cells.

Immune profiling (n=76) measuring Th1 cells, Th2 cells, regulatory T cells (Tregs),^22,23^ and myeloid-derived suppressor cells (MDSCs)^25,26^ was also performed using fluorescent-activated cell sorter (FACS) with blood samples drawn at visit 1, visit 3, 6-month visit, and12-month visit. To identify potential biomarkers predictive of vaccine response and to examine the effects of vaccination, patient samples from those who completed the 6-month visit (n=76) were examined for 51 plasma cytokines, and 1,454 plasma metabolomics. As our pilot data demonstrated that about a third of cytokines levels were significantly different when the blood samples were processed at 2 hours versus 1 hour,^22^ we used plasma samples that were processed within 1 hour of blood draw. If the blood was processed after 1 hour but within 2 hours, data for the labile cytokines were not used from one plasma sample. None of the blood samples were processed beyond 2 hours. The detailed methods are described in the Supplementary Appendix.

### HLA TYPING

Using DNA extracted from PBMC, low-resolution typing for HLA class I A, B, and C and class II DRB1, DQB1, and DPB1 was performed (n=80) with MicroSSP Generic DNA Typing Trays according to the manufacturer’s instructions (One Lambda, Los Angeles, CA). See Supplementary Appendix for more details.

### CERVICAL MICROBIOME

As the phase 1 results suggested potential negative roles of phyla *Caldithrix* and *Nitrospirae* in vaccine response,^27^ cervical microbiome was examined using the DNA extracted from liquid-based cytology samples through amplification and sequencing of the 16S rRNA gene (see details in the Supplementary Appendix).

### TRIAL OVERSIGHT

The study was approved by the Institutional Review Board and was supported by the National Cancer Institute. A written informed consent was obtained from each participant. No commercial support was provided for this study, and no person who is not an author contributed to the manuscript. All authors participated in data acquisition, had access to data, wrote and/or edited the manuscript, and vouched for the accuracy and completeness of the data.

### STATISTICAL ANALYSIS

It was estimated that a sample size of 35 would provide 93% power to detect a difference in response rate of 31% (60% in PepCan group versus a 29% in historical placebo group).^2^ This historical placebo group was selected because patients with CIN2/3 were treated with a placebo in one arm and were observed for regression to no CIN during the same length of time (i.e., 15 months from the treatment initiation).^2^ The sample size was estimated based on exact test statistic and a two-sided type I error of 5%. The goal was to start vaccinating 40 patients in each group to account for attrition. The study was not powered to show differences between the *Candida* and placebo groups since the *Candida* group was expected to be less efficacious than PepCan. Likewise, we did not expect to see a significant difference between the PepCan and *Candida* groups. Each group was compared to the historical placebo using the exact binomial test (two-sided). The PepCan and *Candida* groups were compared using Pearson’s χ2-tests (two-sided). The Clopper-Pearson method was used to determine 95% confidence intervals (CIs). A two-sided *p*-value of less than 0.05 was used to determine statistical significance, and corrections for multiple comparisons were made where appropriate. A more detailed descriptions of statistical methods are described in the Statistical Analysis Plan.

## RESULTS

### PATIENTS AND TREATMENT

Thirty-nine patients were randomly assigned to the PepCan arm and 42 patients were assigned to the *Candida* arm (the intention-to-treat population). Their baseline characteristics were well balanced (Table 1 and Figure 1). Limited demographic information for age, ethnicity, and current oral contraceptive use was available for the historical placebo group.^2^ The placebo group had a significantly higher percentage of patients younger than 25 years old (*p*<0.001 with PepCan and *p*=0.005 with *Candida*), and significantly more current use of oral contraceptives (*p*<0.001 with PepCan and *p*=0.04 with *Candida*).

**Table 1.** Baseline Characteristics of the Patients^.

| Characteristic/Group | PepCan Group<br>N=39 | *Candida* Group<br>N=42 | Total<br>N=81 |
| --- | --- | --- | --- |
| Age, years |  |  |  |
| Mean ± SD | 31.3 ± 6.4 | 31.4 ± 6.1 | 31.3 ± 6.2 |
| Range | 21.5 – 45.1 | 22.4 – 43.6 | 21.5 – 45.1 |
| <25 years – no. (%) | 6 (15.4) | 9 (21.4) | 15 (18.5) |
| ≥25 years – no. (%) | 33 (84.6) | 33 (78.6) | 66 (81.5) |
| Race – no. (%) |  |  |  |
| Caucasian | 34 (87.2) | 37 (88.1) | 71 (87.6) |
| African American | 4 (10.3) | 4 (9.5) | 8 (9.9) |
| Other | 1 (2.6) | 1 (2.4) | 2 (2.5) |
| Ethnicity – no. (%) |  |  |  |
| Hispanic | 11 (28.2) | 12 (28.6) | 23 (28.4) |
| Non-Hispanic | 28 (71.8) | 30 (71.4) | 58 (71.6) |
| >CIN2 – no. (%) | 29 (74.4) | 26 (61.9) | 55 (67.9) |
| =CIN2 – no. (%) | 10 (25.6) | 16 (38.1) | 26 (32.1) |
| No. cervical quadrants with visible lesions |  |  |  |
| Mean ± SD | 1.89 ± 1.06 | 2.05 ± 1.02 | 1.97 ± 1.04 |
| >2 quadrants – no. (%) | 9 (23.7) | 11 (26.8) | 20 (25.3) |
| ≤2 quadrants – no. (%) | 29 (76.3) | 30 (73.2) | 59 (74.7) |
| Body mass index (kg/m<sup>2</sup>) |  |  |  |
| Mean ± SD | 27.3 ± 6.2 | 29.1 ± 7.3 | 28.2 ± 6.8 |
| ≥25 kg/m<sup>2</sup> – no. (%) | 21 (53.8) | 26 (63.4) | 47 (58.8) |
| ≥30 kg/m<sup>2</sup> – no. (%) | 11 (28.2) | 19 (46.3) | 30 (37.5) |
| Albumin < 3.5 g/dL – no. (%) | 1 (2.6) | 3 (7.3) | 4 (5.0) |
| Total protein < 6.4 g/dL – no. (%) | 3 (7.7) | 5 (12.2) | 8 (10.0) |
| History of HPV prophylactic vaccine – no. (%) | 7 (17.9) | 7 (16.7) | 14 (17.3) |
| Any college – no. (%) | 20 (52.6) | 20 (47.6) | 40 (50.0) |
| Any children – no. (%) | 27 (69.2) | 27 (64.3) | 54 (66.7) |
| ≥ 5 sexual partners – no. (%) | 19 (48.7) | 30 (73.2) | 49 (61.3) |
| Ever smoked – no. (%) | 18 (46.1) | 15 (35.7) | 33 (40.7) |
| Currently smoke – no. (%) | 8 (20.5) | 9 (21.4) | 17 (21.0) |
| Ever used oral contraceptives – no. (%) | 27 (69.2) | 32 (76.2) | 59 (72.8) |
| Currently use oral contraceptives – no. (%) | 6 (15.4) | 15 (35.7) | 21 (25.9) |

^Data for the intention-to-treat population are shown which included patients who qualified for vaccination and were randomized.

### EFFICACY

In comparison to the historical placebo group, the *Candida* group, but not the PepCan group, demonstrated significantly higher histological regression rates in the intention-to-treat (47.6%, 95% CI, 32 to 63.6; *p*<0.001) and per-protocol (62.1%, 95% CI, 42.3 to 79.3; *p*<0.001) populations. There were no significant differences in histological response rates of the PepCan and *Candida* groups in both analyses. Therefore, both groups combined and separately were analyzed for stringent histological responses in the intention-to-treat (Figure 2A) and per-protocol populations (Figure 2B) as well as in subgroups characterized by age, initial diagnosis, and initial number of cervical quadrants involved. Analogous analyses were performed using the lenient criterion for histological response (Figure S2A and S2B). No significant differences were found between the subgroups examined except with the comparisons of initial diagnoses between “CIN2” and “>CIN2”. In the intention-to-treat population for both treatment groups combined, “CIN2” had a response rate of 61.5% while “>CIN2” had 29.1% (relative risk [RR], 2.12; 95% CI, 1.26 to 3.53; *p*=0.01) using the stringent criterion. With the lenient criterion, the response rate for “CIN2” was 73.1% and that of “>CIN2” was 36.4% (RR, 2.01; 95% CI, 1.31 to 3.08; *p*=0.004) for both groups, the response rate for “CIN2” was 87.5% and that of “>CIN2” was 38.5% (RR, 2.28, 95% CI, 1.39 to 3.99; *p*=0.003) for the *Candida* group. No cases of invasive cervical cancer were detected within the 12-month follow-up time.

**Figure 2.**
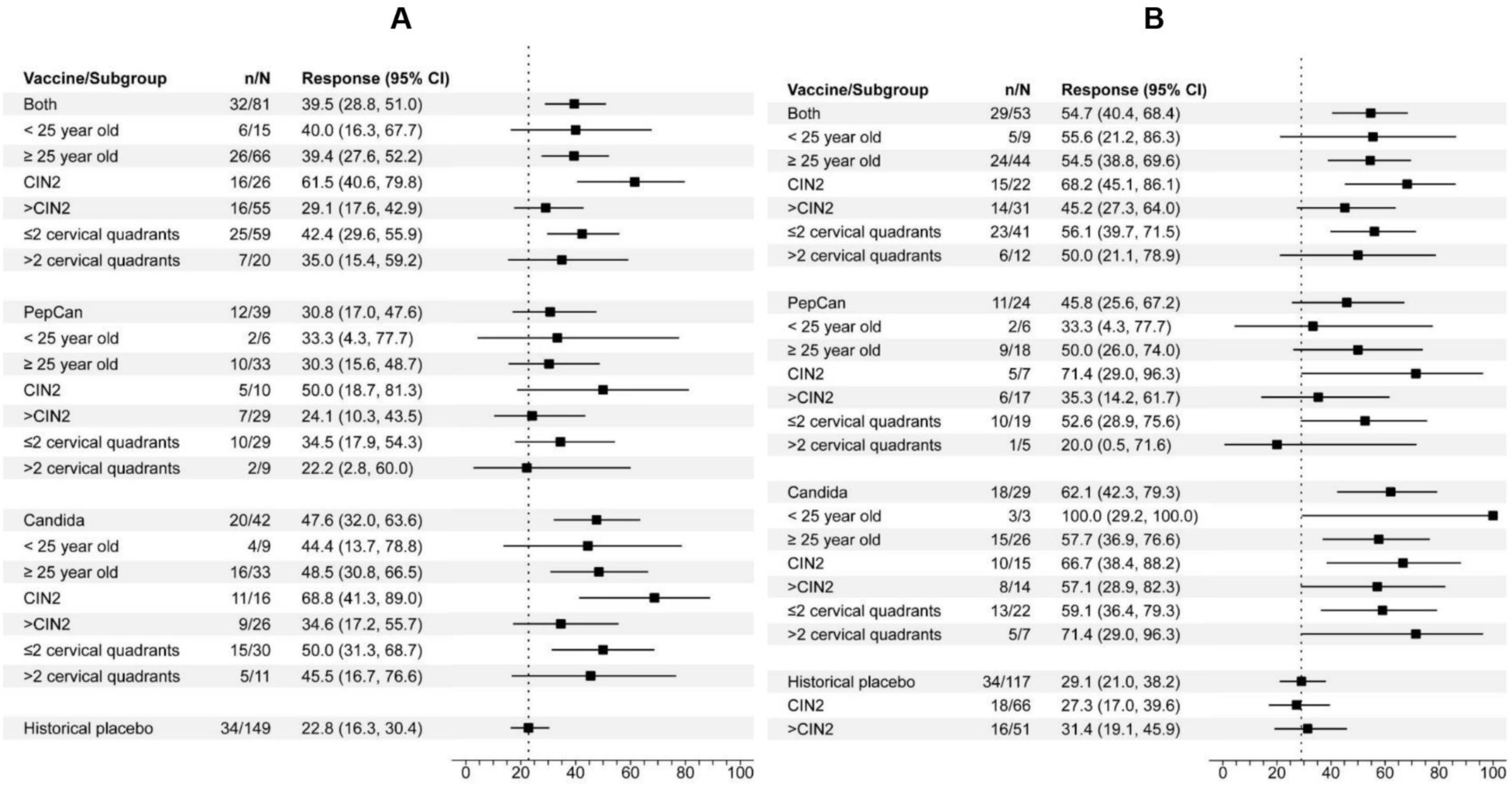
Histological responses and subgroup comparisons. Panel A shows the analyses of the intention-to-treat populations and panel B shows the analyses of the per-protocol analyses. Both panels used the stringent criterion in which only the complete responders (regression to no CIN) were considered to have had responses. The Clopper-Pearson method was used to determine the 95% confidence intervals (CIs).

### SAFETY

Treatment-related commonly occurring (with 5% or more of injections) adverse events (AEs) are summarized in Table 2, and all AEs, regardless of relatedness to treatments are summarized in Supplementary Table 1. No dose-limiting toxicity (DLT) was reported, and the most frequent AE was immediate injection site reaction for both treatment groups. Delayed injection site reaction (*p*=0.02) and myalgia (*p*<0.001) were significantly more frequently observed in the PepCan group in comparison to the *Candida* group (Table 2). No patients discontinued due to AEs.

**TABLE 2.** A comparison of injection-related adverse events occurring in ≥ 5% of injections with PepCan or *Candida*.

|  | PepCan n = 152 |  |  | *Candida* n = 164 |  |  | P-value* |
| --- | --- | --- | --- | --- | --- | --- | --- |
|  | Grade 1<br>n (%) | Grade 2<br>n (%) | All Grades<br>n (%) | Grade 1<br>n (%) | Grade 2<br>n (%) | All Grades<br>n (%) |  |
| **Fever** | 9 (5.9%) | 1 (0.7%) | 10 (6.6%) | 3 (1.8%) | 0 (0.0%) | 3 (1.8%) | 0.06 |
| **Headache** | 13 (8.6%) | 4 (2.6%) | 17 (11.2%) | 15 (9.2%) | 3 (1.8%) | 18 (11.0%) | 0.91 |
| **Injection site reaction, < 24 h** | 63 (41.5%) | 25 (16.5%) | 88 (57.9%) | 58 (35.4%) | 31 (18.9%) | 89 (54.3%) | 0.54 |
| **Injection site reaction, $\geq 24$ h** | 23 (15.1%) | 31 (20.4%) | 54 (35.5%) | 39 (23.8%) | 18 (11.0%) | 57 (34.8%) | **0.02** |
| **Myalgia** | 35 (23.0%) | 2 (1.3%) | 37 (24.3%) | 9 (5.5%) | 1 (0.6%) | 10 (6.1%) | **<0.001** |
| **Nausea** | 19 (12.5%) | 0 (0.0%) | 19 (12.5%) | 18 (11.0%) | 0 (0.0%) | 18 (11.0%) | 0.73 |

\*Fisher's exact test (2-sided)
n, number of injections
The highest grade is reported for each adverse event for a given injection

### IMMUNE RESPONSES

Following vaccination, new CD3 T cell responses to at least one region of the HPV 16 E6 protein were detected in 18 of 33 patients (54.5%) in the PepCan group and in 17 of 38 (44.7%) patients in the *Candida* group. Therefore, exogenous HPV antigens were not necessary to induce anti-HPV T cell responses. Anti-HPV 16 E6 T cell responses were detectable prior to vaccination in 7 of 33 patients (21.2%) in the PepCan group and in 13 of 38 patients (34.2%) in the *Candida* group. The histological responders demonstrated new E6 responses in 14 of 31 (45.2%) and non-responders in 21 of 40 (52.5%). Pre-existing E6 responses were present in 12 of 31 (38.7%) of responders and 8 of 40 (20%) of non-responders. Representative assay results are shown in Figure S3. Epitope spreading was demonstrated in 7 of 18 patients (38.9%) in the PepCan group, in 8 of 14 (57.1%) patients in the *Candida* group, 6 of 10 (60%) histological responders, and 9 of 22 (40.9%) histological non-responders (Figure 3A). None of these comparisons reached statistical significance.

**Figure 3.**
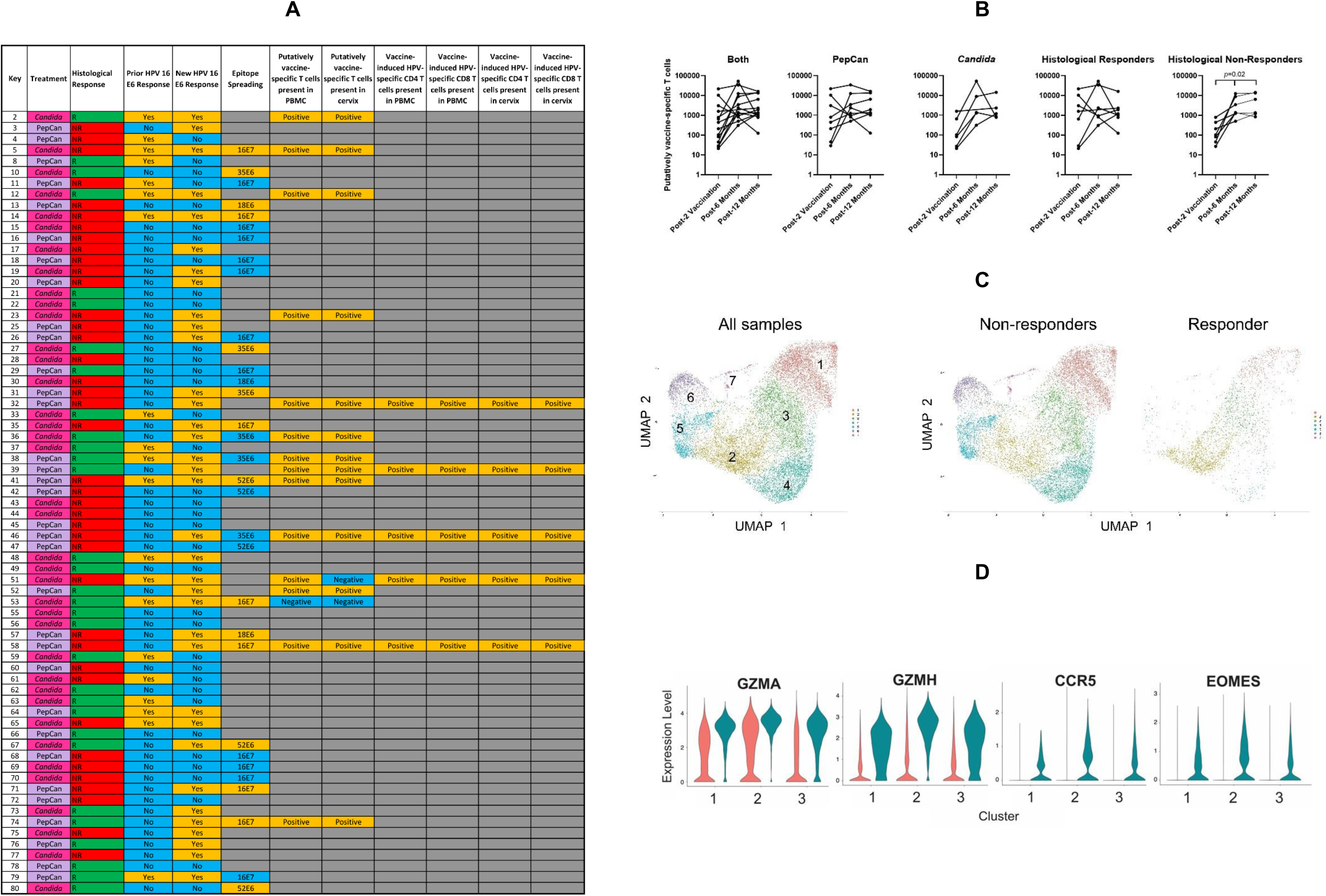
Immune responses assessed using IFN-γ ELISPOT assay, bulk T cell receptor β deep sequencing, and single-cell sequencing. A. The data from IFN-γ ELISPOT assay of 71 of 76 patients whose results were technically adequate are summarized along with the results of epitope spreading (32 of 33 patients with technically adequate results). Putatively vaccine-specific T cells were determined by performing bulk TCR β deep sequencing using PBMC samples of patients with new HPV 16 E6 ELISPOT responses in at least 2 of 3 post-vaccination samples using a beta-binomial model^55^ (n=15). Vaccine-induced HPV-specific CD8-positive and CD8-negative (i.e., CD4) T cells were selected on the basis of interferon-γ production upon stimulation with peptides from the HPV region(s) identified by ELISPOT assays for persistent post-vaccination responses (i.e., present in at least 2 of 3 post-vaccination visits) which were statistically significant (two-sided paired *t*-test). These regions were HPV 16 E6 aa136-aa158 for Patient 32, E6 aa121-aa145 for Patient 39, E6 aa16-aa40 and aa46-aa70 for Patient 46, E6 aa121-aa145 and aa136-aa158 for Patient 51, and E646-aa70 and E7aa16-aa40 for Patient 58. The presence of these vaccine-induced HPV-specific T cells in PBMC and cervix (n=5) were determined by examining CDR3 β amino acid sequences of bulk TCR β deep sequencing results. B. In histological non-responders (stringent), significant increase in the number of putatively vaccine-specific T cells (*p*=0.02, Friedman test) were demonstrated suggesting that the lack of histological response is not due to poor systemic T cell responses. C. CD8-positive interferon-γ producing T cells from 5 patients were enriched by FACS and subjected to single-cell RNA-seq analysis. Seven clusters were identified by clustering analysis, as shown in the UMAP plots. T cells isolated from a responder mostly were found in cluster 1, 2 and 3. D. The expression levels of granzymes (GZMA and GZMH), CCR5 and EOMES in interferon-γ producing CD8 T cells in cluster 1, 2 and 3 were elevated in a responder (teal), compared to non-responders (orange; *p*<1×10^-90^ for all comparisons, Seurat v4.4.0).

TCR β deep sequencing analyses of PBMC samples in patients with sustained new HPV 16 E6 responses demonstrated by the ELISPOT assay (n=15) did not show any difference between the PepCan and *Candida* groups when the number of putatively vaccine-specific T cells were compared. However, there was a significant increase of such T cells among histological non-responders (*p*=0.02), but not in histological responders suggesting that the lack of histological response is not due to poor peripheral response (Figure 3B). Analogous analyses using the fraction of putatively vaccine-specific T cells yielded the same results (data not shown). The putatively vaccine specific T cells in the cervix were detected in 7 of 8 (87.5%) responders and 6 of 7 (85.7%) non-responders. Using single-cell TCR-seq, the presence of vaccine-induced HPV-specific CD4 and CD8 T cells were confirmed in 5 of 5 patients in peripheral blood and in the cervix (Figure 3A). The percentages of vaccine-induced HPV-specific T cells in the cervix varied from 1.7% to 14.1% (Table S6). As only 1 of 5 patients was a histological responder, the mere presence of HPV-specific T cells is not the determining factor in histological response. Single-cell RNA-seq revealed increased expression of granzymes, CCR5, and EOMES in a histological responder compared to non-responders (Figure 3C/D), suggesting that the abundance of terminally exhausted T cells might be associated with a histological response (*p*<1×10^-90^ for all comparisons).^28,29^

When correlations between pre-vaccination individual plasma cytokines (n=51) and metabolites (n=1,454) were examined in histological responders and non-responders (stringent) in hopes of identifying potential biomarkers of vaccine response, none were identified (see Tables S2 and S3 in Supplementary Appendix). When changes over time were examined in order to gain insight into how the vaccines work, significant differences from the baseline were observed for 11 of 51 plasma cytokines examined (Figure 4A). None were significant among the 1,454 metabolites examined (data not shown). Most of the significant cytokine changes were decreases from the baseline. Only a few changes were noted in the PepCan group and histological non-responders while the *Candida* group and histological responders both demonstrated decreased levels of 7 and 11 cytokines respectively (Figure 4A). The *Candida* group and the responders shared decreases of CCL4 [MIP-1β](*p_adj_*=0.01 at 6 months, *p_adj_*=0.02 at 12 months for *Candida*, and p*_adj_*=0.005 at 6 months, *p_ad_*_j_<0.001 at 12 months for responders) CCL5 [RANTES](*p_adj_*<0.001, *p_adj_*=0.005, p*_adj_*=0.002, and *p_ad_*_j_<0.001 respectively), IL-9 (*p_adj_*=0.01, *p_adj_*=NS, p*_adj_*=0.003, and *p_ad_*_j_=0.002 respectively), lymphotoxin-α (LT-α) [TNF-β] (*p_adj_*=0.03, *p_adj_*=NS, p*_adj_*=0.008, and *p_ad_*_j_=0.003 respectively), platelet-derived growth factor-ββ (PDGF-ββ)(*p_adj_*=0.02, *p_adj_*=0.03, p*_adj_*=0.009, and *p_ad_*_j_=0.002 respectively),, and tumor growth factor-β1 (TGF-β1)(*p_adj_*=0.04, *p_adj_*=0.04, p*_adj_*=0.02, and *p_ad_*_j_=0.01 respectively), levels suggesting that histological regression of CIN2/3 may be mediated through *Candida* modulating these systemic immune mediators (Figure 4B). The most pronounced decreases were observed for CCL5.

**Figure 4.**
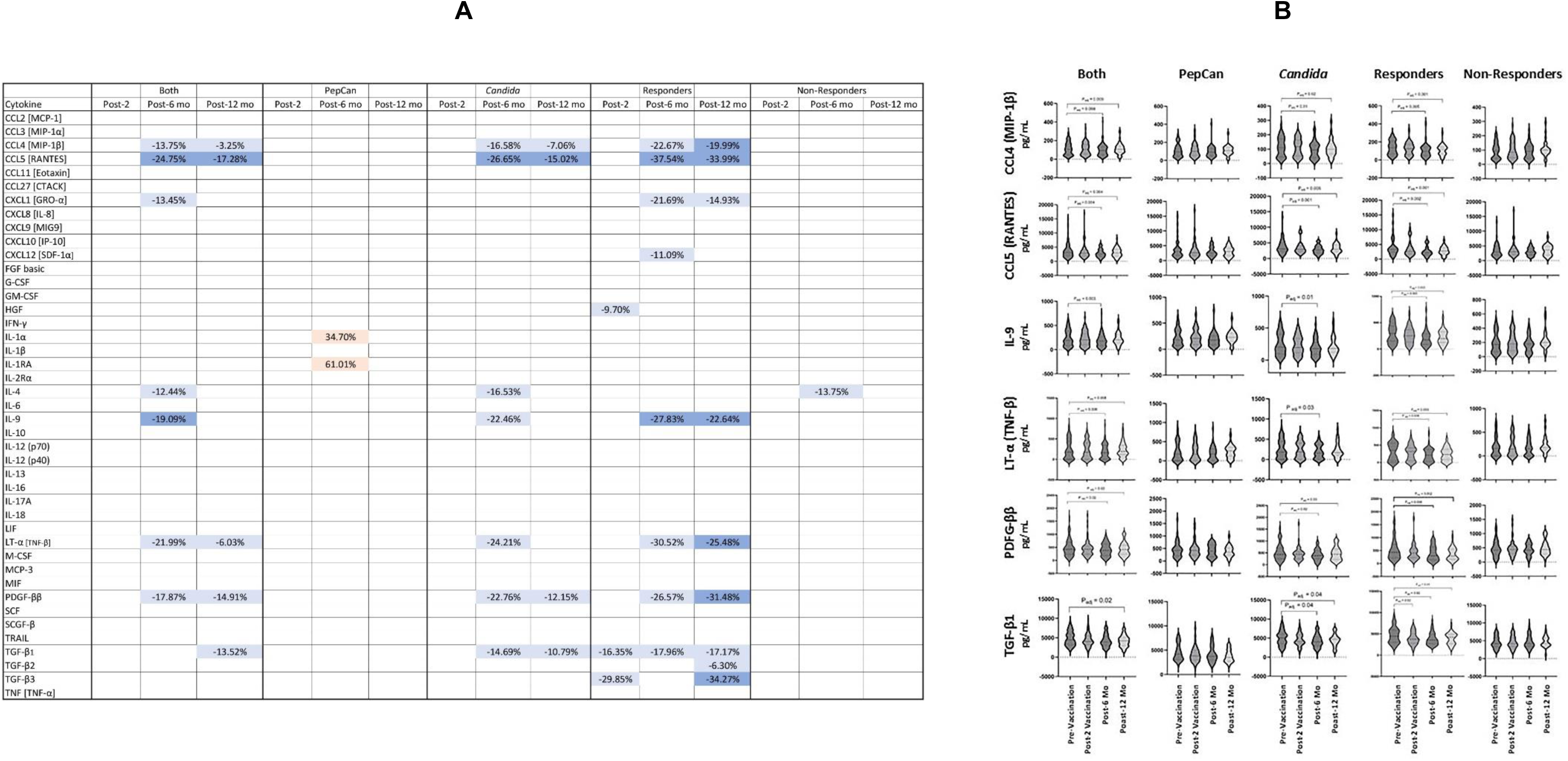
Plasma cytokine levels before and after vaccinations show that *Candida* may mediate CIN2/3 regression through altering key plasma cytokines. A. A heatmap of significant cytokine level changes (i.e., *P*_adj_<0.05) is shown comparing levels at post-2 vaccination, 6-month visit, and 12-month visit to the baseline. Light orange represents increased levels from the baseline with *P*_adj_ between 0.01 and 0.04. Light blue represents decreased levels from the baseline with *P*_adj_ between 0.01 and 0.04 while dark blue represents *P*_adj_ of <0.01. Eight of 51 cytokines that were undetectable in more than 50% of participants were excluded from the analysis (α-NGF, IFN-α2, IL-2, IL-3, IL-5, IL-7, IL-15, and VEGF), and are not shown. *P*-values determined using the Wilcoxon-rank sum test and were adjusted for multiple comparisons using the Benjamini-Hochberg method. B. Violin plots of 6 cytokines which were significantly decreased in the *Candida* group and in the histological responders.

The FACS analysis of peripheral immune cells for both groups revealed the percentage of Tregs was higher in histological non-responders in comparison to responders (stringent) pre-vaccination and at 12 months as reported in our Phase 1 study of PepCan;^22^ however, the comparisons were no longer significant after adjustments for multiple analyses in Phase 2 (Figure S4A). No changes in Th1 cells, Th2 cells, Tregs, and MDSCs following vaccinations that were consistent and significant after adjustments for multiple analyses were found (Figure S4D).

### HPV

When clearance of HPV infection at exit visit (6- or 12-month visit) by risk groups was examined, no significant differences were found between the PepCan and *Candida* groups (Figure 5A). For patients who were HPV 16-positive at the screening visit, the HPV 16 viral load was examined. A trend for HPV 16 viral load decreasing at the 6-month visit was observed in the *Candida* group, suggesting the HPV antigens may not be necessary for reducing viral loads (Figure 5B; *p*=0.05).

**Figure 5.**
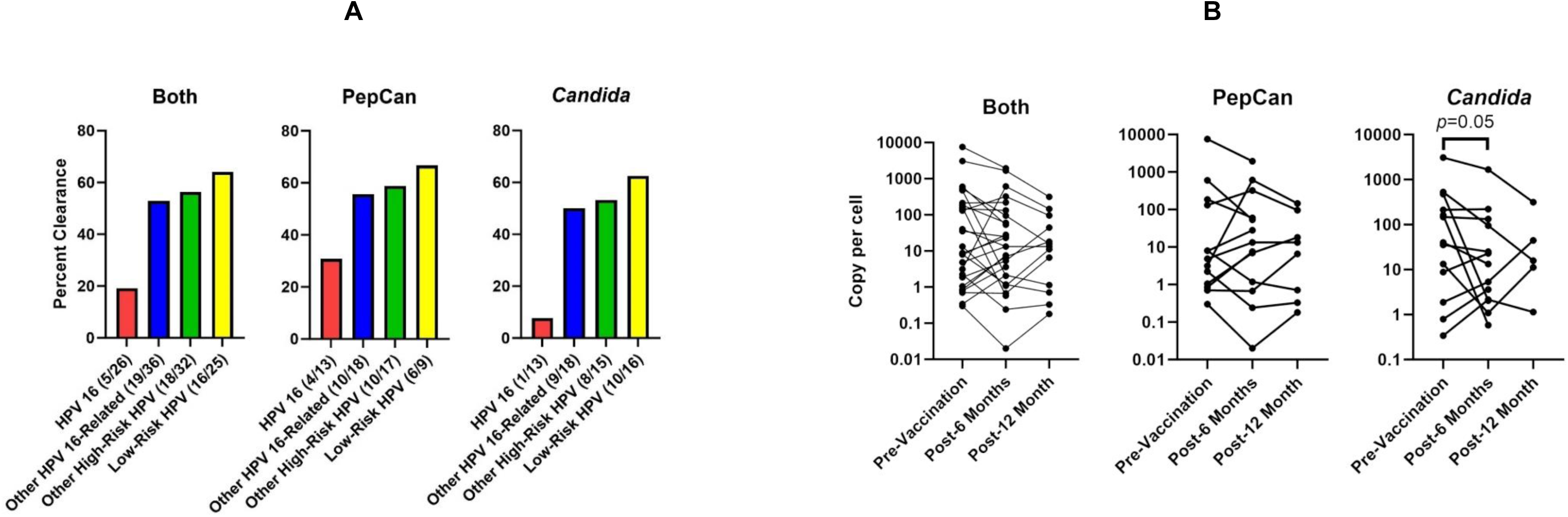
Analyses of HPV clearance by risk groups and HPV 16 viral load. A. The percentages of HPV clearance (i.e., detected at entry but not at the exit visit) by HPV types grouped as (1) HPV 16, (2) HPV 16-related types other than HPV 16 [i.e., HPV types 31, 33, 35, 52, 58, and 67], (3) high-risk HPV types not including HPV 16/HPV16-related [i.e., HPV types 18, 39, 45, 51, 53, 56, 59, 66, 68, 69, 70, 73, and 82], and (4) low-risk HPV types [i.e., HPV types 6, 11, 40, 42, 54, 61, 62, 71, 72, 81, 83, 84, and CP6108] are shown. B. HPV 16 viral loads in patients who were HPV 16-positive at entry. Wilcoxon matched-pairs signed rank test (two-sided) was used.

### HLA Typing

There were no significant associations with HLA types with histological responses (stringent) after correcting for multiple comparisons (Table S5). However, some HLA types were found at significantly higher proportions in the patient population compared to the general population. These included HLA B14, B15, B40, B49, C03, DR03, and DQ03. In particular, DQ03 was found to be nearly ten times more frequent in the patient population (38.1%) in comparison to the United States population (3.9%)(Figure S6D). The increased frequencies of HLA B14, B15, B40, C03, DR03, and DQ03 were also observed in our Phase 1 study.^22,23^ The increased risk for cervical neoplasia in people with DQ03 has also been reported by others.^30–32^

### Cervical Microbiome

No differences in pre-vaccination α diversity matrices between histological responders and non-responders (stringent) were found (Table S4A), and none of the taxa examined correlated with histological responses (Figure S5A). On the other hand, significant increases in α diversity from the baseline were observed at 6-month visit using the Faith’s Phylogenetic analysis in all subjects and subgroups examined [i.e., PepCan. *Candida*, histological responders (stringent) and histological non-responders](Table S4B). Furthermore, at 12-month visit, increases in α diversity from the baseline were observed using all three matrices (Pielou Evenness, Faith’s Phylogenetic, and Shannon Diversity) in all subjects and in subgroups (Table S4B). Therefore, α diversity increases significantly after vaccinations in a manner unrelated to vaccine efficacy.

Furthermore, the number of taxa were differentially expressed after vaccination in all patients as well as in the subgroups (Figure S5B, S5C, S5D, S5E, S5F, and Table S4C).

## DISCUSSION

CIN2/3 is a premalignant lesion of the uterine cervix that poses a significantly higher risk of progressing to malignancy in comparison to its lower grade counterpart, CIN1. While CIN2/3 lesions are usually treated with surgical resection such as LEEP, observation is desirable for women younger than 25 and women who are planning to become pregnant due to possible adverse obstetric outcomes.^5–8^ The availability of an effective HPV therapeutic vaccine that would promote regression of CIN without affecting the cervical anatomy is an unmet need; however, none has been approved by the Food and Drug Administration to date although many different versions have been tested in clinical trials.^33^

In this study, the effectiveness of PepCan and *Candida* were examined. Surprisingly, our findings revealed that only *Candida* was more effective in regressing CIN2/3 in comparison to a historical placebo group.^2^ Given that PepCan with higher amounts of HPV 16 E6 peptides was less effective in regressing CIN2/3 and in mounting anti-HPV T cell responses in our Phase 1 study,^22,23^ it may be possible that the HPV peptides in this formulation may have some unexpected inhibitory effects. Such effects can be inferred from our cytokine data which demonstrate much more robust changes in *Candida* group compared to the PepCan group (Figure 4). These results demonstrate that the adjuvant effect may be more important than the immune stimulation by the exogenous HPV antigens for therapeutic vaccines. It is possible that the immune system is targeting viral antigens from prior natural infection.

A comparison was made between the two treatment arms in the current study and the historical placebo^2^ using the stringent criterion of regression to no CIN. Therefore, the regression rates may seem low as many other studies use a less stringent criterion of regression to less than CIN2.^34^ The use of historical placebo has its limitations in that the study designs were similar but not exactly the same. For example, in our current study, quadrant biopsies were performed for the final histological assessment and the highest grade was resulted, while the comparison study only mentioned “biopsy” being performed.^2^ While the designated primary end points for both studies were 15 months after treatment initiation, the comparison study used a wider definition of “regression” in terms of the time of assessment as being “complete biopsy response (or endocervical curettage if biopsy not available) at 15, 9, 12, 21, or 27 months”.^2^ Therefore, the criterion used to define histological regression was much stricter in our current study. Significant demographic differences were observed in that the placebo group was younger and had higher use of oral contraceptives. Although the higher proportion of younger patients in the placebo group would expect to increase its regression rate, the *Candida* group still had a significantly higher rate of regression. While retention was excellent with 76 of 81 (93.8%) of enrolled patients completing the study, some exited after the 6-month visit due to persistent/progressive disease. While not allowing this option would have been unethical and unsafe, this lowered the number of patients completing the 12-month visit. Despite these concerns, *Candida* would be the chosen investigative product to proceed with a Phase 3 trial.

Safety of PepCan and *Candida* has been demonstrated, as there were no DLTs reported during the study. Interestingly, fever and myalgia were more frequently reported in the PepCan group suggesting that it was caused at least in part by the HPV peptides.

As demonstrated in our Phase 1 study,^22,23^ the presence of new T cell responses to HPV 16 E6 after vaccination was not correlated with histological responses. Putatively vaccine-specific T cells and vaccine-induced HPV-specific T cells were demonstrated in blood and cervix of both histological responders and non-responders (Figure 3A). Single-cell RNA-seq of vaccine-induced HPV-specific T cells demonstrated elevated expression of granzymes, CCR5, and EOMES in a histological responder (Figure 3C/D). The elevated levels of granzymes and EOMES are understandable since granzymes are serine proteases secreted by cytotoxic cells and EOMES is a T-bet-like T-box transcription factor expressed by natural killer cells and activated CD8 T cells.^35^ On the other hand, the elevated level of CCR5 is perplexing given the CCL4- and CCL5-reducing effects of *Candida*. Nevertheless, a larger number of patients are needed to confirm these findings. Furthermore, it is possible that in addition to the T cell phenotype, the local cervical environments may also influence histological response. For example, the number of Tregs in tumor microenvironment has been correlated with CIN2 progression.^36^

Another unexpected finding from this clinical trial was the exploratory role cytokines may be playing in inducing CIN2/3 regression including by chemokines, CCL4, and CCL5 (chemokines are small cytokines). CCL4 and CCL5 are thought to promote tumor progression by enhancing migration of inhibitory immune cells to tumor microenvironment, and promoting tumor metastasis.^37–39^ CCR5 (also known as CD195) is the main receptor of CCL4 and CCL5, and is expressed on T cells, smooth muscle endothelial cells, epithelial cells, parenchymal cells, and tumor cells. CCR5 and/or CXCR4 are used by HIV-1 to enter target immune cells, and are commonly targeted for developing anti-HIV therapies.^38^ More recently, several clinical trials have been initiated to treat multiple types of cancer utilizing CCR5 inhibitors with or without checkpoint inhibitors.^37,38^

IL-9, LT-α (formerly known as TNF-β), PDGF-ββ, and TGF-β1 were also significantly decreased in both the *Candida* and the histological responder groups after vaccinations (Figure 4). IL-9’s main role is to mount immune responses against parasites, but has also been shown to induce the proliferation of hematological malignancies, including Hodgkin’s lymphoma.^40^ LT-α is produced by activated lymphocytes and it has been shown to promote tumor angiogenesis in head and neck squamous cell carcinoma by modulating glycolysis in a PFKFB3 (6-phosphofructo-2-kinase/fructose-2,6-biphosphatase 3 enzyme)-dependent manner.^41^ An inhibitor of PFKFB3, PFK15, has been shown to impair tumor angiogenesis *in vitro* and *in vivo*.^41^ PDGF-ββ, a platelet-derived growth factor family protein, is expressed by cancer-associated fibroblasts, and can promote tumor progression through increased proliferation, migration, invasion, chemotaxis, and tumor angiogenesis. PDFG-ββ also has a role in promoting lymphatic metastasis of cholangiocarcinoma.^42^ PDFG-ββ induces the GSK3β and P65 signaling pathways. A study by Yan and colleagues showed that by inhibiting the GSK3β pathway with CHIR-98014 and the P65 signaling pathway with PG490, cancer-associated fibroblast induced promotion of tumor growth can be prevented *in vivo*.^42^ TGF-β signaling is involved in numerous cellular processes in embryogenesis and mature organisms, and plays key roles in cancer progression. One isoform, TGF-β1, is an inhibitory cytokine secreted by Tregs to inhibit other T cells,^43^ and also B cells, macrophages, and monocytes.^44^ TGF-β1, TGF-β2, and/or TGF-β3 have been targeted for cancer therapies in multiple clinical trials using small molecule inhibitors of TGF-β receptor kinases, antibodies, and a bifunctional fusion protein consisting of the extracellular domain of the TGF-βRII receptor (which would trap TGF-β) fused to an anti-PD-L1 antibody.^45^

*In vitro*, *Candida* has been shown to induce T cell proliferation and secretion of IL-12.^21,46^ However, we did not observe an increase in peripheral Th1 cells in this study, like we did in the Phase 1 study.^22,23^ *Candida* injected intradermally in periphery has been shown to induce a decrease in HPV 16 viral load (Figure 4B). This may be mediated through cytokine/chemokine changes induced by *Candida* which in turn could possibly alter the phenotype of HPV-specific T cells to be more effective in eliminating HPV infection.

It is perplexing as to why *Candida* alone was effective in regressing CIN2/3 when compared to a historical placebo while PepCan, which consists of HPV 16 E6 peptides and *Candida*, was not. This is particularly true when another version of a long peptide-based therapeutic HPV vaccine emulsified with incomplete Freund’s adjuvant seemed to show efficacy in treating high-grade vulvar intraepithelial lesions^47^ as well as HPV-associated oropharyngeal cancer in combination with an anti-PD1 checkpoint inhibitor.^48^ Such a conundrum has also been reported for neoantigen-based cancer vaccines for solid tumors. Rappaport and colleagues tested a shared neoantigen vaccine (a chimp adenovirus and self-amplifying mRNA) in combination with checkpoint inhibitors in treating solid tumors in patients with tumors expressing HLA-matched tumor mutations, mostly *KRAS*.^49^ Despite the fact post-vaccination T cell responses were demonstrated to the particular mutated antigens the patients have, 79% (15 of 19) of patients experienced progressive disease within a few months. On the other hand, personalized RNA neoantigen vaccines given to pancreatic ductal adenocarcinoma patients in conjunction with a checkpoint inhibitor and a four-drug chemotherapy regimen demonstrated longer median recurrence-free survival in patients with vaccine-expanded T cells in comparison to patients without such T cell expansion (*p*=0.003).^50^ In both of these comparisons, variables in patient characteristics, vaccine design, dosage, number of injections, treatment regimens may have contributed to the different outcomes. It would be difficult to tease out which variables are important among so many. In the case of peptide-based vaccines, cancer vaccine-induced T cells often do not eradicate tumors. When studied in mice,^51^ the authors reported that gp100 melanoma peptide in incomplete Freund’s adjuvant primed tumor-specific T cells accumulated at the vaccination site (not the tumor), became dysfunctional, and underwent antigen-mediated interferon-ɣ and Fas ligand-mediated apoptosis. “Persistent vaccine depots” have been shown to induce specific T cell segregation, malfunction and deletion. We have previously reported on the insoluble nature of the vaccine peptides in PepCan resulting the formation of microparticles,^23^ and it is possible that this created the depot effect resulting in lack of clinical efficacy.

As we have demonstrated possible efficacy of *Candida* for treating CIN2/3 in comparison to a historical placebo group, the next step would be to perform a Phase 3 study. However, a placebo-controlled study may be considered unethical due to the risk of disease progression of CIN3.^6,10,52^ Another alternative is to explore whether *Candida* could be more potent by increasing the volume and/or the number of injections. A multi-center Phase 2 study using *Candida* to treat common warts has demonstrated 65.9% efficacy (29/44) at the 0.3 mL dose and 79.5% efficacy (31/39) at the 0.5 mL dose in regressing the largest injected wart.^53^ This is a 21% dose-dependent increase in efficacy. Therefore, it would be possible to examine *Candida* as an alternative to standard surgical treatments in a non-inferiority trial. According to a recent meta-analysis, the success rate of the standard surgical treatments is 71.4%, given 25.9% positive margin rate after LEEP, and 3.7% early recurrence within 18 months.^54^ This would possibly make *Candida*, with an expected enhanced efficacy of 70.8% for regression to CIN1 or no CIN, a first-line option for treating CIN2/3.

In conclusion, we have demonstrated that *Candida* alone was likely effective in regressing CIN2/3 in comparison to a historical placebo group. Furthermore, *Candida* appears to possibly exert such an effect through reducing systemic cytokines which have been shown to promote tumor progression. Some of these cytokines may have direct effects to the tumor environments, but some may alter the phenotypes of the HPV-specific T cells as demonstrated by the single-cell analyses. We did not identify any predictive biomarkers amongst plasma cytokines, plasma metabolomics, and cervical microbiota. The vaccine-induce HPV-specific CD3 T cell responses were demonstrated in PepCan and *Candida* treated groups as well as in histological responders and non-responders suggesting that the induction of HPV-specific T cells alone is not sufficient for high-grade squamous intraepithelial regression. *Candida* may become an alternative to standard surgical treatments for CIN2/3. Furthermore, its addition to treatment of other premalignant and malignant diseases should be considered given its possible effects in reducing tumor-promoting chemokines/cytokines.

## Data Availability

All data produced are available online at GenBank.

https://www.ncbi.nlm.nih.gov/bioproject/PRJNA1201757

## CONTRIBUTIONS

MN and WG conceptualized the study; MN, TE, HC, JC, ND, AJ, HK, BJL, YCL, KM, CMQ, SS, TS, HRW, and WG participated in data acquisition; TE, MB, HC, JC, ND, JLF, BJL, YCL, KM, IN, DU, MR, SS, and TS performed data analysis; MN drafted the original version of the manuscript, and all authors edited drafts and approved the final version of the manuscript.

## ACKNOWLEDGEMENTS

The authors would like to thank James Allred, Angela Trammel, and Nicole Chapman for their clinical trial expertise, Tina Butler, Susan Miller, and Kimberly Spickes for their gynecologic expertise, Patricia Gminski and Traci Ireland for their administrative expertise, Laura Adkins, Brenda Gannon, Joseph A. Holley, Sorena Lo, and Larry Parker for their regulatory expertise, Amy Crisp and Jennifer Roberts for their pharmaceutical expertise, Yang Ou for her technical expertise, and Horace (Trey) Spencer, III and Akul Shrivastava for their assistance with data analysis. In addition, the authors would like to thank the patients for participating in this clinical trial.

