## Appendix for "A Randomized Double-Blind Phase 2 Clinical Trial Treating Cervical Intraepithelial Neoplasia 2/3 with PepCan or *Candida*"

### SUPPLEMENTARY APPENDIX

A Peptide-Based Human Papillomavirus Therapeutic Vaccine, PepCan, or *Candida* Adjuvant Alone in Cervical Intraepithelial Neoplasia 2/3

#### TABLE OF CONTENTS

Supplementary Methods 4
HPV Typing and HPV 16 Viral Load
Peripheral HPV 16-Specific T Cell Responses
Epitope Spreading
Peripheral and Cervical Immune Responses
Peripheral Immune Cells
Plasma Cytokines
Plasma Metabolomics
Cervical Microbiome
HLA Typing

Supplementary Tables 14
Table S1A. All adverse events regardless of treatment-relatedness by event.
Table S1B. All adverse events regardless of treatment-relatedness by patient.
Table S2A. Entry cytokine levels by histological response groups (stringent) for patients who completed the 6-month visit in both treatment groups.
Table S2B. Entry cytokine levels by histological response groups (stringent) for patients who completed the 6-month visit in the PepCan group.
Table S2C. Entry cytokine levels by histological response groups (stringent) for patients who completed the 6-month visit in the *Candida* group.
Table S3A. Entry metabolite levels by histological response groups (stringent) for patients who completed the 6-month visit in both treatment groups.
Table S3B. Entry metabolite levels by histological response groups (stringent) for patients who completed the 6-month visit in the PepCan group.
Table S3C. Entry metabolite levels by histological response groups (stringent) for patients who completed the 6-month visit in the *Candida* group.
Table S4A. Summary of comparisons between α diversity of histological responders and non-responders (stringent) for patients who completed the 6-month visit.
Table S4B. Summary of comparisons between α diversity between before and after vaccinations in all patients and in subgroups.
Table S4C. The top five differentially abundant taxa after vaccinations in all patients and in subgroups.
Table S5A. Comparison of responders to non-responders (stringent) with respect to HLA types with total allele counts of 10 or greater for both groups.
Table S5B. Comparison of responders to non-responders with respect to HLA types with total allele counts of 10 or greater for the PepCan group.
Table S5C. Comparison of responders to non-responders with respect to HLA types with total allele counts of 10 or greater for the *Candida* group.
Table S6. Summary of vaccine-induced HPV-specific T cells detected in cervix.

Supplementary Figures 51
Figure S1. Trial design.
Figure S2. Histological responses and subgroup comparisons.
Figure S3. Representative ELISPOT assay results.
Figure S4. Peripheral immune cell analyses.
Figure S5. Differential abundance of taxa.
Figure S6. Observed HLA frequencies for patients in comparison to the estimates for the United States population based on racial distributions.

Supplementary References 75

#### SUPPLEMENTARY METHODS

*HPV TYPING AND HPV 16 VIRAL LOAD*

HPV typing testing was performed using a Linear Array HPV Genotyping Test (Roche Diagnostics, Indianapolis, IN) according to the manufacturer's instructions using Thin-Prep samples (Hologic, Marlborough, MA). This test detects 37 anogenital HPV types: 6, 11, 16, 18, 26, 31, 33, 35, 39, 40, 42, 45, 51, 52, 53, 54, 55, 56, 58, 59, 61, 62, 64, 66, 67, 68, 69, 70, 71, 72, 73 [MM9], 81, 82 [MM4], 83 [MM7], 84 [MM8], IS39, and CP6108. The human β-globin signal was also assayed as a positive control for sample adequacy of DNA content. Subjects were categorized for being positive for HPV-16, other HPV-16-related types (31, 33, 35, 52, 58, and 67),[1] other high-risk HPV types (18, 39, 45, 51, 53, 56, 59, 66, 68, 69, 70, 73, and 82),[2] and low-risk HPV types (6, 11, 40, 42, 54, 61, 62, 71, 72, 81, 83, 84, and CP6108).[1,2] The difference in HPV clearance rates between the PepCan and *Candida* groups were examined using Fisher's exact test as described in the statistical analysis plan.

A method previously described was followed[3,4] to determine the HPV 16 viral load in subjects in whom HPV 16 was detected at the screening visit. Briefly, quantitative real time PCR was performed using iQ SYBR Green PCR Master Mix (BioRad Laboratories, Hercules, CA) with diluted DNA and HPV 16 E6 primers (5′-aaagccactgtgtcctgaaga-3′ and 5′-ctgggtttctctacgtgttct-3′)[5] or GAPDH primers (5′-cgagatccctccaaaatcaa-3' and 5-catgagtccttccacgataccaa-3').[6] Amplifications were carried out using a BioRad CFX 96 Real-Time PCR Detection System (BioRad Laboratories) with an initial denaturation at 95°C for 5 min. This was followed by 40 cycles of 95°C for 10 sec and 55°C for 20 sec for HPV 16 E6 or 30 cycles of 95°C for 15 sec, 50°C for 30 sec, and 72°C for 20 sec for GAPDH. Data acquisition was done at 510 nm at 55°C and 72°C respectively. The standard curve for HPV was established by amplification of a serially diluted quantitative synthetic HPV 16 DNA (American Type Culture Collection, Manassas, VA) in the presence of 100pg of human placental DNA. The standard curve for GAPDH was established using serial 10-fold dilution of placental DNA starting with 250ng of DNA. The amount of DNA was converted to cell number with an assumption that 6.6pg of DNA is present in a diploid cell.[4] Mean threshold cycle (CT) values of triplicate samples at each dilution were used to generate standard curves, and mean CT values of specimen duplicates were used for calculating quantities. All DNA samples from the same subjects were analyzed in the same PCR run. HPV 16 viral loads were determined using E6 copy numbers and were not meant to be an absolute quantification. The specificity of amplifications was examined through melting curve analyses. The significance in the changes of HPV 16 viral load was examined using Wilcoxon matched-pairs signed rank test as described in the statistical analysis plan.

*PERIPHERAL HPV 16-SPECIFIC T CELL RESPONSES*

Peripheral T cell responses specific to the HPV 16 E6 protein were assessed using an enzyme-linked immunospot (ELISPOT) assay with blood samples drawn before and after vaccinations (pre-vaccination, post-2 vaccination, at 6 month visit, and at 12 month visit). Peripheral blood mononuclear cells (PBMCs) were isolated from heparinized whole blood using a Ficoll density-gradient centrifugation method and were then cryopreserved. CD3 T cell lines were established from three or four blood draws from each patient as described previously by stimulating magnetically selected CD3 T cells with autologous monocyte-derived dendritic cells pulsed with recombinant vaccinia virus and recombinant glutathione *S*-transferase-fusion protein expressing HPV16 E6.[3,7] The ELISPOT assay was performed on day 22 in which HPV 16 E6 (10 regions) were assessed using a method adapted from a previously published version.[3,7] Briefly, 96-well plates (MultiScreen-MSIPS 4510 plates, Sigma-Aldrich, St. Louis MO) were treated with 35% EtOH in sterile PBS and rinsed immediately. They were subsequently coated with 5 μg/ml of primary anti-interferon γ monoclonal antibody (1-D1K, Mabtech, Sweden), washed, and blocked. Then, 2.5 x $10^4$ CD3 T cells were added per well in triplicate along with pooled peptides (10 μM per peptide). Each peptide pool contained 3 overlapping HPV 16 E6 15-mer peptides. For example, the E6 1-25 pool contained E6 1-15, E6 6-20, and E6 11-25 peptides. Negative control wells contained media only, and positive control wells contained phytohaemagglutinin at 10 μg/mL, (Remel, Lenexa, KS). Plates were incubated for 24 hours at 37°C with 5% carbon dioxide. After washing, a secondary biotin-conjugated anti-Interferon γ monoclonal antibody (7-B6-1, Mabtech, Nacka Strand, Sweden) was added at 1 μg/ml and incubated for 2 hours. After washing, streptavidin-horseradish peroxidase (Mabtech) was added at a dilution of one to 750 and incubated at room temperature for one hour. Wells were washed and coated with 3,3',5,5' tetramethylbenzidine (Mabtech) for 10 to 30 minutes and allowed to develop at room temperature until spots emerged. The plates were then rinsed in water and allowed to dry overnight. Peptide pools with spot forming units twice or greater to the media only control were considered to be positive. Paired *t*-test was used to examine the significance of increased responses as described in the statistical analysis plan.

*EPITOPE SPREADING*

For patients who were HPV 16-positive at entry, epitope spreading to HPV 16 E7 protein was examined using ELISPOT assay as described above. For those who were HPV 18, 35, or 52-positive at entry, epitope spreading to respective E6 protein was similarly examined. These HPV types were selected for their high prevalence in earlier studies examining the patient population.[8] Recombinant vaccinia virus constructs expressing HPV 35 E6 protein and HPV 52 E6 protein separately were constructed as well as recombinant GST fusion proteins containing the E6 protein of HPV types 18, 35, and 52 separately. The full-length of HPV 18 E6 gene was amplified by polymerase chain reaction (PCR) from the vector HPV 18-pBR322 (American Tissue Culture Collection, Manassas,VA). The HPV 18-pBR322 vector contains the full-length of HPV 18 genome. The amplified PCR product of HPV 18-E6 gene was cloned into pGEX-2T vector (GE Healthcare, Chicago, IL) at EcoRI site, introduced at 5’ end and 3’ end of primer sequences. The full-length of HPV 35 E6 gene was amplified by PCR from the vector HPV 35-1-

pBR322 (provided by Carina Eklund, Karolinska Institutet, Stockholm, Sweden), which contains partial fragment of HPV 35 genome including the HPV 35 E6 gene. The amplified PCR product of HPV 35 E6 gene was then cloned into pGEX-2T vector at 5' BamHI site and 3' EcoRI site, introduced via primer sequence design. In a similar way, the full-length of HPV 52 E6 gene was amplified by PCR from the vector HPV 52-pBR322 (ATCC). The HPV 52-pBR322 contains the full-length of HPV 52 genome. The amplified PCR product of HPV 52 E6 gene was cloned into pGEX-2T vector at EcoRI sites, added at 5' end and 3' end of primer sequences. The primer sequences for application of HPV 18 E6 gene were 5' ATAGAATTCCTATGGCGCGCTTTGAGG 3' for forward and 5' GACGAATTCTTATACTTGTGTTTCTCTGCG 3' for reserves, and they contained an EcoRI sites; for HPV 35 E6 gene were 5' ACGTGGATCCATGTTTCAGGACCCAGC 3' for forward and 5' GTCAGAATTCCATGCATGATTACACCTCG 3' for reverse and contained BamHI and EcoRI sites; and for HPV 52 E6 gene were 5' TGACGAATTCCCATGTTTGAGGATCCAGC 3' for forward and 5' CACGGAATTCGTTACACTTGGGTCACAGG 3' for reverse and contained EcoRI sites. The insertion of the HPV 18-E6, HPV 35-E6, HPV 52-E6 in pGEX-2T vector was confirmed by restriction enzyme digestion and sequence analysis. Expression of the HPV 18-E6-GST, HPV 35-E6-GST, HPV 52-E6-GST fusion proteins in pGEX-2T vector was demonstrated with Western blot using goat-anti-GST polyclonal antibody (GE healthcare) with expected 44 kD in size.

*PERIPHERAL AND CERVICAL IMMUNE RESPONSES*

The bulk T cell receptor (TCR) β deep sequencing of PBMCs and liquid-based cytology samples was performed (Adaptive Biotechnologies, Seattle, WA) for patients who demonstrated new HPV 16 E6 responses in at least two of three post-vaccination PBMC samples (n=15). Putatively vaccine-specific T cells were identified by comparing post-vaccination PBMC sample to the pre-vaccination PBMC sample using a beta-binomial model.[9] The presence of such putatively vaccine-specific T cells were examined in cervix by searching for complementarity determining region 3 (CDR3) β nucleotide sequences in liquid-based cytology samples. In order to confirm whether these T cells were HPV-specific, T cells were stimulated with HPV 16 peptides from the region of statistically significantly increased ELISPOT positivity after vaccination and were sorted based on interferon-γ secretion (Interferon-γ Secretion Assay, Miltenyi Biotec, Auburn, CA) and CD8 staining (n=5). The CD8-positive and -negative interferon-γ secreting T cells were analyzed using single-cell RNA-seq and TCR-seq (10X Genomics, Pleasanton, CA).

The experimental procedures for the single-cell analyses have been published previously.[10] Briefly, post-vaccination PBMC samples were thawed and cultured overnight in RPMI medium with 5% human serum and 1,200 IU/mL of recombinant human interleukin-2 (PeproTech, Cranbury, NJ). The cells were stimulated for 3 hours with 10 μM of HPV peptides selected from positive ELISPOT results. interferon-γ secreting cells were labeled by the interferon-γ catch reagent and phycoerythrin (PE)-labeled interferon-γ detection antibody, according to manufacturer's instruction (interferon-γ Secretion Assay – Detection Kit, Miltenyi Biotec). interferon-γ secreting CD8-positive and interferon-γ secreting CD8-negative cells were enriched by FACS Aria III and subjected to single-cell sequencing analysis. Briefly, the manufacturer's protocol for single-cell sample preparation was followed using single cell 5' reagent kits v2 (10X Genomics, Pleasanton, CA). Single-cell sequencing for gene expression and TCR was performed using an Illumina Novaseq 6000 system. Sequencing data were first processed by a Cell Ranger pipeline (v6.1.2; 10X Genomics). Gene expression sequencing data were mapped to human reference (GRCh38-2020-A) dataset. The single-cell data were further analyzed by Seurat v.4.4.0, by following the recommended steps and settings. The low-quality cells and doublets were filtered out by the following setting: percentage of mitochondrial genes > 5%, number of detected genes < 1000 and number of detected genes > 5000. The clustering was performed with the resolution setting at 0.2. The UMAP (Uniform Manifold

Approximation and Projection) plot, violin plots, feature plots, and *p*-values were also generated by Seurat v4.4.0. TCR sequencing data were mapped to human TCR reference (GRCh38-alts-ensembl-5.0.0) dataset, and they were further analyzed by Loupe V(D)J Browser (v4.0.0; 10X Genomics). T cell clonotypes were defined based on TCR Vβ CDR3 amino acid sequences.

*PERIPHERAL IMMUNE CELLS*

Thawed PBMCs were stained with appropriate isotype controls and combinations of monoclonal antibodies to analyze T-helper type 1 (Th1, promoting cellular immunity), T-helper type 2 (Th2, promoting antibody production and is anti-Th1), regulatory T-cells (Tregs, promoting immune suppression), and myeloid-derived suppressor cells (MDSCs). As there is no consensus in the literature as to how to analyze MDSCs; two commonly used methods were used to measure MDSC1[11] and MDSC2.[12,13]

To analyze Th1 cells (CD4- and Tbet-positive), Th2 cells (CD4- and GATA3-positive), and Tregs (CD4-, CD25-, and FoxP3-positive), thawed PBMCs were stained using fluorescein isothiocyanate-labeled anti-human CD4 (clone RPA-T4, Thermo Fisher Scientific, Hillsboro, OR), phycoerythrin-labeled anti-human/mouse T-bet (clone 4B10, Thermo Fisher Scientific), PerCP-Cy5.5-labeled anti-human CD25 (clone BC96, Thermo Fisher Scientific), allophycocyanin-labeled anti-human Foxp3 (clone PCH101, Thermo Fisher Scientific ), and phycoerythrin -Cy7 labeled anti-human/mouse GATA3 (clone L50-823, BD, Franklin, NJ). Cells were initially stained with antibodies for surface markers CD3, CD4, and CD25.  Intracellular staining for T-bet, GATA3, and Foxp3 was performed using the Foxp3 staining kit (Thermo Fisher Scientific).

For the analyses of MDSC1 (CD33-, IL4Ralpha-, CD11b- CD14- positive and HLA-DR-intermediate/negative) and MDSC2 (CD14-positive and HLA-DR-low/negative), thawed PBMCs were treated with Fc receptor blocker (Innovex Biosciences, Richmond, CA) first. Then, they were stained with anti-CD11b/Mac-1 Pacific Blue (BD), anti-CD14-APC-Cy7 (BD), anti-CD33-

APC (BD), anti-IL4Ralpha (R&D, Minneapolis, MN) and anti-HLA-DR Brilliant Violet 711 (Biolegend, San Diego, CA).

Flow cytometric analysis was performed with FACS Fortessa using FACS Diva software (BD) in the University of Arkansas for Medical Sciences Microbiology and Immunology Flow Cytometry Core Laboratory. One hundred thousand events were acquired in the lymphocyte gate for Th1, Th2, and Treg determinations.[14] For MDSC1 and MDSC2, all events excluding debris and doublets were acquired. The values before and after vaccinations were compared using paired *t*-test, and those between histological responders and non-responders (stringent) were compared using unpaired *t*-test as descried in the statistical analysis plan.

*PLASMA CYTOKINES*

Plasma samples were obtain by centrifugation of whole blood collected in sodium heparin tubes, and frozen in a -80°C freezer within one hour after blood draw as our previous work demonstrated that levels of some cytokines were altered significantly at two hours.[3] Data from one sample which was received after one hour but within two hours was utilized for cytokines shown not to be labile.[3] The quantities of β-nerve growth factor (β-NGF), cutaneous T cell-attracting chemokine (CTACK), eotaxin, basic fibroblast growth factor (FGF), granulocyte-colony stimulating factor (G-CSF), granulocyte monocyte-colony stimulating factor (GM-CSF), chemokine (C-X-C motif) ligand 1 (CXCL1, also known as GRO-α), hepatocyte growth factor (HGF), interferon-α2 (IFN-α2), interferon-γ (IFN-γ), interleukin-1α (IL-1α), interleukin-1β (IL-1β), IL-1 receptor agonist (IL-1RA), interleukin-2 (IL-2), interleukin-2Rα, (IL-2Rα), interleukin-3 (IL-3), interleukin-4 (IL-4), interleukin-5 (IL-5), interleukin-6 (IL-6), interleukin-7 (IL-7), interleukin-8 (IL-8), interleukin-9 (IL-9), interleukin-10 (IL-10), interleukin-12 (IL-12) (p70), IL-12 (p40), interleukin-13 (IL-13), interleukin-15 (IL-15), interleukin-16 (IL-16), interleukin-17A (IL-17A), interleukin-18 (IL-18), IFN-γ induced protein 10 (IP-10), leukemia inhibitory factor (LIF), monocyte-colony stimulating factor (M-CSF), monocyte chemotactic protein 1 (MCP-1), monocyte chemotactic protein 3 (MCP-3), macrophage migration inhibitory factor (MIF), chemokine (C-X-C motif) ligand 9 (CXCL9, also known as MIG), macrophage inflammatory protein1α (MIP-1α), macrophage inflammatory protein-1β (MIP-1β), platelet-derived growth factor subunit B (PDGF-BB), regulated on activation, normal T-cell expressed and secreted (RANTES), stem cell factor (SCF), stem cell growth factor β (SCGF-β), stromal-derived cell factor-1α (SDF-1α), tumor necrosis factor α (TNF-α), tumor necrosis factor β (TNF-β), tumor necrosis factor-related apoptosis inducing ligand, (TRAIL), and vascular endothelial growth factor (VEGF) were determined using a commercially available Bio-Plex kit (Bio-Rad Laboratories) according to the manufacturer's instructions using a Bio-Plex 200 instrument (Bio-Rad Laboratories). Tumor growth factor-β1 (TGF-β1), tumor growth factor-β2 (TGF-β2), and tumor growth factor-β3 (TGF-β3) were determined separately using another Bio-Plex kit which included an acid treatment. The correlations between the base line cytokine level and histological responses (stringent) were analyzed as explained in the statistical analysis plan.

*PLASMA METABOLOMICS*

The replicate of samples used for cytokine analyses was analyzed for global profiling at Metabolon, Inc. (Morrisville, NC). Several recovery standards were added prior to the first step in the extraction process for quality control purposes. Proteins were precipitated with methanol under vigorous shaking for 2 minutes followed by centrifugation for their removal. The resulting extract was divided into five fractions. Two fractions were used for analysis by two separate reverse phase ultrahigh performance liquid chromatography tandem mass spectroscopy methods with positive ion mode electrospray ionization. One fraction was used for analysis by reverse phase ultrahigh performance liquid chromatography tandem mass spectroscopy methods with negative ion mode electrospray ionization. Another fraction was used for analysis by hydrophilic interaction liquid chromatography ultrahigh performance liquid chromatography tandem mass spectroscopy with negative ion mode electrospray. The last fraction was reserved for backup. Several types of controls were analyzed in conjunction with the experimental samples. A pooled matrix sample made by taking a small volume of each experimental sample (or alternatively, use of a pool of well-characterized human plasma) served as a technical replicate throughout the data set. Extracted water samples served as process blanks. A panel of quality control standards that were carefully chosen not to interfere with the determination of endogenous compounds were added into every analyzed sample. This allowed instrument performance monitoring, and chromatographic alignment. The correlations between individual baseline metabolite and histological responses (stringent) were examined as outlined in the statistical analysis plan.

*CERVICAL MICROBIOME*

The cervical cytology specimens in this current study were collected and reserved in the vial of the ThinPrep Pap Test (Hologic) as previously described.[10,15] The specimens were frozen (-80ºC) on the day of collection. DNA was extracted using QIAamp DNA Midi Kit (Qiagen).[10] A positive control utilized in this study was a mock vaginal microbial community composed of a mixture of genomic DNA from the American Type Culture Collection (Manassas, VA). The negative control was ThinPrep preservation solution without the sample as blank extraction.[16] Controls and the extracted DNA were sent to Argonne National Laboratory (Argonne,IL) for amplification and sequencing of the 16S rRNA gene on an Illumina MiSeq sequencing platform.[17] The same volume of DNA was used for each reaction, and then normalized at the PCR pooling step. This ensures that equal amounts of each amplified sample are added to the sequencing pool. Paired-end reads from libraries with ~250-bp inserts were generated for the V4 region using the barcoded primer set: 515FB: 5'-GTGYCAGCMGCCGCGGTAA-3' and 806RB: 5'-GGACTACNVGGGTWTCTAAT-3'.[18-22] MiSeq Reagent Kit v2 (2 × 150 cycles, MS-102-2002) was used. Initial sequence processing and analyses were performed using QIIME 2.[23] Correlation between $\alpha$ diversity of pre-vaccination samples and vaccine response (strict) and that of individual taxon and vaccine response (strict) were examined as explained in the statistical analysis plan.

#### HLA TYPING

Low-resolution typing for HLA class I A, B, and C and class II DRB1, DQB1, and DPB1 was performed with MicroSSP Generic DNA Typing Trays (One Lambda, Los Angeles, Ca), using DNA extracted using QIAamp DNA Mini Kit (Qiagen, Germantown, MD) from PBMCs. The typing results were analyzed using HLA Fusion (One Lambda). The correlations between HLA types and histological response (stringent) and between the expected HLA frequencies among the United States population and the patient population were examined as explained in the statistical analysis plan.

### SUPPLEMENTARY TABLES

#### TABLE S1A. All adverse events regardless of treatment-relatedness by event.

| **All Adverse Events at the Event Level by Treatment and Grade** | | **CTCAE v4.03 Adverse Event Severity Scale Grade** | | | | | | | | | | | |
|---|---|---|---|---|---|---|---|---|---|---|---|---|---|
| | | **PepCan** | | | | | | ***Candida*** | | | | | |
| **CTCAE v4.03 System Organ Class** | **CTCAE v4.03 Adverse Event Term** | **1** | **2** | **3** | **4** | **5** | **Total** | **1** | **2** | **3** | **4** | **5** | **Total** |
| Blood and Lymphatic System Disorders [17]$^{u}$ | Anemia [17]$^{u}$ | 9$^{u}$ | - | - | - | - | 9$^{u}$ | 8$^{u}$ | - | - | - | - | 8$^{u}$ |
| Cardiac Disorders [7]$^{u}$ | Chest Pain - Cardiac [1]$^{u}$ | - | - | - | - | - | - | 1$^{u}$ | - | - | - | - | 1$^{u}$ |
| | Palpitations [4]$^{u}$ | 1$^{u}$ | - | - | - | - | 1$^{u}$ | 3$^{u}$ | - | - | - | - | 3$^{u}$ |
| | Sinus Bradycardia [1]$^{u}$ | - | - | - | - | - | - | 1$^{u}$ | - | - | - | - | 1$^{u}$ |
| | Sinus Tachycardia [1]$^{u}$ | 1$^{u}$ | - | - | - | - | 1$^{u}$ | - | - | - | - | - | - |
| Ear and Labyrinth Disorders [3]$^{u}$ | Ear Pain [3]$^{u}$ | 2$^{u}$ | 1$^{u}$ | - | - | - | 3$^{u}$ | - | - | - | - | - | - |
| Endocrine Disorders [3]$^{u}$ | Hypothyroidism [3]$^{u}$ | - | - | - | - | - | - | - | 3$^{u}$ | - | - | - | 3$^{u}$ |
| Eye Disorders [2]$^{u}$ | Photophobia [1]$^{u}$ | - | 1$^{u}$ | - | - | - | 1$^{u}$ | - | - | - | - | - | - |
| | Eye Disorders - Other, Specify [1]$^{u}$ | - | - | - | - | - | - | 1$^{u}$ | - | - | - | - | 1$^{u}$ |
| Gastrointestinal Disorders [93]$^{u53}$ | Abdominal Pain [8]$^{u7}$ | 5$^{u4}$ | 1$^{u}$ | - | - | - | 6$^{u5}$ | 2$^{u}$ | - | - | - | - | 2$^{u}$ |
| | Bloating [1]$^{u}$ | - | - | - | - | - | - | 1$^{u}$ | - | - | - | - | 1$^{u}$ |

| | | | | | | | | | | | | | |
|---|---|---|---|---|---|---|---|---|---|---|---|---|---|
| | Colitis [1][u] | - | - | - | - | - | - | - | - | 1[u] | - | - | 1[u] |
| | Constipation [8][u] | 5[u] | 1[u] | - | - | - | 6[u] | 2[u] | - | - | - | - | 2[u] |
| | Diarrhea [2][u] | - | - | - | - | - | - | 2[u] | - | - | - | - | 2[u] |
| | Gastritis [1][u] | - | 1[u] | - | - | - | 1[u] | - | - | - | - | - | - |
| | Gastroesophageal Reflux Disease [1][u] | - | 1[u] | - | - | - | 1[u] | - | - | - | - | - | - |
| | Gastrointestinal Pain [2][u] | 1[u] | - | - | - | - | 1[u] | - | - | 1[u] | - | - | 1[u] |
| | Gastroparesis [1][u] | 1[u] | - | - | - | - | 1[u] | - | - | - | - | - | - |
| | Nausea [53][u16] | 30[u11] | 1[u] | - | - | - | 31[u12] | 21[u3] | 1[u] | - | - | - | 22[u4] |
| | Stomach Pain [3][u] | 1[u] | - | - | - | - | 1[u] | 1[u] | 1[u] | - | - | - | 2[u] |
| | Vomiting [4][u3] | 3[u2] | - | - | - | - | 3[u2] | - | - | 1[u] | - | - | 1[u] |
| | Gastrointestinal Disorders - Other, Specify [8][u7] | - | 4[u] | - | - | - | 4[u] | 2[u1] | 2[u] | - | - | - | 4[u3] |
| General Disorders and Site Administration Conditions [355][u33] | Edema Limbs [1][u] | - | - | - | - | - | - | 1[u] | - | - | - | - | 1[u] |
| | Fatigue [18][u8] | 9[u5] | - | - | - | - | 9[u5] | 9[u3] | - | - | - | - | 9[u3] |
| | Fever [19][u6] | 12[u3] | 2[u1] | - | - | - | 14[u4] | 5[u2] | - | - | - | - | 5[u2] |
| | Flu-Like Symptoms [13][u3] | 5 | 3[u1] | - | - | - | 8[u1] | 3 | 2[u] | - | - | - | 5[u2] |
| | Injection Site Reaction [288] | 86 | 56 | - | - | - | 142 | 97 | 49 | - | - | - | 146 |
| | Localized Edema [1][u] | - | - | - | - | - | - | 1[u] | - | - | - | - | 1[u] |
| | Malaise [1] | 1 | - | - | - | - | 1 | - | - | - | - | - | - |

| | | | | | | | | | | | | | |
|---|---|---|---|---|---|---|---|---|---|---|---|---|---|
| | Non-Cardiac Chest Pain [2][u] | - | - | - | - | - | - | 2[u] | - | - | - | - | 2[u] |
| | Pain [12][u] | 3[u] | 4[u] | 1[u] | - | - | 8[u] | 2[u] | 1[u] | 1[u] | - | - | 4[u] |
| Immune System Disorders [1][u] | Allergic Reaction [1][u] | - | 1[u] | - | - | - | 1[u] | - | - | - | - | - | - |
| Infections and Infestations [47][u, 1 SAE] | Appendicitis [1][u, SAE] | - | - | - | - | - | - | - | - | 1[u, SAE] | - | - | 1[u, SAE] |
| | Laryngitis [1][u] | - | 1[u] | - | - | - | 1[u] | - | - | - | - | - | - |
| | Pharyngitis [1][u] | - | 1[u] | - | - | - | 1[u] | - | - | - | - | - | - |
| | Sinusitis [2][u] | - | 2[u] | - | - | - | 2[u] | - | - | - | - | - | - |
| | Skin Infection [3][u] | 1[u] | 1[u] | - | - | - | 2[u] | 1[u] | - | - | - | - | 1[u] |
| | Tooth Infection [1][u] | - | 1[u] | - | - | - | 1[u] | - | - | - | - | - | - |
| | Upper Respiratory Infection [10][u] | - | 9[u] | - | - | - | 9[u] | - | 1[u] | - | - | - | 1[u] |
| | Urinary Tract Infection [11][u] | - | 7[u] | - | - | - | 7[u] | - | 4[u] | - | - | - | 4[u] |
| | Vaginal Infection [10][u] | - | 5[u] | - | - | - | 5[u] | - | 5[u] | - | - | - | 5[u] |
| | Infections and Infestations - Other, Specify [7][u] | - | 3[u] | - | - | - | 3[u] | 1[u] | 3[u] | - | - | - | 4[u] |
| Injury, Poisoning and Procedural Complications [6][u3] | Bruising [6][u3] | 4[u2] | - | - | - | - | 4[u2] | 2[u1] | - | - | - | - | 2[u1] |

| | | | | | | | | | | | | | |
|---|---|---|---|---|---|---|---|---|---|---|---|---|---|
| Investigations [60][u] | Alanine Aminotransferase Increased [13][u] | 8[u] | - | - | - | - | 8[u] | 5[u] | - | - | - | - | 5[u] |
| | Alkaline Phosphatase Increased [3][u] | 2[u] | - | - | - | - | 2[u] | 1[u] | - | - | - | - | 1[u] |
| | Aspartate Aminotransferase Increased [14][u] | 7[u] | - | - | - | - | 7[u] | 7[u] | - | - | - | - | 7[u] |
| | Blood Bilirubin Increased [3][u] | 2[u] | - | - | - | - | 2[u] | 1[u] | - | - | - | - | 1[u] |
| | Creatinine Increased [9][u] | 4[u] | 1[u] | - | - | - | 5[u] | 4[u] | - | - | - | - | 4[u] |
| | Hemoglobin Increased [6][u] | 4[u] | - | - | - | - | 4[u] | 2[u] | - | - | - | - | 2[u] |
| | Platelet Count Decreased [3][u] | 1[u] | - | - | - | - | 1[u] | 2[u] | - | - | - | - | 2[u] |
| | Weight Loss [1][u] | 1[u] | - | - | - | - | 1[u] | - | - | - | - | - | - |
| | White Blood Cell Decreased [4][u] | 2[u] | - | - | - | - | 2[u] | 1[u] | 1[u] | - | - | - | 2[u] |
| | Investigations - Other, Specify [4][u] | 2[u] | - | - | - | - | 2[u] | 2[u] | - | - | - | - | 2[u] |
| Metabolism and Nutrition Disorders [57][u54] | Dehydration [1][u] | - | - | - | - | - | - | - | 1[u] | - | - | - | 1[u] |
| | Hyperglycemia [4][u] | 2[u] | - | 1[u] | - | - | 3[u] | 1[u] | - | - | - | - | 1[u] |
| | Hyperkalemia [1][u] | 1[u] | - | - | - | - | 1[u] | - | - | - | - | - | - |
| | Hypoalbuminemia [12][u] | 3[u] | - | - | - | - | 3[u] | 9[u] | - | - | - | - | 9[u] |

| | | | | | | | | | | | | | |
|---|---|---|---|---|---|---|---|---|---|---|---|---|---|
| | Hypocalcemia [7][u] | 3[u] | - | - | - | - | 3[u] | 3[u] | 1[u] | - | - | - | 4[u] |
| | Hypoglycemia [2][u] | 2[u] | - | - | - | - | 2[u] | - | - | - | - | - | - |
| | Hypokalemia [19][u16] | 7[u5] | - | - | - | - | 7[u5] | 11[u10] | - | 1[u] | - | - | 12[u11] |
| | Hyponatremia [9][u] | 7[u] | - | - | - | - | 7[u] | 2[u] | - | - | - | - | 2[u] |
| | Obesity [2][u] | - | - | - | 1[u] | - | 1[u] | - | - | 1[u] | - | - | 1[u] |
| Musculo-skeletal and Connective Tissue Disorders [68][u18] | Back Pain [5][u4] | 2[u] | - | - | - | - | 2[u] | 1[u] | 2[u1] | - | - | - | 3[u2] |
| | Bone Pain [2][u] | - | - | - | - | - | - | 1[u] | 1[u] | - | - | - | 2[u] |
| | Joint Range of Motion Decreased [1][u] | 1[u] | - | - | - | - | 1[u] | - | - | - | - | - | - |
| | Muscle Weakness Upper Limb [1] | - | - | - | - | - | - | 1 | - | - | - | - | 1 |
| | Myalgia [56][u9] | 40[u5] | 4[u2] | - | - | - | 44[u7] | 9 | 3[u2] | - | - | - | 12[u2] |
| | Osteoporosis [1][u] | - | - | - | - | - | - | 1[u] | - | - | - | - | 1[u] |
| | Pain in Extremity [1][u] | - | - | - | - | - | - | 1[u] | - | - | - | - | 1[u] |
| | Musculoskeletal and Connective Tissue Disorder - Other, Specify [1] | - | 1 | - | - | - | 1 | - | - | - | - | - | - |
| Nervous System Disorders [78][u30, 1 SAE] | Concentration Impairment [1][u] | - | 1[u] | - | - | - | 1[u] | - | - | - | - | - | - |
| | Dizziness [18][u7] | 6[u3] | 2[u1] | - | - | - | 8[u4] | 9[u2] | 1[u] | - | - | - | 10[u3] |
| | Headache [51][u16, 1 SAE] | 19[u6] | 6[u2] | 1[u, SAE] | - | - | 26[u9, 1 SAE] | 18[u3] | 7[u4] | - | - | - | 25[u7] |

| | | | | | | | | | | | | | |
|---|---|---|---|---|---|---|---|---|---|---|---|---|---|
| | Lethargy [1][u] | 1[u] | - | - | - | - | 1[u] | - | - | - | - | - | - |
| | Paresthesia [2][u1] | 1[u] | - | - | - | - | 1[u] | 1 | - | - | - | - | 1 |
| | Peripheral Sensory Neuropathy [1][u] | 1[u] | - | - | - | - | 1[u] | - | - | - | - | - | - |
| | Somnolence [2][u1] | 1[u] | - | - | - | - | 1[u] | 1 | - | - | - | - | 1 |
| | Syncope [1][u] | - | - | 1[u] | - | - | 1[u] | - | - | - | - | - | - |
| | Nervous System Disorders - Other, Specify [1][u] | - | 1[u] | - | - | - | 1[u] | - | - | - | - | - | - |
| Pregnancy, Puerperium and Perinatal Conditions [9][u, 2 SAE] | Fetal Death [2][u, SAE] | - | - | - | - | 1[u, SAE] | 1[u, SAE] | - | - | - | - | 1[u, SAE] | 1[u, SAE] |
| | Unintended Pregnancy [7][u] | - | - | 4[u] | - | - | 4[u] | - | - | 3[u] | - | - | 3[u] |
| Psychiatric Disorders [20][u, 1 SAE] | Anxiety [9][u] | 3[u] | 3[u] | - | - | - | 6[u] | 1[u] | 2[u] | - | - | - | 3[u] |
| | Depression [6][u] | - | 2[u] | 1[u] | - | - | 3[u] | 3[u] | - | - | - | - | 3[u] |
| | Psychiatric Disorders - Other, Specify [5][u, 1 SAE] | 2[u] | 2[u] | - | 1[u, SAE] | - | 5[u, 1 SAE] | - | - | - | - | - | - |
| Renal and Urinary Disorders [5][u] | Acute Kidney Injury [1][u] | - | 1[u] | - | - | - | 1[u] | - | - | - | - | - | - |
| | Hematuria [1][u] | - | - | - | - | - | - | 1[u] | - | - | - | - | 1[u] |
| | Urinary Urgency [2][u] | 1[u] | - | - | - | - | 1[u] | 1[u] | - | - | - | - | 1[u] |
| | Renal and Urinary Disorders - Other, Specify [1][u] | - | 1[u] | - | - | - | 1[u] | - | - | - | - | - | - |

| | | | | | | | | | | | | | |
|---|---|---|---|---|---|---|---|---|---|---|---|---|---|
| Reproductive System and Breast Disorders [31][u] | Breast Pain [3][u] | 1[u] | 1[u] | - | - | - | 2[u] | 1[u] | - | - | - | - | 1[u] |
| | Dysmenorrhea [4][u] | 3[u] | 1[u] | - | - | - | 4[u] | - | - | - | - | - | - |
| | Dyspareunia [4][u] | 2[u] | - | - | - | - | 2[u] | 2[u] | - | - | - | - | 2[u] |
| | Irregular Menstruation [2][u] | - | - | - | - | - | - | 2[u] | - | - | - | - | 2[u] |
| | Pelvic Pain [1][u] | - | 1[u] | - | - | - | 1[u] | - | - | - | - | - | - |
| | Vaginal Discharge [4][u] | 4[u] | - | - | - | - | 4[u] | - | - | - | - | - | - |
| | Vaginal Dryness [1][u] | 1[u] | - | - | - | - | 1[u] | - | - | - | - | - | - |
| | Vaginal Hemorrhage [5][u] | 2[u] | - | - | - | - | 2[u] | 3[u] | - | - | - | - | 3[u] |
| | Vaginal Inflammation [1][u] | 1[u] | - | - | - | - | 1[u] | - | - | - | - | - | - |
| | Vaginal Pain [1][u] | - | - | - | - | - | - | 1[u] | - | - | - | - | 1[u] |
| | Vaginismus [1][u] | 1[u] | - | - | - | - | 1[u] | - | - | - | - | - | - |
| | Reproductive System and Breast Disorders - Other, Specify [4][u] | - | 2[u] | - | - | - | 2[u] | 2[u] | - | - | - | - | 2[u] |
| Respiratory, Thoracic and Mediastinal Disorders [24][u] | Allergic Rhinitis [2][u] | - | - | - | - | - | - | 1[u] | 1[u] | - | - | - | 2[u] |
| | Cough [3][u] | 2[u] | - | - | - | - | 2[u] | 1[u] | - | - | - | - | 1[u] |
| | Dyspnea [6][u] | - | - | 1[u] | - | - | 1[u] | 3[u] | 1[u] | 1[u] | - | - | 5[u] |
| | Nasal Congestion [5][u] | 3[u] | 1[u] | - | - | - | 4[u] | 1[u] | - | - | - | - | 1[u] |
| | Sore Throat [2][u] | - | - | - | - | - | - | 2[u] | - | - | - | - | 2[u] |
| | Wheezing [6][u] | 4[u] | 2[u] | - | - | - | 6[u] | - | - | - | - | - | - |

| | | | | | | | | | | | | | |
|---|---|---|---|---|---|---|---|---|---|---|---|---|---|
| Skin and Subcutaneous Tissue Disorders [18]$^{u15}$ | Alopecia [3]$^{u}$ | $2^{u}$ | - | - | - | - | $2^{u}$ | $1^{u}$ | - | - | - | - | $1^{u}$ |
| | Erythema Multiforme [1]$^{u}$ | - | - | - | - | - | - | $1^{u}$ | - | - | - | - | $1^{u}$ |
| | Hyperhidrosis [1]$^{u}$ | - | $1^{u}$ | - | - | - | $1^{u}$ | - | - | - | - | - | - |
| | Rash Acneiform [1] | - | - | - | - | - | - | 1 | - | - | - | - | 1 |
| | Rash Maculo-Papular [1]$^{u}$ | $1^{u}$ | - | - | - | - | $1^{u}$ | - | - | - | - | - | - |
| | Skin Ulceration [1]$^{u}$ | - | - | - | - | - | - | $1^{u}$ | - | - | - | - | $1^{u}$ |
| | Skin and Subcutaneous Tissue Disorders - Other, Specify [10]$^{u8}$ | $2^{u}$ | $2^{u}$ | - | - | - | $4^{u}$ | $3^{u1}$ | $3^{u}$ | - | - | - | $6^{u4}$ |
| Vascular Disorders [6]$^{u4}$ | Hot Flashes [3]$^{u2}$ | $1^{u}$ | - | - | - | - | $1^{u}$ | $2^{u1}$ | - | - | - | - | $2^{u1}$ |
| | Hypertension [2]$^{u}$ | - | - | $1^{u}$ | - | - | $1^{u}$ | - | - | $1^{u}$ | - | - | $1^{u}$ |
| | Hypotension [1] | - | - | - | - | - | - | - | 1 | - | - | - | 1 |

U = unrelated to treatment

The highest grade shown for each adverse event.

**TABLE S1B. All adverse events regardless of treatment-relatedness by patient.**

| All Adverse Events at the Patient Level by Treatment Group | | CTCAE v4.03 Adverse Event | | | |
|---|---|---|---|---|---|
| | | PepCan (n=39) | | *Candida* (n=42) | |
| CTCAE v4.03 System Organ Class | CTCAE v4.03 Adverse Event Term | Number of Patients with the Given AE | Total | Number of Patients with the Given AE | Total |
| Blood and Lymphatic System Disorders [17][u] | Anemia [17][u] | 9 | 9[u] | 7 | 8[u] |
| Cardiac Disorders [7][u] | Chest Pain - Cardiac [1][u] | - | - | 1 | 1[u] |
| | Palpitations [4][u] | 1 | 1[u] | 1 | 3[u] |
| | Sinus Bradycardia [1][u] | - | - | 1 | 1[u] |
| | Sinus Tachycardia [1][u] | 1 | 1[u] | - | - |
| Ear and Labyrinth Disorders [3][u] | Ear Pain [3][u] | 1 | 3[u] | - | - |
| Endocrine Disorders [3][u] | Hypothyroidism [3][u] | - | - | 3 | 3[u] |
| Eye Disorders [2][u] | Photophobia [1][u] | 1 | 1[u] | - | - |
| | Eye Disorders - Other, Specify [1][u] | - | - | 1 | 1[u] |
| Gastrointestinal Disorders [93][u53] | Abdominal Pain [8][u7] | 4 | 6[u5] | 2 | 2[u] |
| | Bloating [1][u] | - | - | 1 | 1[u] |
| | Colitis [1][u] | - | - | 1 | 1[u] |
| | Constipation [8][u] | 4 | 6[u] | 1 | 2[u] |
| | Diarrhea [2][u] | - | - | 2 | 2[u] |

| | | | | | |
|---|---|---|---|---|---|
| | Gastritis [1][u] | 1 | 1[u] | - | - |
| | Gastroesophageal Reflux Disease [1][u] | 1 | 1[u] | - | - |
| | Gastrointestinal Pain [2][u] | 1 | 1[u] | 1 | 1[u] |
| | Gastroparesis [1][u] | 1 | 1[u] | - | - |
| | Nausea [53][u16] | 15 | 31[u12] | 12 | 22[u4] |
| | Stomach Pain [3][u] | 1 | 1[u] | 1 | 2[u] |
| | Vomiting [4][u3] | 3 | 3[u2] | 1 | 1[u] |
| | Gastrointestinal Disorders - Other, Specify [8][u7] | 3 | 4[u] | 4 | 4[u3] |
| General Disorders and Site Administration Conditions [355][u33] | Edema Limbs [1][u] | - | - | 1 | 1[u] |
| | Fatigue [18][u8] | 8 | 9[u5] | 7 | 9[u3] |
| | Fever [19][u6] | 10 | 14[u4] | 4 | 5[u2] |
| | Flu-Like Symptoms [13][u3] | 6 | 8[u1] | 4 | 5[u2] |
| | Injection Site Reaction [288] | 34 | 142 | 38 | 146 |
| | Localized Edema [1][u] | - | - | 1 | 1[u] |
| | Malaise [1] | 1 | 1 | - | - |
| | Non-Cardiac Chest Pain [2][u] | - | - | 1 | 2[u] |
| | Pain [12][u] | 3 | 8[u] | 4 | 4[u] |
| Immune System Disorders [1][u] | Allergic Reaction [1][u] | 1 | 1[u] | - | - |
| Infections and | Appendicitis [1][u, SAE] | - | - | 1 | 1[u, SAE] |

| | | | | | |
|---|---|---|---|---|---|
| Infestations [47]u, 1 SAE | Laryngitis [1]u | 1 | 1u | - | - |
| | Pharyngitis [1]u | 1 | 1u | - | - |
| | Sinusitis [2]u | 2 | 2u | - | - |
| | Skin Infection [3]u | 1 | 2u | 1 | 1u |
| | Tooth Infection [1]u | 1 | 1u | - | - |
| | Upper Respiratory Infection [10]u | 6 | 9u | 1 | 1u |
| | Urinary Tract Infection [11]u | 5 | 7u | 4 | 4u |
| | Vaginal Infection [10]u | 4 | 5u | 5 | 5u |
| | Infections and Infestations - Other, Specify [7]u | 3 | 3u | 4 | 4u |
| Injury, Poisoning and Procedural Complications [6]u3 | Bruising [6]u3 | 4 | 4u2 | 2 | 2u1 |
| Investigations [60]u | Alanine Aminotransferase Increased [13]u | 8 | 8u | 5 | 5u |
| | Alkaline Phosphatase Increased [3]u | 2 | 2u | 1 | 1u |
| | Aspartate Aminotransferase Increased [14]u | 6 | 7u | 7 | 7u |

| | | | | | |
|---|---|---|---|---|---|
| | Blood Bilirubin Increased [3]$^{u}$ | 2 | 2$^{u}$ | 1 | 1$^{u}$ |
| | Creatinine Increased [9]$^{u}$ | 5 | 5$^{u}$ | 3 | 4$^{u}$ |
| | Hemoglobin Increased [6]$^{u}$ | 3 | 4$^{u}$ | 2 | 2$^{u}$ |
| | Platelet Count Decreased [3]$^{u}$ | 1 | 1$^{u}$ | 1 | 2$^{u}$ |
| | Weight Loss [1]$^{u}$ | 1 | 1$^{u}$ | - | - |
| | White Blood Cell Decreased [4]$^{u}$ | 1 | 2$^{u}$ | 2 | 2$^{u}$ |
| | Investigations - Other, Specify [4]$^{u}$ | 2 | 2$^{u}$ | 2 | 2$^{u}$ |
| Metabolism and Nutrition Disorders [57]$^{u54}$ | Dehydration [1]$^{u}$ | - | - | 1 | 1$^{u}$ |
| | Hyperglycemia [4]$^{u}$ | 3 | 3$^{u}$ | 1 | 1$^{u}$ |
| | Hyperkalemia [1]$^{u}$ | 1 | 1$^{u}$ | - | - |
| | Hypoalbuminemia [12]$^{u}$ | 3 | 3$^{u}$ | 9 | 9$^{u}$ |
| | Hypocalcemia [7]$^{u}$ | 3 | 3$^{u}$ | 4 | 4$^{u}$ |
| | Hypoglycemia [2]$^{u}$ | 2 | 2$^{u}$ | - | - |
| | Hypokalemia [19]$^{u16}$ | 7 | 7$^{u5}$ | 10 | 12$^{u11}$ |
| | Hyponatremia [9]$^{u}$ | 5 | 7$^{u}$ | 1 | 2$^{u}$ |
| | Obesity [2]$^{u}$ | 1 | 1$^{u}$ | 1 | 1$^{u}$ |
| Musculo-skeletal and Connective Tissue | Back Pain [5]$^{u4}$ | 2 | 2$^{u}$ | 3 | 3$^{u2}$ |
| | Bone Pain [2]$^{u}$ | - | - | 2 | 2$^{u}$ |

| | | | | | |
|---|---|---|---|---|---|
| Disorders [68]u18 | Joint Range of Motion Decreased [1]u | 1 | 1u | - | - |
| | Muscle Weakness Upper Limb [1] | - | - | 1 | 1 |
| | Myalgia [56]u9 | 12 | 44u7 | 9 | 12u2 |
| | Osteoporosis [1]u | - | - | 1 | 1u |
| | Pain in Extremity [1]u | - | - | 1 | 1u |
| | Musculoskeletal and Connective Tissue Disorder - Other, Specify [1] | 1 | 1 | - | - |
| Nervous System Disorders [78]u30, 1 SAE | Concentration Impairment [1]u | 1 | 1u | - | - |
| | Dizziness [18]u7 | 7 | 8u4 | 5 | 10u3 |
| | Headache [51]u16, 1 SAE | 11 | 26u9, 1 SAE | 12 | 25u7 |
| | Lethargy [1]u | 1 | 1u | - | - |
| | Paresthesia [2]u1 | 1 | 1u | 1 | 1 |
| | Peripheral Sensory Neuropathy [1]u | 1 | 1u | - | - |
| | Somnolence [2]u1 | 1 | 1u | 1 | 1 |
| | Syncope [1]u | 1 | 1u | - | - |
| | Nervous System Disorders - Other, Specify [1]u | 1 | 1u | - | - |

| | | | | | |
|---|---|---|---|---|---|
| Pregnancy, Puerperium and Perinatal Conditions [9]u, 2 SAE | Fetal Death [2]u, SAE | 1 | 1u, SAE | 1 | 1u, SAE |
| | Unintended Pregnancy [7]u | 4 | 4u | 3 | 3u |
| Psychiatric Disorders [20]u, 1 SAE | Anxiety [9]u | 4 | 6u | 3 | 3u |
| | Depression [6]u | 2 | 3u | 2 | 3u |
| | Psychiatric Disorders - Other, Specify [5]u, 1 SAE | 3 | 5u, 1 SAE | - | - |
| Renal and Urinary Disorders [5]u | Acute Kidney Injury [1]u | 1 | 1u | - | - |
| | Hematuria [1]u | - | - | 1 | 1u |
| | Urinary Urgency [2]u | 1 | 1u | 1 | 1u |
| | Renal and Urinary Disorders - Other, Specify [1]u | 1 | 1u | - | - |
| Reproductive System and Breast Disorders [31]u | Breast Pain [3]u | 2 | 2u | 1 | 1u |
| | Dysmenorrhea [4]u | 4 | 4u | - | - |
| | Dyspareunia [4]u | 2 | 2u | 2 | 2u |
| | Irregular Menstruation [2]u | - | - | 2 | 2u |
| | Pelvic Pain [1]u | 1 | 1u | - | - |
| | Vaginal Discharge [4]u | 4 | 4u | - | - |
| | Vaginal Dryness [1]u | 1 | 1u | - | - |
| | Vaginal Hemorrhage [5]u | 2 | 2u | 3 | 3u |

| | | | | | |
|---|---|---|---|---|---|
| | Vaginal Inflammation [1][u] | 1 | 1[u] | - | - |
| | Vaginal Pain [1][u] | - | - | 1 | 1[u] |
| | Vaginismus [1][u] | 1 | 1[u] | - | - |
| | Reproductive System and Breast Disorders - Other, Specify [4][u] | 2 | 2[u] | 2 | 2[u] |
| Respiratory, Thoracic and Mediastinal Disorders [24][u] | Allergic Rhinitis [2][u] | - | - | 2 | 2[u] |
| | Cough [3][u] | 2 | 2[u] | 1 | 1[u] |
| | Dyspnea [6][u] | 1 | 1[u] | 2 | 5[u] |
| | Nasal Congestion [5][u] | 2 | 4[u] | 1 | 1[u] |
| | Sore Throat [2][u] | - | - | 2 | 2[u] |
| | Wheezing [6][u] | 4 | 6[u] | - | - |
| Skin and Subcutaneous Tissue Disorders [18][u15] | Alopecia [3][u] | 2 | 2[u] | 1 | 1[u] |
| | Erythema Multiforme [1][u] | - | - | 1 | 1[u] |
| | Hyperhidrosis [1][u] | 1 | 1[u] | - | - |
| | Rash Acneiform [1] | - | - | 1 | 1 |
| | Rash Maculo-Papular [1][u] | 1 | 1[u] | - | - |
| | Skin Ulceration [1][u] | - | - | 1 | 1[u] |
| | Skin and Subcutaneous Tissue Disorders - Other, | 4 | 4[u] | 6 | 6[u4] |

| | | | | | |
|---|---|---|---|---|---|
| | Specify [10][u8] | | | | |
| Vascular Disorders [6][u4] | Hot Flashes [3][u2] | 1 | 1[u] | 1 | 2[u1] |
| | Hypertension [2][u] | 1 | 1[u] | 1 | 1[u] |
| | Hypotension [1] | - | - | 1 | 1 |

u=unrelated to treatment

**TABLE S2A. Entry cytokine levels by histological response groups (stringent) for patients who completed the 6-month visit in both treatment groups.**

| Cytokine | No Response (N=44) | Response (N=32) | *P*-value* | |
|---|---|---|---|---|
| | | | Unadjusted | Adjusted^ |
| TGFb1 | 4346.8 (3446.0, 6572.5) | 4450.7 (3058.6, 5867.4) | 0.68 | 0.86 |
| TGFb2 | 329.1 (275.5, 1809.5) | 1366.4 (331.5, 2848.3) | 0.09 | 0.49 |
| TGFb3 | 23.0 (14.5, 72.4) | 53.6 (23.0, 117.6) | 0.06 | 0.41 |
| CTACK | 506.3 (398.7, 646.0) | 435.5 (338.1, 682.8) | 0.37 | 0.61 |
| Eotaxin | 90.6 (60.9, 128.7) | 69.9 (50.1, 96.7) | 0.18 | 0.49 |
| FGF | 19.9 (10.9, 28.0) | 19.8 (11.3, 31.6) | 0.90 | 0.96 |
| G-CSF | 42.6 (14.4, 49.4) | 38.0 (24.9, 67.9) | 0.38 | 0.61 |
| GM-CSF | 0.6 (0.0, 1.3) | 0.0 (0.0, 0.7) | **0.007** | 0.24 |
| GROa | 191.3 (92.7, 397.1) | 226.3 (127.7, 942.7) | 0.17 | 0.49 |
| HGF | 251.1 (210.5, 332.5) | 246.6 (198.7, 326.8) | 0.71 | 0.86 |
| IFNg | 2.4 (0.0, 6.7) | 4.1 (0.0, 6.8) | 1.00 | 1.00 |
| IL01a | 6.0 (2.0, 7.6) | 3.5 (1.8, 6.2) | 0.36 | 0.61 |
| IL01b | 1.8 (1.3, 2.5) | 2.0 (1.4, 4.1) | 0.38 | 0.61 |
| IL01ra | 191.5 (112.6, 264.7) | 240.1 (114.1, 397.7) | 0.29 | 0.61 |
| IL02ra | 67.3 (54.5, 92.4) | 58.6 (48.6, 74.7) | 0.18 | 0.49 |
| IL04 | 2.6 (0.9, 3.9) | 2.1 (1.1, 2.9) | 0.34 | 0.61 |
| IL06 | 1.2 (0.0, 3.1) | 0.0 (0.0, 1.7) | 0.10 | 0.49 |
| IL08 | 7.5 (5.8, 9.7) | 8.1 (6.4, 9.4) | 0.72 | 0.86 |
| IL09 | 170.9 (68.6, 288.6) | 295.3 (154.2, 434.1) | 0.02 | 0.26 |
| IL10 | 2.1 (0.0, 4.0) | 1.0 (0.0, 3.0) | 0.38 | 0.61 |
| IL12p70 | 1.9 (0.0, 3.4) | 1.0 (0.0, 2.6) | 0.15 | 0.49 |
| IL12p40 | 22.6 (6.3, 27.7) | 25.8 (11.1, 38.3) | 0.14 | 0.49 |
| IL13 | 0.5 (0.0, 0.9) | 0.9 (0.4, 1.6) | **0.02** | 0.24 |
| IL16 | 87.4 (76.8, 110.2) | 96.8 (78.1, 128.7) | 0.30 | 0.61 |
| IL17 | 3.4 (0.0, 6.0) | 2.9 (0.0, 9.1) | 0.61 | 0.80 |
| IL18 | 37.0 (28.8, 69.0) | 37.7 (28.9, 61.1) | 0.83 | 0.94 |
| IP10 | 592.0 (344.2, 939.7) | 577.5 (279.1, 868.0) | 0.37 | 0.61 |
| LIF | 11.6 (0.0, 45.4) | 24.1 (8.5, 47.9) | 0.14 | 0.49 |
| M-CSF | 23.9 (19.6, 31.8) | 22.4 (17.0, 31.5) | 0.47 | 0.66 |
| MCP01 | 44.5 (33.1, 56.8) | 40.7 (33.0, 52.4) | 0.48 | 0.66 |
| MCP03 | 0.2 (0.0, 1.9) | 0.4 (0.0, 1.8) | 0.58 | 0.78 |
| MIF | 1838.9 (1384.8, 2448.3) | 1732.1 (1216.1, 3045.4) | 0.98 | 1.00 |
| MIG | 273.2 (215.1, 484.7) | 319.8 (224.9, 466.5) | 0.92 | 0.96 |
| MIP01a | 0.7 (0.5, 1.0) | 0.9 (0.6, 1.2) | 0.45 | 0.66 |
| MIP01b | 93.7 (41.1, 131.6) | 136.7 (71.0, 198.1) | **0.02** | 0.24 |

| | | | | |
|---|---|---|---|---|
| PDGFbb | 428.4 (278.9, 654.1) | 430.6 (215.5, 707.2) | 0.86 | 0.95 |
| RANTES | 2994.6 (1967.3, 4742.7) | 3348.5 (2369.9, 4694.2) | 0.46 | 0.66 |
| SCF | 59.1 (33.8, 93.9) | 83.4 (34.9, 109.9) | 0.30 | 0.61 |
| SCGFb | 46162.8 (35531.8, 61272.2) | 53651.9 (39641.4, 84447.7) | 0.16 | 0.49 |
| SDF01a | 1753.8 (1502.0, 2492.1) | 2200.6 (1634.6, 3066.1) | 0.11 | 0.49 |
| TNFa | 45.5 (30.2, 64.9) | 45.0 (28.7, 65.4) | 0.78 | 0.91 |
| TNFb | 160.0 (0.0, 260.9) | 352.2 (75.3, 549.8) | **0.04** | 0.32 |
| TRAIL | 101.2 (75.3, 136.6) | 93.6 (31.8, 134.7) | 0.28 | 0.61 |

*Wilcoxon rank-sum test; ^Benjamini and Hochberg method

**TABLE S2B. Entry cytokine levels by histological response groups (stringent) for patients who completed the 6-month visit in the PepCan group.**

| Cytokine | No Response (N=23) | Response (N=12) | *P*-value* | |
|---|---|---|---|---|
| | | | Unadjusted | Adjusted^ |
| TGFb1 | 4199.5 (3333.1, 6169.9) | 3933.6 (3006.6, 5131.4) | 0.69 | 0.94 |
| TGFb2 | 325.6 (239.0, 2153.2) | 1996.1 (993.2, 2468.6) | 0.18 | 0.41 |
| TGFb3 | 23.2 (12.4, 89.8) | 69.8 (31.8, 108.1) | 0.14 | 0.40 |
| CTACK | 552.1 (422.5, 648.2) | 412.7 (282.9, 474.5) | 0.09 | 0.40 |
| Eotaxin | 97.3 (62.3, 122.3) | 65.4 (49.3, 90.7) | 0.15 | 0.40 |
| FGF | 18.8 (12.7, 28.2) | 14.1 (5.0, 27.8) | 0.34 | 0.66 |
| G-CSF | 40.1 (19.8, 48.0) | 31.7 (24.9, 54.9) | 0.85 | 0.94 |
| GM-CSF | 0.1 (0.0, 0.7) | 0.0 (0.0, 0.0) | 0.07 | 0.40 |
| GROa | 169.3 (106.1, 305.4) | 510.2 (114.1, 1177.4) | 0.31 | 0.62 |
| HGF | 246.8 (221.4, 288.9) | 246.3 (181.6, 310.5) | 0.74 | 0.94 |
| IFNg | 3.8 (0.4, 7.0) | 0.8 (0.0, 5.1) | 0.17 | 0.41 |
| IL01a | 3.2 (0.4, 7.4) | 4.1 (1.4, 5.1) | 0.94 | 0.97 |
| IL01b | 2.1 (1.6, 2.8) | 2.0 (1.2, 4.3) | 0.88 | 0.94 |
| IL01ra | 218.6 (110.7, 314.9) | 240.1 (174.6, 306.6) | 0.65 | 0.94 |
| IL02ra | 68.3 (61.5, 87.8) | 52.1 (47.6, 70.3) | 0.10 | 0.40 |
| IL04 | 2.5 (1.2, 3.8) | 1.6 (0.7, 2.5) | 0.12 | 0.40 |
| IL06 | 0.8 (0.0, 2.0) | 0.0 (0.0, 0.3) | 0.10 | 0.40 |
| IL08 | 7.0 (5.3, 9.5) | 7.1 (4.2, 8.9) | 0.72 | 0.94 |
| IL09 | 155.7 (68.3, 253.2) | 337.0 (174.0, 413.3) | 0.05 | 0.40 |
| IL10 | 2.3 (0.0, 4.2) | 1.1 (0.0, 2.6) | 0.15 | 0.40 |
| IL12p70 | 1.5 (0.0, 2.7) | 0.0 (0.0, 1.2) | 0.05 | 0.40 |
| IL12p40 | 21.6 (6.4, 27.5) | 28.6 (7.5, 42.0) | 0.41 | 0.74 |
| IL13 | 0.4 (0.0, 0.7) | 0.9 (0.5, 1.6) | 0.10 | 0.40 |
| IL16 | 88.6 (81.6, 121.1) | 98.4 (79.5, 121.3) | 0.74 | 0.94 |
| IL17 | 1.5 (0.0, 6.2) | 3.1 (0.0, 5.8) | 0.55 | 0.92 |
| IL18 | 38.1 (29.5, 77.0) | 43.9 (31.5, 54.2) | 0.99 | 0.99 |
| IP10 | 601.0 (463.9, 898.8) | 417.9 (203.8, 791.7) | 0.09 | 0.40 |
| LIF | 11.6 (0.3, 55.5) | 20.8 (5.7, 35.6) | 0.75 | 0.94 |
| M-CSF | 24.2 (19.6, 32.6) | 19.1 (10.5, 23.7) | 0.13 | 0.40 |
| MCP01 | 47.1 (34.9, 57.4) | 38.7 (34.6, 44.1) | 0.16 | 0.40 |
| MCP03 | 0.0 (0.0, 1.9) | 0.2 (0.0, 1.1) | 0.84 | 0.94 |
| MIF | 1853.7 (1294.2, 2935.4) | 1817.7 (1216.1, 2437.5) | 0.90 | 0.95 |
| MIG | 265.7 (213.9, 543.4) | 208.9 (145.0, 265.0) | 0.06 | 0.40 |
| MIP01a | 0.8 (0.6, 1.1) | 0.8 (0.6, 1.0) | 0.81 | 0.94 |
| MIP01b | 85.0 (40.6, 124.6) | 147.4 (91.1, 231.3) | **0.02** | 0.40 |

| | | | | |
|---|---|---|---|---|
| PDGFbb | 448.1 (294.0, 643.4) | 335.9 (192.1, 490.5) | 0.36 | 0.67 |
| RANTES | 2258.2 (1859.7, 3852.7) | 3426.0 (2053.2, 5661.9) | 0.52 | 0.89 |
| SCF | 60.2 (33.7, 91.5) | 73.8 (41.9, 109.5) | 0.61 | 0.94 |
| SCGFb | 49314.6 (38359.5, 70213.2) | 45926.2 (36638.7, 72393.3) | 0.79 | 0.94 |
| SDF01a | 1635.3 (1505.6, 2474.4) | 2154.2 (1664.6, 2867.8) | 0.29 | 0.62 |
| TNFa | 45.2 (33.1, 63.1) | 41.2 (21.8, 58.9) | 0.79 | 0.94 |
| TNFb | 125.6 (0.0, 247.9) | 484.1 (140.5, 566.2) | **0.04** | 0.40 |
| TRAIL | 100.6 (74.3, 139.9) | 92.4 (31.2, 142.6) | 0.79 | 0.94 |

*Wilcoxon rank-sum test; ^Benjamini and Hochberg method

**TABLE S2C. Entry cytokine levels by histological response groups (stringent) for patients who completed the 6-month visit in the *Candida* group.**

| Cytokine | No Response (N=21) | Response (N=20) | *P*-value* | |
|---|---|---|---|---|
| | | | Unadjusted | Adjusted^ |
| TGFb1 | 5043.3 (3752.6, 6782.9) | 5196.3 (3341.4, 5949.0) | 0.78 | 0.93 |
| TGFb2 | 332.6 (308.5, 1745.8) | 856.1 (323.5, 3146.1) | 0.35 | 0.66 |
| TGFb3 | 22.8 (18.0, 66.1) | 42.1 (20.9, 129.4) | 0.28 | 0.66 |
| CTACK | 464.2 (375.5, 612.5) | 476.3 (362.8, 769.2) | 0.97 | 1.00 |
| Eotaxin | 87.9 (60.8, 138.4) | 75.4 (59.5, 115.1) | 0.56 | 0.89 |
| FGF | 19.9 (10.9, 27.8) | 20.4 (15.1, 40.7) | 0.33 | 0.66 |
| G-CSF | 43.4 (9.8, 51.4) | 40.4 (25.0, 74.7) | 0.42 | 0.75 |
| GM-CSF | 1.1 (0.3, 2.1) | 0.0 (0.0, 0.9) | **0.01** | 0.53 |
| GROa | 197.3 (87.4, 498.1) | 168.2 (130.2, 662.4) | 0.46 | 0.76 |
| HGF | 264.3 (171.9, 368.7) | 252.4 (203.3, 326.8) | 0.79 | 0.93 |
| IFNg | 0.0 (0.0, 6.3) | 5.5 (0.0, 8.2) | 0.26 | 0.66 |
| IL01a | 6.2 (4.1, 7.7) | 3.3 (2.0, 6.6) | 0.13 | 0.66 |
| IL01b | 1.6 (1.2, 2.4) | 2.0 (1.4, 3.9) | 0.17 | 0.66 |
| IL01ra | 170.9 (115.3, 249.1) | 241.4 (110.3, 420.1) | 0.33 | 0.66 |
| IL02ra | 64.7 (53.4, 100.3) | 63.0 (54.7, 77.5) | 0.80 | 0.93 |
| IL04 | 2.7 (0.0, 3.8) | 2.4 (1.7, 3.1) | 0.95 | 1.00 |
| IL06 | 1.6 (0.0, 4.9) | 0.9 (0.0, 1.9) | 0.23 | 0.66 |
| IL08 | 7.8 (6.4, 10.8) | 8.4 (6.9, 10.1) | 0.63 | 0.90 |
| IL09 | 192.2 (78.0, 359.0) | 244.2 (140.3, 434.1) | 0.25 | 0.66 |
| IL10 | 1.3 (0.0, 3.7) | 1.0 (0.0, 5.0) | 1.00 | 1.00 |
| IL12p70 | 3.0 (0.0, 4.1) | 1.8 (0.3, 2.7) | 0.32 | 0.66 |
| IL12p40 | 23.7 (6.9, 27.7) | 25.8 (14.4, 37.5) | 0.23 | 0.66 |
| IL13 | 0.5 (0.0, 0.9) | 0.9 (0.4, 1.6) | 0.10 | 0.66 |
| IL16 | 83.2 (75.8, 108.2) | 91.4 (77.2, 131.2) | 0.22 | 0.66 |
| IL17 | 4.4 (0.0, 5.6) | 1.9 (0.0, 9.9) | 0.79 | 0.93 |
| IL18 | 36.1 (28.9, 63.0) | 36.0 (27.3, 80.0) | 0.71 | 0.93 |
| IP10 | 521.6 (291.7, 1055.3) | 681.0 (432.1, 868.0) | 0.84 | 0.95 |
| LIF | 8.4 (0.0, 30.2) | 25.5 (10.6, 98.4) | 0.12 | 0.66 |
| M-CSF | 23.6 (19.6, 31.5) | 26.7 (19.0, 34.2) | 0.77 | 0.93 |
| MCP01 | 42.5 (31.0, 55.1) | 43.2 (30.9, 53.4) | 0.87 | 0.95 |
| MCP03 | 0.5 (0.0, 2.0) | 0.7 (0.0, 3.4) | 0.59 | 0.90 |
| MIF | 1715.5 (1395.7, 2276.9) | 1554.1 (1198.7, 3324.1) | 0.69 | 0.93 |
| MIG | 286.8 (235.7, 387.1) | 358.8 (301.2, 472.7) | 0.07 | 0.66 |
| MIP01a | 0.7 (0.5, 0.9) | 0.9 (0.6, 1.3) | 0.29 | 0.66 |
| MIP01b | 107.0 (42.6, 156.5) | 129.0 (62.3, 176.3) | 0.33 | 0.66 |
| PDGFbb | 409.9 (244.4, 640.2) | 488.6 (288.4, 807.9) | 0.35 | 0.66 |

| | | | | |
|---|---|---|---|---|
| RANTES | 3089.1 (2286.0, 4932.7) | 3209.5 (2494.7, 4570.5) | 0.99 | 1.00 |
| SCF | 51.8 (34.2, 96.6) | 84.9 (34.8, 117.0) | 0.35 | 0.66 |
| SCGFb | 40792.1 (32968.8, 54366.2) | 54553.5 (46306.1, 90334.5) | **0.04** | 0.66 |
| SDF01a | 1811.5 (1505.7, 2488.0) | 2244.6 (1591.7, 3635.4) | 0.19 | 0.66 |
| TNFa | 45.9 (20.6, 68.9) | 51.0 (32.8, 65.4) | 0.63 | 0.90 |
| TNFb | 173.0 (0.0, 389.6) | 314.8 (67.3, 460.1) | 0.45 | 0.76 |
| TRAIL | 101.8 (79.0, 133.3) | 93.6 (37.6, 113.9) | 0.26 | 0.66 |

*Wilcoxon rank-sum test; ^Benjamini and Hochberg method

#### TABLE S3A. Entry metabolite levels~ by histological response groups (stringent) for patients who completed the 6-month visit in both treatment groups.

| Metabolite | Non-Responders (N=44) | Responders (N=32) | Log2Flod changes Response/No Response | P-value* | |
|---|---|---|---|---|---|
| | | | | Unadjusted | Adjusted^ |
| 4-hydroxyhippurate | 1.7406(0.25374,2.0056) | 0.90291(0.26411,3.616) | -0.94697 | **<0.001** | 0.49 |
| succinoyltaurine | 0.8343(0.20521,15.292) | 1.2809(0.38772,13.604) | 0.61847 | **0.003** | 1 |
| 3-acetylphenol sulfate | 2.141(0.090435,2.273) | 1.1901(0.090435,1.8475) | -0.84726 | **0.003** | 1 |
| leukotriene B5 | 0.91962(0.21937,8.0027) | 0.58526(0.21937,5.6794) | -0.65198 | **0.004** | 1 |
| 1-eicosenoyl-GPE (20:1)* | 1.4158(0.088073,2.3575) | 2.0961(0.40567,0.63139) | 0.56615 | **0.005** | 1 |
| X-17692 | 0.43164(0.14419,16.496) | 0.15941(0.14419,13.664) | -1.4371 | **0.005** | 1 |
| 5-hydroxy-2-methylpyridine sulfate | 4.0765(0.020551,3.1963) | 1.4689(0.020551,1.2364) | -1.4725 | **0.007** | 1 |
| 3-(4-hydroxyphenyl)propionate | 0.76362(0.20896,2.531) | 0.35636(0.20896,2.0814) | -1.0995 | **0.007** | 1 |
| X-12707 | 1.2278(0.25278,28.603) | 0.84302(0.25278,12.203) | -0.54239 | **0.01** | 1 |
| glucuronide of C10H18O2 (7)* | 2.5067(0.10615,7.6889) | 1.1666(0.10615,5.3618) | -1.1034 | **0.01** | 1 |
| X-13695 | 1.5456(0.13863,3.5854) | 0.79192(0.13863,8.2192) | -0.96471 | **0.01** | 1 |
| cinnamoylglycine | 0.85528(0.079964,19.446) | 1.6723(0.079964,9.1314) | 0.96737 | **0.01** | 1 |
| gentisate | 2.0963(0.25039,2.4853) | 1.028(0.25039,1.646) | -1.028 | **0.01** | 1 |
| 1-palmitoyl-2-alpha-linolenoyl-GPC (16:0/18:3n3)* | 0.67932(0.31754,2.4475) | 0.47295(0.31754,12.042) | -0.52238 | **0.01** | 1 |
| X-21815 | 0.37526(0.14339,6.3913) | 1.6165(0.14339,2.3239) | 2.1069 | **0.01** | 1 |
| feruloylquinate (1) | 0.76616(0.19266,12.209) | 0.28141(0.19266,1.3734) | -1.445 | **0.02** | 1 |
| N,N-dimethyl-5-aminovalerate | 1.3298(0.15991,2.3037) | 0.41546(0.15991,2.759) | -1.6784 | **0.02** | 1 |
| 5-hydroxymethyl-2-furoic acid | 0.68867(0.34454,6.3864) | 0.51447(0.34454,3.3157) | -0.42073 | **0.02** | 1 |
| X-12221 | 1.5831(0.10566,3.5081) | 0.79207(0.10566,3.802) | -0.99907 | **0.02** | 1 |
| 1-palmitoyl-2-docosahexaenoyl-GPE (16:0/22:6)* | 0.97735(0.27287,3.7336) | 1.4384(0.27287,4.5729) | 0.55752 | **0.02** | 1 |
| 3-phenylpropionate (hydrocinnamate) | 0.98558(0.065799,37.119) | 1.4845(0.065799,12.427) | 0.59091 | **0.02** | 1 |
| X-12730 | 3.3998(0.05611,32.898) | 1.2526(0.05611,11.801) | -1.4405 | **0.02** | 1 |
| 4-vinylguaiacol sulfate | 2.7388(0.052813,5.668) | 1.0739(0.052813,2.8789) | -1.3507 | **0.02** | 1 |

| | | | | | |
|---|---|---|---|---|---|
| 2-methoxyhydroquinone sulfate (2) | 1.1984(0.18937,58.206) | 0.58849(0.18937,20.21) | -1.0261 | **0.02** | 1 |
| citraconate/glutaconate | 7.375(0.17734,7.3612) | 2.6648(0.17707,5.2333) | -1.4686 | **0.02** | 1 |
| 4-ethyl-2-methoxyphenol sulfate | 0.82856(0.092197,4.22) | 0.42258(0.092197,2.5869) | -0.97136 | **0.02** | 1 |
| X-13007 | 1.4388(0.16538,4.1929) | 0.98886(0.16538,3.7876) | -0.54099 | **0.02** | 1 |
| sulfate of piperine metabolite C16H19NO3 (2)* | 1.3691(0.035245,2.3477) | 0.90736(0.040137,1.3095) | -0.59344 | **0.02** | 1 |
| X-13553 | 1.071(0.20384,3.9639) | 0.82031(0.20384,3.6948) | -0.38468 | **0.02** | 1 |
| sulfate of piperine metabolite C16H19NO3 (3)* | 1.4501(0.030699,5.0202) | 0.93535(0.05211,2.5353) | -0.63254 | **0.02** | 1 |
| 1-pentadecanoyl-2-docosahexaenoyl-GPC (15:0/22:6)* | 1.0462(0.28868,11.454) | 1.1927(0.61265,2.2376) | 0.1891 | **0.02** | 1 |
| X-12729 | 1.5857(0.15439,2.6731) | 0.88314(0.15439,1.6465) | -0.84443 | **0.02** | 1 |
| 1-pentadecanoyl-2-arachidonoyl-GPC (15:0/20:4)* | 0.95656(0.26954,2.5482) | 1.095(0.6239,2.6881) | 0.19497 | **0.02** | 1 |
| cysteine-glutathione disulfide | 0.90289(0.22017,157.12) | 1.1519(0.22017,56.932) | 0.35146 | **0.03** | 1 |
| N-(2-furoyl)glycine | 8.5333(0.11543,18.478) | 3.2669(0.11543,26.368) | -1.3852 | **0.03** | 1 |
| X-17348 | 1.2522(0.1801,2.8829) | 2.3582(0.1801,0.034695) | 0.9132 | **0.03** | 1 |
| X-25468 | 0.16028(0.034695,2.0066) | 0.034695(0.034695,2.0502) | -2.2078 | **0.03** | 1 |
| 1-palmitoleoyl-2-linoleoyl-GPC (16:1/18:2)* | 0.71811(0.22666,22.179) | 0.99425(0.22666,14.318) | 0.4694 | **0.03** | 1 |
| X-23655 | 1.7915(0.041948,3.0028) | 0.83356(0.041948,12.03) | -1.1038 | **0.03** | 1 |
| X-25450 | 0.41626(0.30623,3.7284) | 0.86233(0.30623,2.8858) | 1.0507 | **0.03** | 1 |
| 1-oleoyl-2-arachidonoyl-GPE (18:1/20:4)* | 0.99726(0.28073,2.6928) | 1.2522(0.41461,2.2995) | 0.32843 | **0.03** | 1 |
| X-25790 | 1.0323(0.27542,52.073) | 1.2537(0.43322,30.12) | 0.28024 | **0.03** | 1 |
| 3-hydroxypyridine sulfate | 7.1997(0.039269,11.846) | 2.9146(0.028775,11.251) | -1.3046 | **0.04** | 1 |
| X-13846 | 1.4465(0.15065,2.1092) | 0.95563(0.15065,2.1802) | -0.59799 | **0.04** | 1 |
| gamma-tocopherol/beta-tocopherol | 1.1068(0.35884,1.9405) | 0.93946(0.32908,9.4172) | -0.23652 | **0.04** | 1 |
| 1-dihomo-linolenylglycerol (20:3) | 0.90461(0.3116,23.24) | 1.4678(0.31489,2.8999) | 0.69831 | **0.04** | 1 |
| 12,13-DiHOME | 1.6475(0.16318,2.2363) | 1.2066(0.32528,2.4323) | -0.44936 | **0.04** | 1 |
| X-24309 | 1.0124(0.40024,9.5089) | 1.2319(0.35333,3.696) | 0.28309 | **0.04** | 1 |
| glucuronide of C10H18O2 (8)* | 0.96235(0.18425,559.88) | 0.48559(0.18425,1.8023) | -0.98682 | **0.04** | 1 |
| saccharin | 15.373(0.16952,10.402) | 0.40571(0.16952,2.3269) | -5.2438 | **0.04** | 1 |

| | | | | | |
|---|---|---|---|---|---|
| maleate | 1.8663(0.44542,8.1817) | 0.95914(0.44542,12.641) | -0.96034 | **0.04** | 1 |
| 1,6-anhydroglucose | 1.3928(0.035287,75.756) | 1.4052(0.035287,12.7) | 0.012765 | **0.04** | 1 |
| X-18901 | 2.7461(0.15103,6.0871) | 2.3217(0.15103,1.9629) | -0.24221 | **0.04** | 1 |
| indoleacetate | 1.3158(0.40687,2.7355) | 0.99832(0.50362,5.9158) | -0.39832 | **0.04** | 1 |
| 1-arachidonylglycerol (20:4) | 1.0061(0.3772,12.765) | 1.2842(0.46995,4.6573) | 0.35206 | **0.04** | 1 |
| X-24541 | 1.1027(0.2287,1.7734) | 0.51059(0.2287,2.1508) | -1.1108 | **0.04** | 1 |
| X-11632 | 0.83936(0.30637,3.5308) | 1.0203(0.30637,2.2399) | 0.28164 | **0.04** | 1 |
| N-succinyl-phenylalanine | 1.0378(0.25307,2.7615) | 0.76609(0.25307,2.1666) | -0.43795 | **0.04** | 1 |
| X-24949 | 0.95043(0.18361,4.7923) | 1.1693(0.18361,7.4194) | 0.29894 | **0.05** | 1 |
| X-12112 | 1.2393(0.31591,2.7631) | 1.6957(0.19933,3.7611) | 0.45233 | **0.05** | 1 |
| androsterone sulfate | 0.98851(0.21876,1.8472) | 1.3636(0.1729,2.0689) | 0.46414 | **0.05** | 1 |
| mead acid (20:3n9) | 0.44157(0.32863,22.047) | 0.62004(0.32863,3.9052) | 0.48971 | **0.05** | 1 |

~Sixty-two of 1,454 metabolites detected with unadjusted *p*-values of less than 0.05 before rounding are shown.
*Wilcoxon rank-sum test; ^Benjamini and Hochberg method

**TABLE S3B. Entry metabolite levels~ by histological response groups (stringent) for patients who completed the 6-month visit in the PepCan group.**

| Metabolite | Non-Responders (N=23) | Responders (N=12) | Log2Flod changes Response/No Response | P-value* | |
|---|---|---|---|---|---|
| | | | | Unadjusted | Adjusted^ |
| X-13553 | 1.1546(0.48063,2.3477) | 0.73268(0.20384,1.1999) | -0.65609 | **0.002** | 0.97 |
| glycoursodeoxycholic acid sulfate (1) | 1.7181(0.10076,6.3587) | 0.55575(0.10076,1.7905) | -1.6283 | **0.005** | 0.97 |
| X-24728 | 1.1745(0.45708,2.7264) | 0.65009(0.27437,1.1453) | -0.85333 | **0.006** | 0.97 |
| 3-(4-hydroxyphenyl)lactate (HPLA) | 1.0991(0.66934,1.4656) | 0.93848(0.62915,2.0023) | -0.22791 | **0.007** | 0.97 |
| 4-hydroxyhippurate | 1.7516(0.25374,7.2148) | 0.86431(0.26411,3.9462) | -1.019 | **0.007** | 0.97 |
| X-22162 | 0.89658(0.51549,1.1784) | 1.1467(0.81264,1.7312) | 0.355 | **0.007** | 0.97 |
| picolinate | 1.5231(0.24217,3.614) | 0.77739(0.24217,2.0059) | -0.97031 | **0.009** | 0.97 |
| alpha-CEHC | 0.66918(0.28975,6.6058) | 0.79669(0.28975,1.5188) | 0.25164 | **0.01** | 0.97 |
| 3-acetylphenol sulfate | 2.0831(0.090435,15.292) | 0.70773(0.090435,4.0116) | -1.5574 | **0.01** | 0.97 |
| N,N-dimethyl-5-aminovalerate | 2.0544(0.15991,12.209) | 0.46717(0.15991,0.9082) | -2.1367 | **0.01** | 0.97 |
| 1-palmitoleoyl-GPC (16:1)* | 0.91676(0.43029,1.3756) | 1.2742(0.44536,1.8725) | 0.47502 | **0.01** | 0.97 |
| X-12216 | 1.8772(0.19876,5.8947) | 0.87387(0.10924,2.5828) | -1.1031 | **0.01** | 0.97 |
| X-24585 | 1.0888(0.43248,2.0733) | 0.94565(0.41741,3.7883) | -0.20331 | **0.02** | 0.97 |
| 1-pentadecanoyl-GPC (15:0)* | 0.9151(0.44616,1.5625) | 1.1849(0.79315,1.8104) | 0.37279 | **0.02** | 0.97 |
| 2-methoxyhydroquinone sulfate (2) | 1.2289(0.18937,5.1797) | 0.36297(0.18937,0.95779) | -1.7595 | **0.02** | 0.97 |
| 3-(4-hydroxyphenyl)propionate | 0.64408(0.20896,2.303) | 0.26247(0.20896,0.85099) | -1.2951 | **0.02** | 0.97 |
| X-25450 | 0.32076(0.30623,0.6405) | 0.42343(0.30623,0.8081) | 0.40062 | **0.02** | 0.97 |
| stearoyl sphingomyelin (d18:1/18:0) | 1.0638(0.71825,1.6295) | 0.89923(0.63537,1.2276) | -0.24246 | **0.02** | 0.97 |
| 5alpha-androstan-3alpha,17beta-diol monosulfate (1) | 1.0804(0.11669,3.5399) | 2.015(0.54754,3.9785) | 0.89914 | **0.02** | 0.97 |
| 5alpha-androstan-3beta,17beta-diol monosulfate (2) | 1.1701(0.062618,5.1921) | 2.0388(0.50938,3.8236) | 0.80112 | **0.02** | 0.97 |
| 1-methyl-5-imidazolelactate | 1.8169(0.64208,4.0931) | 1.0511(0.18345,2.5259) | -0.78949 | **0.02** | 0.97 |
| X-13695 | 1.4451(0.13863,7.6889) | 0.60149(0.13863,2.6966) | -1.2645 | **0.02** | 0.97 |
| X-12707 | 1.2413(0.25278,2.531) | 0.73159(0.25278,1.6297) | -0.76274 | **0.02** | 0.97 |
| allopurinol | 1.1921(0.363,3.94) | 0.80557(0.082473,3.045) | -0.56539 | **0.02** | 0.97 |

| | | | | | |
|---|---|---|---|---|---|
| 1-docosahexaenoyl-GPE (22:6)* | 0.97207(0.52828,2.2596) | 1.2152(0.79097,2.0889) | 0.32208 | **0.03** | 0.97 |
| 1-dihomo-linolenoyl-GPE (20:3n3 or 6)* | 1.0103(0.41946,2.38) | 1.502(0.30427,2.8962) | 0.57218 | **0.03** | 0.97 |
| X-13846 | 1.7831(0.15065,11.846) | 0.36514(0.15065,1.7027) | -2.2879 | **0.03** | 0.97 |
| sulfate of piperine metabolite C16H19NO3 (3)* | 1.7125(0.030699,3.9639) | 0.81963(0.083502,3.4765) | -1.0631 | **0.03** | 0.97 |
| piperine | 2.1138(0.014616,5.9067) | 0.89207(0.027531,3.6812) | -1.2446 | **0.03** | 0.97 |
| N6-acetyllysine | 1.0432(0.65319,1.4217) | 0.89514(0.60041,1.0625) | -0.22083 | **0.03** | 0.97 |
| 1-palmitoyl-2-pentadecanoyl-GPC (16:0/15:0)* | 0.89526(0.3381,1.4673) | 1.144(0.81929,1.782) | 0.35377 | **0.03** | 0.97 |
| 5alpha-androstan-3beta,17alpha-diol disulfate | 1.2596(0.10612,6.707) | 2.1128(0.28672,4.8469) | 0.74624 | **0.03** | 0.97 |
| X-25217 | 1.7225(0.087023,16.563) | 0.36786(0.087023,3.457) | -2.2273 | **0.03** | 0.97 |
| X-12906 | 0.77533(0.42542,2.0898) | 1.1183(0.42542,2.0502) | 0.52847 | **0.03** | 0.97 |
| X-24411 | 0.82976(0.29091,2.115) | 1.5251(0.29091,4.1813) | 0.87813 | **0.03** | 0.97 |
| glycosyl-N-(2-hydroxynervonoyl)-sphingosine (d18:1/24:1(2OH))* | 0.88583(0.099912,2.9954) | 0.31039(0.099912,1.2886) | -1.5129 | **0.03** | 0.97 |
| alpha-hydroxyisovalerate | 1.4687(0.47252,6.0707) | 0.8357(0.50793,1.0936) | -0.81347 | **0.03** | 0.97 |
| N6-methyllysine | 1.0804(0.34339,3.7808) | 1.3329(0.45838,3.066) | 0.30297 | **0.03** | 0.97 |
| glucuronide of C10H18O2 (7)* | 2.7799(0.10615,14.691) | 1.565(0.10615,12.203) | -0.82892 | **0.03** | 0.97 |
| 5alpha-androstan-3alpha,17beta-diol disulfate | 0.74958(0.22544,2.5021) | 1.1539(0.29452,2.5857) | 0.62242 | **0.04** | 0.97 |
| cyclo(leu-pro) | 1.1482(0.36524,3.2346) | 0.74258(0.08945,1.9289) | -0.62874 | **0.04** | 0.97 |
| androsterone sulfate | 0.96525(0.31709,2.7631) | 1.6472(0.40194,3.7611) | 0.77102 | **0.04** | 0.97 |
| citraconate/glutaconate | 5.5369(0.37207,35.309) | 2.0782(0.17707,11.527) | -1.4137 | **0.04** | 0.97 |
| dihomo-linolenoylcarnitine (C20:3n3 or 6)* | 1.065(0.40395,2.1799) | 1.4283(0.80877,2.3465) | 0.42339 | **0.04** | 0.97 |
| sulfate of piperine metabolite C16H19NO3 (2)* | 1.5981(0.035245,4.1929) | 0.80401(0.074735,3.0342) | -0.99105 | **0.04** | 0.97 |
| X-12112 | 1.0083(0.31591,3.6727) | 1.2906(0.19933,3.0058) | 0.35612 | **0.04** | 0.97 |
| hexanoylglycine (C6) | 0.3884(0.23515,1.0151) | 1.1705(0.23515,3.7545) | 1.5915 | **0.04** | 0.97 |
| feruloylquinate (1) | 0.58149(0.19266,2.8902) | 0.19266(0.19266,0.19266) | -1.5937 | **0.04** | 0.97 |
| 5alpha-androstan-3alpha,17alpha-diol monosulfate | 0.82679(0.26228,2.0754) | 1.6985(0.26228,5.217) | 1.0387 | **0.04** | 0.97 |
| X-12221 | 1.4059(0.10566,4.2575) | 0.68835(0.10566,2.7967) | -1.0303 | **0.04** | 0.97 |

| | | | | | |
|---|---|---|---|---|---|
| X-13729 | 1.5633(0.067739,5.5981) | 0.80211(0.067739,1.5375) | -0.96275 | **0.04** | 0.97 |
| 2,3-dihydroxy-5-methylthio-4-pentenoate (DMTPA)* | 1.0828(0.69674,1.5655) | 0.92491(0.71113,1.2307) | -0.22736 | **0.04** | 0.97 |
| X-18922 | 1.1554(0.34168,2.5706) | 0.83907(0.51538,1.2716) | -0.46158 | **0.04** | 0.97 |
| X-17348 | 1.2162(0.1801,12.089) | 4.214(0.1801,26.368) | 1.7928 | **0.04** | 0.97 |
| 3-methyl catechol sulfate (2) | 1.4041(0.055876,11.4) | 0.58877(0.055876,2.9166) | -1.2539 | **0.04** | 0.97 |
| X-15674 | 0.48445(0.098927,1.9421) | 0.98044(0.098927,2.8679) | 1.0171 | **0.04** | 0.97 |
| 1-methyl-5-imidazoleacetate | 2.0072(0.20363,5.3761) | 1.0631(0.058333,3.775) | -0.9169 | **0.04** | 0.97 |
| linoleoyl-arachidonoyl-glycerol (18:2/20:4) [1]* | 1.2625(0.41394,2.4386) | 0.85691(0.31584,1.5715) | -0.55903 | **0.04** | 0.97 |
| 1-palmitoyl-2-alpha-linolenoyl-GPC (16:0/18:3n3)* | 0.59418(0.31754,1.3234) | 0.41653(0.31754,1.5054) | -0.51248 | **0.05** | 0.97 |
| 2-ketocaprylate | 1.3333(0.37814,2.2371) | 0.91561(0.25153,1.8668) | -0.54223 | **0.05** | 0.97 |
| 2,6-dihydroxybenzoic acid | 1.087(0.32509,2.2821) | 1.9288(0.56807,4.2793) | 0.82738 | **0.05** | 0.97 |
| X-18886 | 1.0901(0.60536,2.0564) | 0.81802(0.33106,2.1358) | -0.41431 | **0.05** | 0.97 |

~Sixty-two of 1,447 metabolites detected with unadjusted *p*-values of less than 0.05 before rounding are shown.
*Wilcoxon rank-sum test; ^Benjamini and Hochberg method

**TABLE S3C. Entry metabolite levels~ by histological response groups (stringent) for patients who completed the 6-month visit in the *Candida* group.**

| Metabolite | Non-Responders (N=21) | Responders (N=20) | Log2Flod changes Response/No Response | P-value* | |
|---|---|---|---|---|---|
| | | | | Unadjusted | Adjusted^ |
| X-24949 | 0.69339(0.18361,2.1932) | 1.2588(0.18361,2.1666) | 0.8603 | **0.001** | 1 |
| X-18922 | 0.79561(0.31814,1.2483) | 1.1318(0.60575,1.6185) | 0.50849 | **0.002** | 1 |
| X-21815 | 0.29802(0.14339,1.7348) | 1.9512(0.14339,12.042) | 2.7109 | **0.003** | 1 |
| leukotriene B5 | 0.93653(0.21937,1.7726) | 0.48294(0.21937,1.8475) | -0.95549 | **0.004** | 1 |
| X-18886 | 0.85454(0.15988,1.4593) | 1.2352(0.54123,2.0177) | 0.53148 | **0.01** | 1 |
| 1-eicosenoyl-GPE (20:1)* | 1.402(0.088073,5.0705) | 2.4253(0.40567,5.6794) | 0.79067 | **0.01** | 1 |
| isoleucine | 0.92642(0.53519,1.488) | 1.0918(0.52349,1.9558) | 0.23701 | **0.01** | 1 |
| X-21736 | 0.98615(0.25123,3.1004) | 1.3686(0.60288,3.7766) | 0.47278 | **0.01** | 1 |
| 4-hydroxyhippurate | 1.7286(0.30513,6.9719) | 0.92606(0.33225,2.2162) | -0.90045 | **0.02** | 1 |
| 5-(galactosylhydroxy)-lysine | 0.77209(0.22961,1.2172) | 1.0745(0.22961,1.8547) | 0.47677 | **0.02** | 1 |
| cysteine-glutathione disulfide | 0.92006(0.22017,2.2631) | 1.2911(0.57266,2.2141) | 0.4888 | **0.02** | 1 |
| X-17398 | 0.65867(0.25324,5.5462) | 0.27088(0.25324,0.60601) | -1.2819 | **0.02** | 1 |
| sulfate* | 0.91218(0.70424,1.2189) | 1.0108(0.72792,1.2973) | 0.14818 | **0.02** | 1 |
| X-23780 | 1.4354(0.10684,3.084) | 1.2099(0.10684,8.3647) | -0.24655 | **0.02** | 1 |
| isovalerate (C5) | 0.93951(0.45225,1.9262) | 1.2186(0.3622,2.1951) | 0.37524 | **0.03** | 1 |
| 12,13-DiHOME | 0.9166(0.16318,2.3946) | 1.2704(0.65814,2.8999) | 0.47092 | **0.03** | 1 |
| 5-methylthioadenosine (MTA) | 0.91244(0.5461,1.1431) | 1.0442(0.57199,1.3626) | 0.19465 | **0.03** | 1 |
| glutarylcarnitine (C5-DC) | 0.93983(0.486,1.5397) | 1.2415(0.53975,2.04) | 0.40158 | **0.03** | 1 |
| X-14939 | 0.86008(0.1944,1.8797) | 1.2346(0.35791,3.6694) | 0.52144 | **0.03** | 1 |
| 3-phenylpropionate (hydrocinnamate) | 0.87066(0.065799,3.7329) | 1.5897(0.065799,4.5729) | 0.86856 | **0.03** | 1 |
| diphenhydramine | 0.05196(0.05196,0.05196) | 0.32152(0.05196,4.0185) | 2.6294 | **0.04** | 1 |
| 5,6-dihydrothymine | 0.94006(0.6672,1.4314) | 1.1481(0.5973,1.6066) | 0.28844 | **0.04** | 1 |
| glycerol | 0.90979(0.31524,2.667) | 1.2759(0.54348,2.619) | 0.48792 | **0.04** | 1 |
| choline | 0.95988(0.73591,1.3521) | 1.044(0.79357,1.3492) | 0.12123 | **0.04** | 1 |
| formiminoglutamate | 1.0005(0.28956,2.2656) | 1.3554(0.28956,2.7161) | 0.43794 | **0.04** | 1 |
| cinnamoylglycine | 0.81662(0.079964,3.5854) | 1.4908(0.079964,4.3476) | 0.86834 | **0.04** | 1 |
| X-13723 | 2.6526(0.22288,31.255) | 1.1599(0.22288,15.089) | -1.1935 | **0.04** | 1 |
| succinoyltaurine | 0.90232(0.20521,1.7286) | 1.3808(0.39347,3.616) | 0.61376 | **0.04** | 1 |
| 1-oleoyl-2-docosahexaenoyl-GPE (18:1/22:6)* | 0.82329(0.21517,3.7965) | 1.3513(0.21517,3.1799) | 0.71483 | **0.05** | 1 |

| | | | | | |
|---|---|---|---|---|---|
| X-17692 | 0.38637(0.14419,2.3575) | 0.14419(0.14419,0.14419) | -1.422 | **0.05** | 1 |
| 5-hydroxy-2-methylpyridine sulfate | 4.4033(0.020551,16.496) | 1.185(0.020551,8.6926) | -1.8937 | **0.05** | 1 |
| X-25419 | 0.96124(0.090972,4.59) | 2.53(0.090972,26.572) | 1.3961 | **0.05** | 1 |

~Thirty-two of 1,448 metabolites detected with unadjusted *p*-values of less than 0.05 before rounding are shown.
*Wilcoxon rank-sum test; ^Benjamini and Hochberg method

**Table S4A. Summary of comparisons between $\alpha$ diversity of histological responders and non-responders (strict) for patients who completed the 6-month visit.**

| Diversity Index | Group | Non-Responders | Responders | $P_{adj}$* |
|---|---|---|---|---|
| Pielou Evenness | Both | 0.43 ± 0.21 | 0.44 ± 0.22 | 0.97 |
| | PepCan | 0.41 ± 0.20 | 0.45 ± 0.21 | 0.49 |
| | *Candida* | 0.43 ± 0.21 | 0.41 ± 0.20 | 0.73 |
| Faith's Phylogenetic | Both | 24.77 ± 6.75 | 24.70 ± 6.72 | 0.83 |
| | PepCan | 25.01 ± 7.00 | 24.70 ± 7.03 | 0.61 |
| | *Candida* | 24.34 ± 6.91 | 24.77 ± 6.75 | 0.87 |
| Shannon Diversity | Both | 2.51 ± 1.32 | 2.57 ± 1.33 | 0.90 |
| | PepCan | 2.42 ± 1.29 | 2.63 ± 1.30 | 0.46 |
| | *Candida* | 2.51 ± 1.32 | 2.43 ± 1.28 | 0.87 |

* Benjaminin-Hochberg corrected *p*-value of Wilcoxon test

**Table 4B. Summary of comparisons between a diversity between before and after vaccinations in all patients and in subgroups.**

| Diversity Index | Group | Pre-vaccination | Post-Vaccination | Time | $P_{adj}$* |
|---|---|---|---|---|---|
| Pielou Evenness | Both | 0.44 ± 0.22 | 0.40 ± 0.23 | 6 month | 0.16 |
| | | | 0.54 ± 0.21 | 12 month | **0.007** |
| | PepCan | 0.44 ± 0.22 | 0.40 ± 0.24 | 6 month | 0.34 |
| | | | 0.55 ± 0.22 | 12 month | **0.01** |
| | *Candida* | 0.43 ± 0.21 | 0.40 ± 0.23 | 6 month | 0.74 |
| | | | 0.54 ± 0.22 | 12month | **0.03** |
| | Responders | 0.44 ± 0.22 | 0.40 ± 0.23 | 6 month | 0.36 |
| | | | 0.54 ± 0.22 | 12 month | **0.03** |
| | Non-Responders | 0.43 ± 0.21 | 0.40 ± 0.24 | 6 month | 0.84 |
| | | | 0.55 ± 0.21 | 12 month | 0.07 |
| Faith's Phylogenetic | Both | 24.60 ± 6.72 | 66.63 ± 21.31 | 6 month | **<0.001** |
| | | | 73.51 ± 13.78 | 12 month | **<0.001** |
| | PepCan | 24.72 ± 6.78 | 66.86 ± 21.55 | 6 month | **<0.001** |
| | | | 73.71 ± 14.21 | 12 month | **<0.001** |
| | *Candida* | 24.77 ± 6.76 | 66.30 ± 21.26 | 6 month | **<0.001** |
| | | | 73.51 ± 13.78 | 12month | **<0.001** |
| | Responders | 24.70 ± 6.72 | 66.80 ± 21.40 | 6 month | **<0.001** |
| | | | 73.42 ± 13.91 | 12 month | **<0.001** |
| | Non-Responders | 24.77 ± 6.76 | 66.18 ± 21.57 | 6 month | **<0.001** |
| | | | 73.63 ± 13.95 | 12 month | **<0.001** |
| Shannon Diversity | Both | 2.55 ± 1.32 | 2.90 ± 1.77 | 6 month | 0.49 |
| | | | 4.06 ± 1.68 | 12 month | **<0.001** |
| | PepCan | 2.55 ± 1.33 | 2.89 ± 1.79 | 6 month | 0.87 |
| | | | 4.19 ± 1.71 | 12 month | **<0.001** |
| | *Candida* | 2.51 ± 1.32 | 2.90 ± 1.78 | 6 month | 0.31 |
| | | | 4.06 ± 1.68 | 12month | **<0.001** |
| | Responders | 2.57 ± 1.33 | 2.90 ± 1.78 | 6 month | 0.68 |
| | | | 4.07 ± 1.70 | 12 month | **<0.001** |
| | Non-Responders | 2.51 ± 1.32 | 2.90 ± 1.80 | 6 month | 0.24 |
| | | | 4.15 ± 1.68 | 12 month | **0.001** |

* Benjaminin-Hochberg corrected *p*-value of Wilcoxon test

**Table S4C. The top five differentially abundant taxa after vaccinations in all patients and in subgroups.**

| Comparison | Group | Family | Genus |
|---|---|---|---|
| 6-month vs pre-vaccination | Both | *Pseudomonadaceae, Bifidobacteriaceae, Yersiniaceae, Nitrososphaeraceae, Enterobacteriaceae* | *Pseudomonas, Gardnerella, Finegoldia, Enterobacteriaceae, Nitrososphaeraceae* |
| 12-month vs pre-vaccination | | *Bifidobacteriaceae, Pseudomonadaceae, Yersiniaceae, Moraxellaceae, Lactobacillaceae* | *Gardnerella, Pseudomonas, Enhydrobacter, Lacticaseibacillus, Nitrososphaeraceae* |
| 6-month vs pre-vaccination | Histological Responders | *Enterobacteriaceae, Yersiniaceae, Micrococcaceae, Burkholderiaceae, Nitrososphaeraceae* | *Ralstonia, Nitrososphaeraceae, Blastococcus, Rubrobacter* |
| 12-month vs pre-vaccination | | *Nitrososphaeraceae, Micrococcaceae, Burkholderiaceae, Nitrososphaeraceae, Geodermatophilaceae* | *Nitrososphaeraceae, Ralstonia, Nitrososphaeraceae, Blastococcus, Rubrobacter* |
| 6-month vs pre-vaccination | Histological Non-Responders | *Bifidobacteriaceae, Pseudomonadaceae, Yersiniaceae, Enterobacteriaceae, Nitrososphaeraceae* | *Gardnerella, Pseudomonas, Nitrososphaeraceae* |
| 12-month vs pre-vaccination | | *Pseudomonadaceae, Yersiniaceae, Enterobacteriaceae, Rubrobacteriaceae* | *Pseudomonas, Rubrobacter* |
| 6-month vs pre-vaccination | PepCan | *Yersiniaceae, Bifidobacteriaceae, Enterobacteriaceae, Nitrososphaeraceae* | *Gardnerella, Nitrososphaeraceae* |
| 12-month vs pre-vaccination | | *Pseudomonadaceae, Enterobacteriaceae, Yersiniaceae* | *Pseudomonas* |
| 6-month vs pre-vaccination | *Candida* | *Xanthomonadaceae, Yersiniaceae, Nitrososphaeraceae, Micrococcaceae, Nitrososphaeraceae* | *Stenotrophomonas, Nitrososphaeraceae, Rubrobacter* |
| 12-month vs pre-vaccination | | *Yersiniaceae, Micrococcaceae, Nitrososphaeraceae, Bacteroidaceae, Rubrobacteriaceae* | *Nitrososphaeraceae, Bacteroides, Rubrobacter, Chryseobacterium* |

Detection of microbial features differentially enriched across treatment groups were assessed with standard and machine-learning–based analyses, through the use of ALDEx2 (ANOVA-like differential expression, version 2). ALDEx2 is a robust compositionally aware differential abundance method that does not require rarefying of microbial amplicon data. This approach uses the Benjamini-Hochberg corrected *p*-value of Wilcoxon test, to detect differentially abundant features (i.e. unique microbial sequence) between time points.

**Table S5A. Comparison of responders to non-responders (stringent) with respect to HLA types with total allele counts of 10 or greater for both groups for patients who completed the 6-month visit.**

| HLA Type | Non-Responders | Responders | *P*-value* | |
|---|---|---|---|---|
| | | | Unadjusted | Adjusted^ |
| A01 | 0.1548 (13/84) | 0.1290 (8/62) | 0.81 | >0.99 |
| A02 | 0.2738 (23/84) | 0.3871 (24/62) | 0.16 | 0.80 |
| A03 | 0.0833 (7/84) | 0.0806 (5/62) | >0.99 | >0.99 |
| A24 | 0.1310 (11/84) | 0.0806 (5/62) | 0.43 | 0.94 |
| B07 | 0.1786 (15/84) | 0.0781 (5/64) | 0.09 | 0.80 |
| B08 | 0.0714 (6/84) | 0.0938 (6/64) | 0.76 | >0.99 |
| B15 | 0.0357 (3/84) | 0.1250 (8/64) | 0.06 | 0.80 |
| B35 | 0.1310 (11/84) | 0.0781 (5/64) | 0.42 | 0.94 |
| B40 | 0.0476 (4/84) | 0.0938 (6/64) | 0.33 | 0.94 |
| B44 | 0.1429 (12/84) | 0.1562 (10/64) | 0.82 | >0.99 |
| C03 | 0.1023 (9/88) | 0.1429 (8/56) | 0.60 | 0.94 |
| C04 | 0.0909 (8/88) | 0.0714 (4/56) | 0.77 | >0.99 |
| C05 | 0.0455 (4/88) | 0.1071 (6/56) | 0.19 | 0.80 |
| C06 | 0.1250 (11/88) | 0.0714 (4/56) | 0.41 | 0.94 |
| C07 | 0.3750 (33/88) | 0.3214 (18/56) | 0.59 | 0.94 |
| DP02 | 0.1500 (9/60) | 0.0476 (2/42) | 0.12 | 0.80 |
| DP03 | 0.1000 (6/60) | 0.0952 (4/42) | >0.99 | >0.99 |
| DP04 | 0.5000 (30/60) | 0.5238 (22/42) | 0.84 | >0.99 |
| DQ02 | 0.2273 (20/88) | 0.2031 (13/64) | 0.84 | >0.99 |
| DQ03 | 0.3409 (30/88) | 0.4531 (29/64) | 0.18 | 0.80 |
| DQ04 | 0.0795 (7/88) | 0.0781 (5/64) | >0.99 | >0.99 |
| DQ05 | 0.1023 (9/88) | 0.1094 (7/64) | >0.99 | >0.99 |
| DQ06 | 0.2500 (22/88) | 0.1562 (10/64) | 0.23 | 0.85 |
| DR01 | 0.0682 (6/88) | 0.0938 (6/64) | 0.56 | 0.94 |
| DR03 | 0.0795 (7/88) | 0.1250 (8/64) | 0.41 | 0.94 |
| DR04 | 0.1932 (17/88) | 0.2344 (15/64) | 0.55 | 0.94 |
| DR07 | 0.1705 (15/88) | 0.1250 (8/64) | 0.50 | 0.94 |
| DR11 | 0.0682 (6/88) | 0.0938 (6/64) | 0.56 | 0.94 |
| DR13 | 0.0909 (8/88) | 0.0938 (6/64) | >0.99 | >0.99 |
| DR15 | 0.2159 (19/88) | 0.0469 (3/64) | **0.004** | 0.13 |

*Fisher’s exact test (two-sided); ^Benjamini and Hochberg method.

**Table S5B. Comparison of responders to non-responders with respect to HLA types with total allele counts of 10 or greater for the PepCan group for patients who completed the 6-month visit.**

| HLA Type | Non-Responders | Responders | *P*-value* | |
|---|---|---|---|---|
| | | | Unadjusted | Adjusted^ |
| A01 | 0.1591 (7/44) | 0.1250 (3/24) | >0.99 | >0.99 |
| A02 | 0.2045 (9/44) | 0.5417 (13/24) | **0.007** | 0.08 |
| B07 | 0.2143 (9/42) | 0.1250 (3/24) | 0.51 | 0.86 |
| B44 | 0.1429 (6/42) | 0.1667 (4/24) | >0.99 | >0.99 |
| C07 | 0.4130 (19/46) | 0.4583 (11/24) | 0.80 | 0.96 |
| DP04 | 0.5000 (13/26) | 0.5714 (8/14) | 0.75 | 0.96 |
| DQ02 | 0.2826 (13/46) | 0.2083 (5/24) | 0.58 | 0.86 |
| DQ03 | 0.3261 (15/46) | 0.4583 (11/24) | 0.31 | 0.84 |
| DQ06 | 0.3043 (14/46) | 0.2083 (5/24) | 0.57 | 0.86 |
| DR04 | 0.1087 (5/46) | 0.2500 (6/24) | 0.17 | 0.67 |
| DR07 | 0.2391 (11/46) | 0.1250 (3/24) | 0.35 | 0.84 |
| DR15 | 0.2609 (12/46) | 0.0833 (2/24) | 0.12 | 0.67 |

*Fisher’s exact test (two-sided); ^Benjamini and Hochberg method.

**Table S5C. Comparison of responders to non-responders with respect to HLA types with total allele counts of 10 or greater for the *Candida* group for patients who completed the 6-month visit.**

| HLA Type | Non-Responders | Responders | *P*-value* | |
|---|---|---|---|---|
| | | | Unadjusted | Adjusted^ |
| A01 | 0.1500 (6/40) | 0.1316 (5/38) | >0.99 | >0.99 |
| A02 | 0.3500 (14/40) | 0.2895 (11/38) | 0.63 | >0.99 |
| A24 | 0.1500 (6/40) | 0.1053 (4/38) | 0.74 | >0.99 |
| B35 | 0.1667 (7/42) | 0.1000 (4/40) | 0.52 | >0.99 |
| B44 | 0.1429 (6/42) | 0.1500 (6/40) | >0.99 | >0.99 |
| C03 | 0.1190 (5/42) | 0.2188 (7/32) | 0.34 | >0.99 |
| C07 | 0.3333 (14/42) | 0.2188 (7/32) | 0.31 | >0.99 |
| DP04 | 0.5000 (17/34) | 0.5000 (14/28) | >0.99 | >0.99 |
| DQ02 | 0.1667 (7/42) | 0.2000 (8/40) | 0.78 | >0.99 |
| DQ03 | 0.3571 (15/42) | 0.4500 (18/40) | 0.50 | >0.99 |
| DQ05 | 0.1429 (6/42) | 0.1500 (6/40) | >0.99 | >0.99 |
| DQ06 | 0.1905 (8/42) | 0.1250 (5/40) | 0.55 | >0.99 |
| DR04 | 0.2857 (12/42) | 0.2250 (9/40) | 0.62 | >0.99 |

*Fisher’s exact test (two-sided); ^Benjamini and Hochberg method.

**Table S6. Summary of vaccine-induced HPV-specific T cells detected in cervix.**

| ID | Treatment | Histologic Response | 6-Month | | | 12-Month | | |
|---|---|---|---|---|---|---|---|---|
| | | | CD4 T Cells | CD8 T Cells | % HPV | CD4 T Cells | CD8 T Cells | % HPV |
| 32 | PepCan | NR | 8 | 18 | 1.7% (26/1529) | 9 | 18 | 2.1% (27/1259) |
| 39 | PepCan | R | 57 | 42 | 7.7% (99/1289) | 125 | 81 | 7.0% (206/2937) |
| 46 | PepCan | NR | 11 | 13 | 4.8% (24/495) | 2 | 3 | 3.0% (5/166) |
| 51 | *Candida* | NR | 29 | 51 | 11.4% (80/703) | 21 | 38 | 14.1% (59/418) |
| 58 | PepCan | NR | 44 | 130 | 6.1% (174/2866) | 23 | 26 | 3.2% (49/1525) |

#### SUPPLEMENTARY FIGURES AND FIGURE LEGENDS

**Figure S1. Trial design.**

Vaccination (PepCan or *Candida*) visits were scheduled 3 weeks apart for patients who had biopsy-confirmed CIN2/3. Research blood draws were performed at visits 1 (pre-vaccination), 2, 3, 4, 6-month visit, and 12-month visit. Liquid-based cytology samples (Thin-Prep) were collected at screening visit, 6-month visit, and 12-month visit. Biopsies were performed at screening visit, at 6-month visit for suspicion of progressive disease, and at 12-month visit. At the 12-month visit, at least four biopsies were taken (one from each cervical quadrant), and the biopsies were examined by two pathologists blinded to each other's diagnoses.

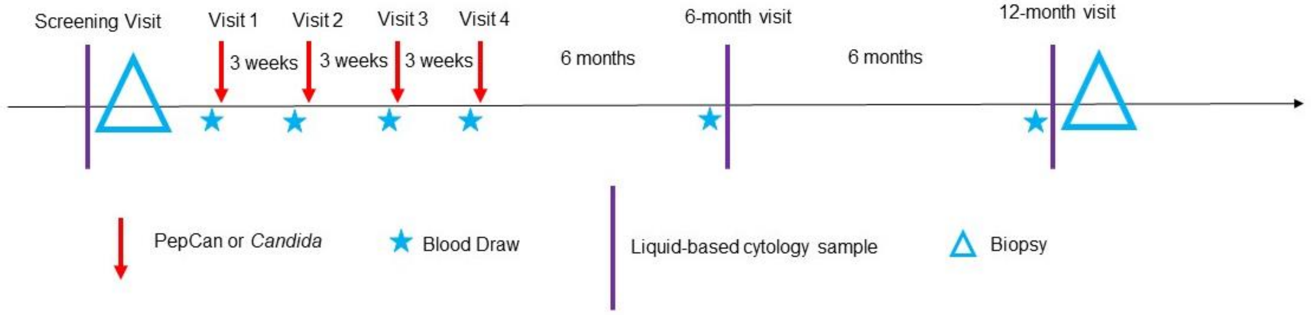

**Figure S2. Histological responses and subgroup comparisons.**

Panel A shows the analyses of the intention-to-treat populations and panel B shows the analyses of the per-protocol analyses. Both panels used the lenient criterion in which the complete responders (regression to no CIN) and partial responders (regression to CIN 1) were considered to have had responses. The Clopper-Pearson method was used to determine the 95% confidence intervals.

**A**

| Vaccine/Subgroup | n/N | Response (95% CI) |
|---|---|---|
| Both | 39/81 | 48.1 (36.9, 59.5) |
| < 25 year old | 7/15 | 46.7 (21.3, 73.4) |
| ≥ 25 year old | 32/66 | 48.5 (36.0, 61.1) |
| CIN2 | 19/26 | 73.1 (52.2, 88.4) |
| >CIN2 | 20/55 | 36.4 (23.8, 50.4) |
| ≤2 cervical quadrants | 30/59 | 50.8 (37.5, 64.1) |
| >2 cervical quadrants | 9/20 | 45.0 (23.1, 68.5) |
| PepCan | 15/39 | 38.5 (23.4, 55.4) |
| < 25 year old | 3/6 | 50.0 (11.8, 88.2) |
| ≥ 25 year old | 12/33 | 36.4 (20.4, 54.9) |
| CIN2 | 5/10 | 50.0 (18.7, 81.3) |
| >CIN2 | 10/29 | 34.5 (17.9, 54.3) |
| ≤2 cervical quadrants | 12/29 | 41.4 (23.5, 61.1) |
| >2 cervical quadrants | 3/9 | 33.3 (7.5, 70.1) |
| Candida | 24/42 | 57.1 (41.0, 72.3) |
| < 25 year old | 4/9 | 44.4 (13.7, 78.8) |
| ≥ 25 year old | 20/33 | 60.6 (42.1, 77.1) |
| CIN2 | 14/16 | 87.5 (61.7, 98.4) |
| >CIN2 | 10/26 | 38.5 (20.2, 59.4) |
| ≤2 cervical quadrants | 18/30 | 60.0 (40.6, 77.3) |
| >2 cervical quadrants | 6/11 | 54.5 (23.4, 83.3) |

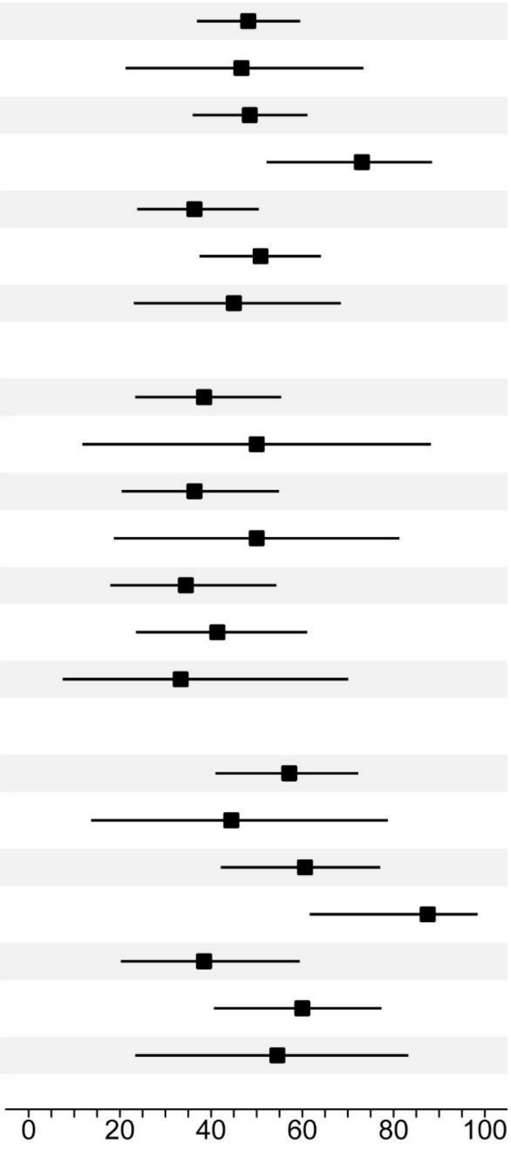

**B**

| Vaccine/Subgroup | n/N | Response (95% CI) |
|---|---|---|
| Both | 35/53 | 66.0 (51.7, 78.5) |
| < 25 year old | 6/9 | 66.7 (29.9, 92.5) |
| ≥ 25 year old | 29/44 | 65.9 (50.1, 79.5) |
| CIN2 | 18/22 | 81.8 (59.7, 94.8) |
| >CIN2 | 17/31 | 54.8 (36.0, 72.7) |
| ≤2 cervical quadrants | 27/41 | 65.9 (49.4, 79.9) |
| >2 cervical quadrants | 8/12 | 66.7 (34.9, 90.1) |
| PepCan | 13/24 | 54.2 (32.8, 74.4) |
| < 25 year old | 3/6 | 50.0 (11.8, 88.2) |
| ≥ 25 year old | 10/18 | 55.6 (30.8, 78.5) |
| CIN2 | 5/7 | 71.4 (29.0, 96.3) |
| >CIN2 | 8/17 | 47.1 (23.0, 72.2) |
| ≤2 cervical quadrants | 11/19 | 57.9 (33.5, 79.7) |
| >2 cervical quadrants | 2/5 | 40.0 (5.3, 85.3) |
| Candida | 22/29 | 75.9 (56.5, 89.7) |
| < 25 year old | 3/3 | 100.0 (29.2, 100.0) |
| ≥ 25 year old | 19/26 | 73.1 (52.2, 88.4) |
| CIN2 | 13/15 | 86.7 (59.5, 98.3) |
| >CIN2 | 9/14 | 64.3 (35.1, 87.2) |
| ≤2 cervical quadrants | 16/22 | 72.7 (49.8, 89.3) |
| >2 cervical quadrants | 6/7 | 85.7 (42.1, 99.6) |

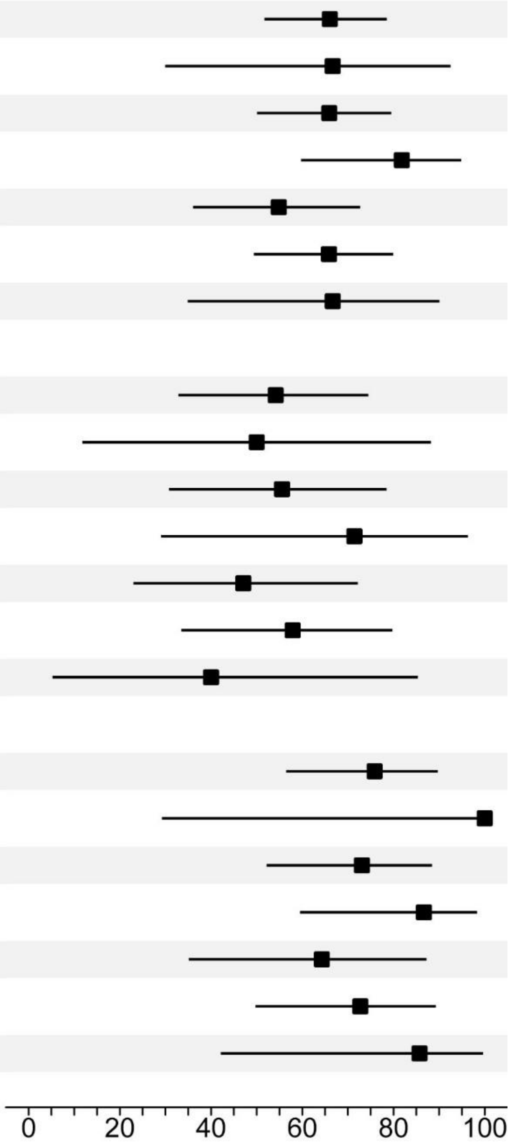

**Figure S3 Representative ELISPOT assay results.**

ELISPOT assays were performed, for patients who completed the 6-month visit, testing 10 regions within the HPV 16 E6 protein at visit 1 (pre-vaccination), visit 3 (post-2 vaccinations), 6-month visit, and 12-month visit. Patients 39 and 58 were histological complete responders (i.e., regressed to no CIN) while patients 23 and 46 were histological non-responders (i.e., had persistent CIN2/3). Patients 39, 46, and 58 received PepCan while patient 23 received *Candida*. Epitope spreading to HPV 35 E6 protein was examined for subject 46 since she was HPV 35-positive at entry, but epitope spreading was not detected. Subject 58 was HPV 16-positive at entry, and demonstrated epitope spreading to HPV 16 E7 protein. The numbers in boxes show positivity indices of 2 or greater. *P*-values less than 0.05 before rounding (two-sided paired *t*-test) are shown for statistically significant increases in T cell responses after vaccination.

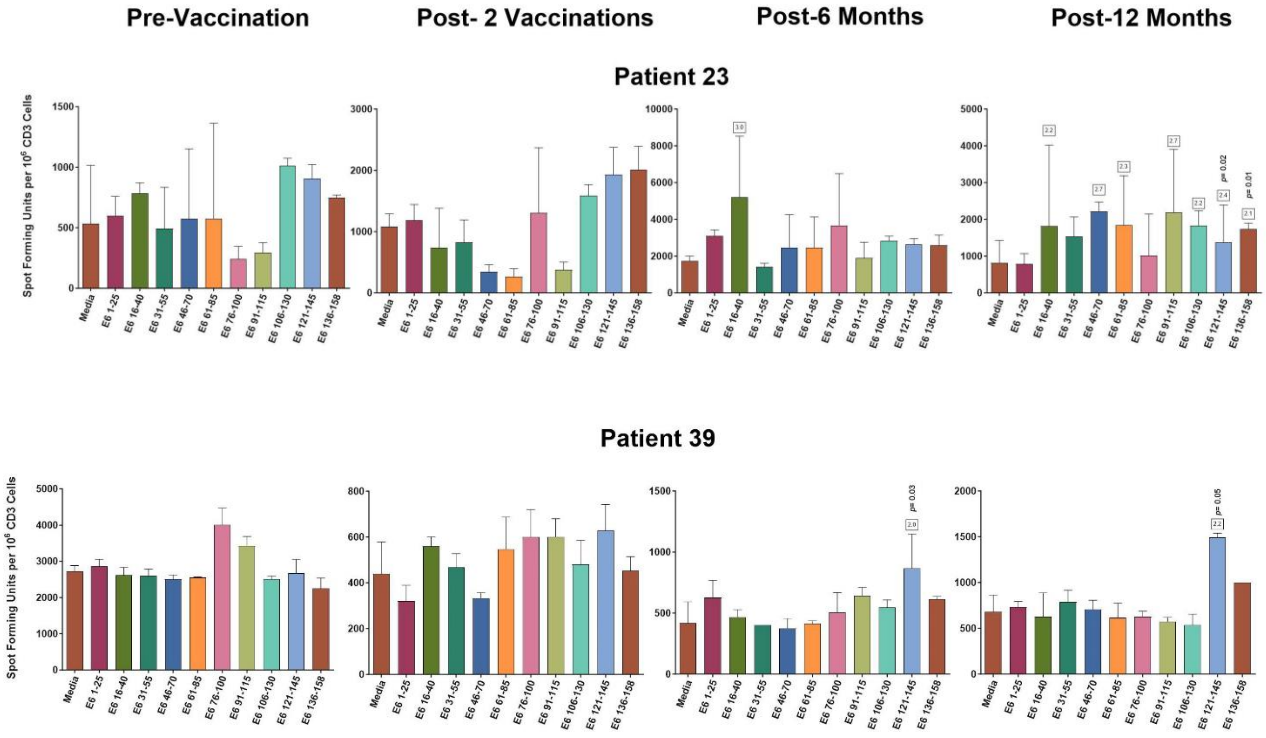

#### Patient 46

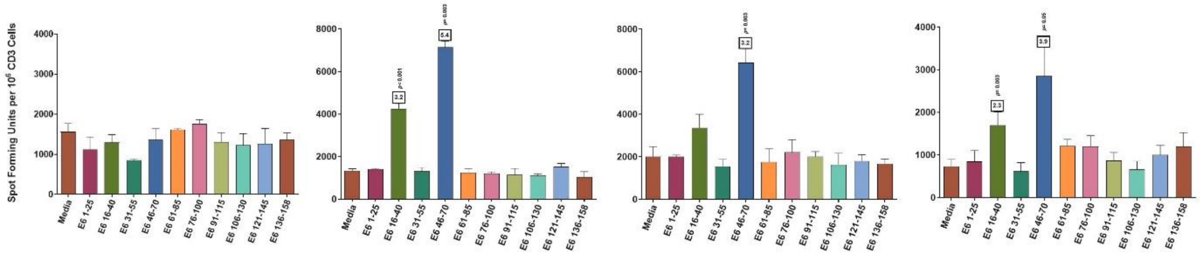

#### No epitope spreading to HPV 35 E6

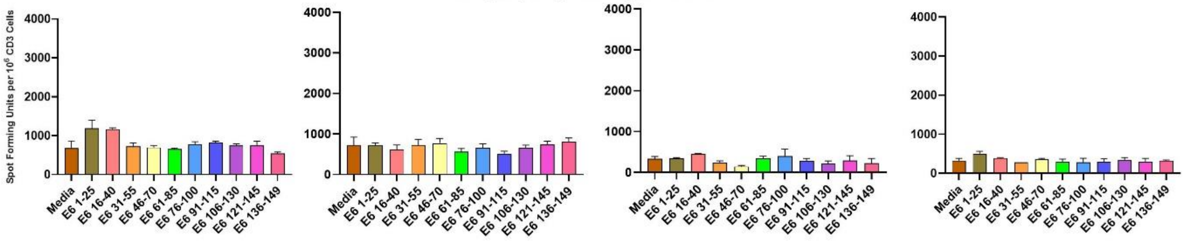

#### Patient 58

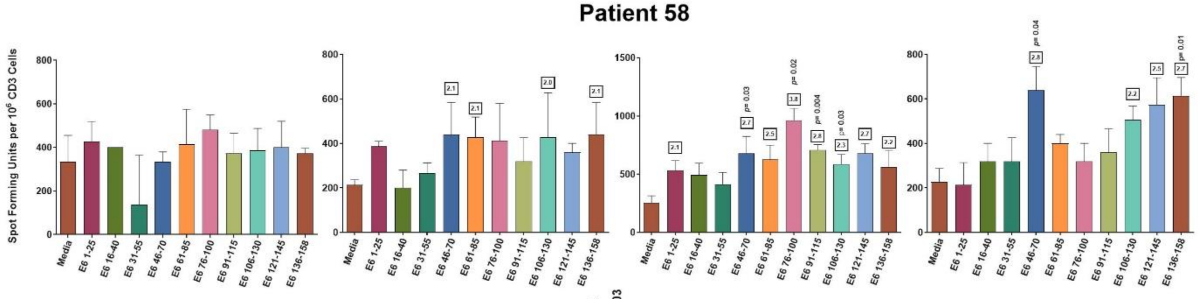

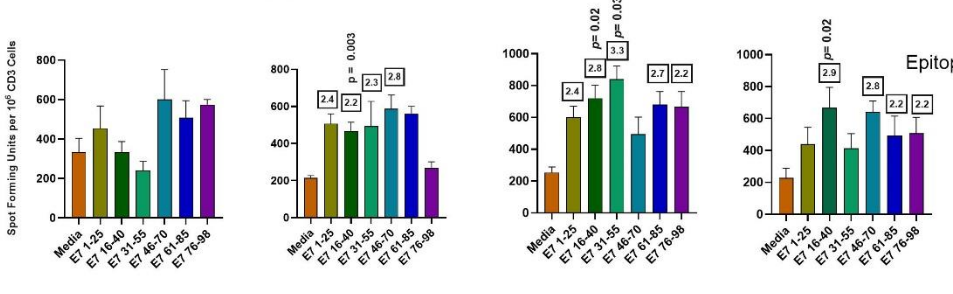

Epitope spreading to HPV 16 E7

**Figure S4 Peripheral immune cell analyses.**

Panel A shows representative raw data from T cell analysis of Patient 19 who was in the *Candida* group, and she was a non-responder. The cells were gated on lymphocytes. CD4+Tbet+ cells represent Th1 cells, CD4+Gata+ T cells represent Th2 cells, and CD4+CD25+FoxP3+ cells represent Treg. Panel B shows representative raw data from the same patient for examining MDSC1 cells which are CD33-, IL4Ralpha-, CD11b- CD14- positive and HLA-DR-intermediate/negative. All cells were gated except for debris, and were sequentially gated further as shown. Panel C shows representative analysis of MDSC2 cells which are CD14-positive and HLA-DR-low/negative. Panel D shows comparisons of Th1, Th2, Treg, MDSC1 and MDSC2 between histological responders and non-responders (stringent) at pre-vaccination, post-2 vaccination, post-6 months, and post-12 months for patients who completed the 6-month visit in both treatment groups. Panel E shows comparisons of Th1, Th2, Treg, MDSC1 and MDSC2 between histological responders and non-responders (stringent) at pre-vaccination, post-2 vaccination, post-6 months, and post-12 months for patients who completed the 6-month visit in the PepCan group. Panel F shows comparisons of Th1, Th2, Treg, MDSC1 and MDSC2 between histological responders and non-responders (stringent) at pre-vaccination, post-2 vaccination, post-6 months, and post-12 months for the patients who completed the 6-month visit in the *Candida* group. Panel G shows percentages of Th1, Th2, Tregs, MDSC1, and MDSC2 at pre-vaccination, post-2 vaccination, post-6 months, and post-12 months for the patients who completed the 6-month visit in both treatment groups. Panel H shows percentages of Th1, Th2, Tregs, MDSC1, and MDSC2 at pre-vaccination, post-2 vaccination, post-6 months, and post-12 months for the patients who completed the 6-month visit in the PepCan group. Panel I shows percentages of Th1, Th2, Tregs, MDSC1, and MDSC2 at pre-vaccination, post-2 vaccination, post-6 months, and post-12months for the patients who completed the 6-month visit the *Candida* group. Unadjusted *p*-values (two-sided paired *t*-test) less than 0.05 before rounding, and corresponding adjusted *p*-values (shown as “q”) are shown (Benjamini and Hochberg method).

**A**

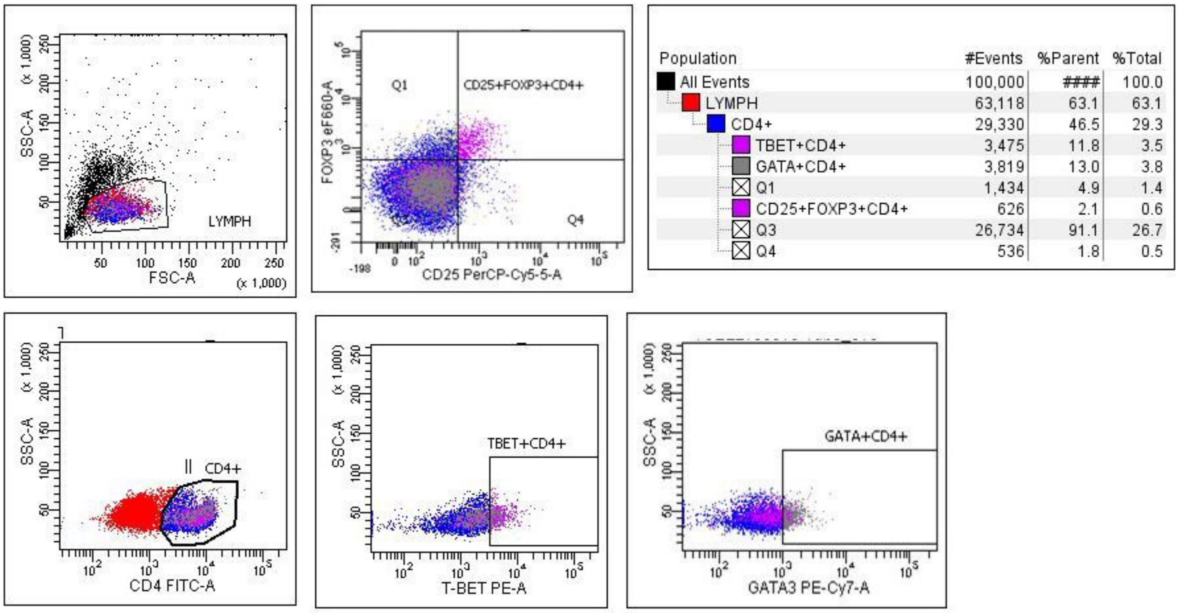

SSC-A (x 1,000)
FSC-A (x 1,000)
LYMPH
Q1
CD25+FOXP3+CD4+
Q4
FOXP3 eF660-A
CD25 PerCP-Cy5-5-A
Population #Events %Parent %Total
All Events 100,000 #### 100.0
LYMPH 63,118 63.1 63.1
CD4+ 29,330 46.5 29.3
TBET+CD4+ 3,475 11.8 3.5
GATA+CD4+ 3,819 13.0 3.8
Q1 1,434 4.9 1.4
CD25+FOXP3+CD4+ 626 2.1 0.6
Q3 26,734 91.1 26.7
Q4 536 1.8 0.5
CD4+
CD4 FITC-A
TBET+CD4+
T-BET PE-A
GATA+CD4+
GATA3 PE-Cy7-A

**B**

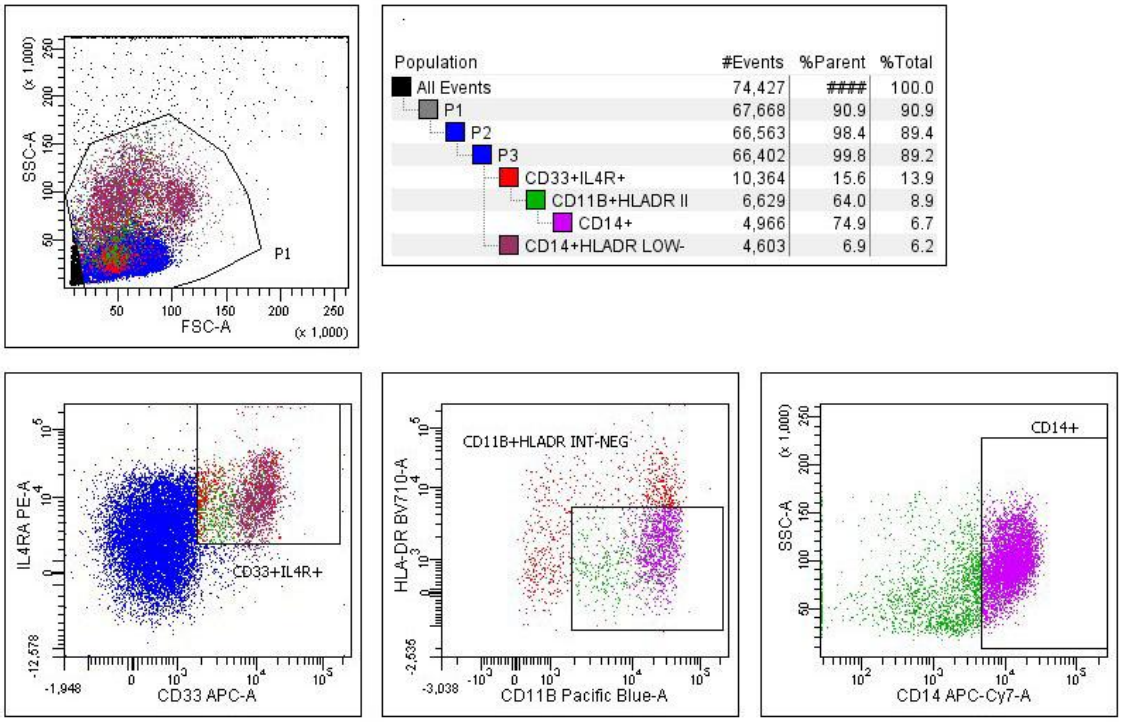

SSC-A (x 1,000)
FSC-A (x 1,000)
P1
Population #Events %Parent %Total
All Events 74,427 #### 100.0
P1 67,668 90.9 90.9
P2 66,563 98.4 89.4
P3 66,402 99.8 89.2
CD33+IL4R+ 10,364 15.6 13.9
CD11B+HLADR II 6,629 64.0 8.9
CD14+ 4,966 74.9 6.7
CD14+HLADR LOW- 4,603 6.9 6.2
IL4RA PE-A
CD33 APC-A
CD33+IL4R+
CD11B+HLADR INT-NEG
HLA-DR BV710-A
CD11B Pacific Blue-A
CD14+
SSC-A
CD14 APC-Cy7-A

**C**

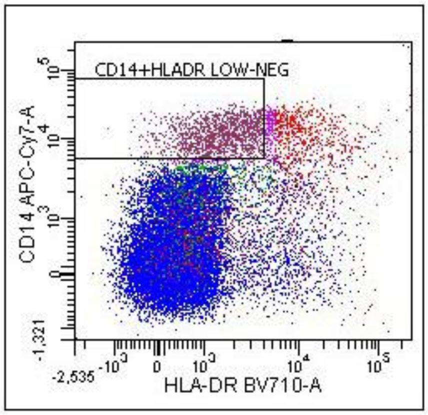

| Population | #Events | %Parent | %Total |
|---|---|---|---|
| All Events | 74,427 | #### | 100.0 |
| P1 | 67,668 | 90.9 | 90.9 |
| P2 | 66,563 | 98.4 | 89.4 |
| P3 | 66,402 | 99.8 | 89.2 |
| CD33+IL4R+ | 10,364 | 15.6 | 13.9 |
| CD11B+HLADR II | 6,629 | 64.0 | 8.9 |
| CD14+ | 4,966 | 74.9 | 6.7 |
| CD14+HLADR LOW- | 4,603 | 6.9 | 6.2 |

**D**

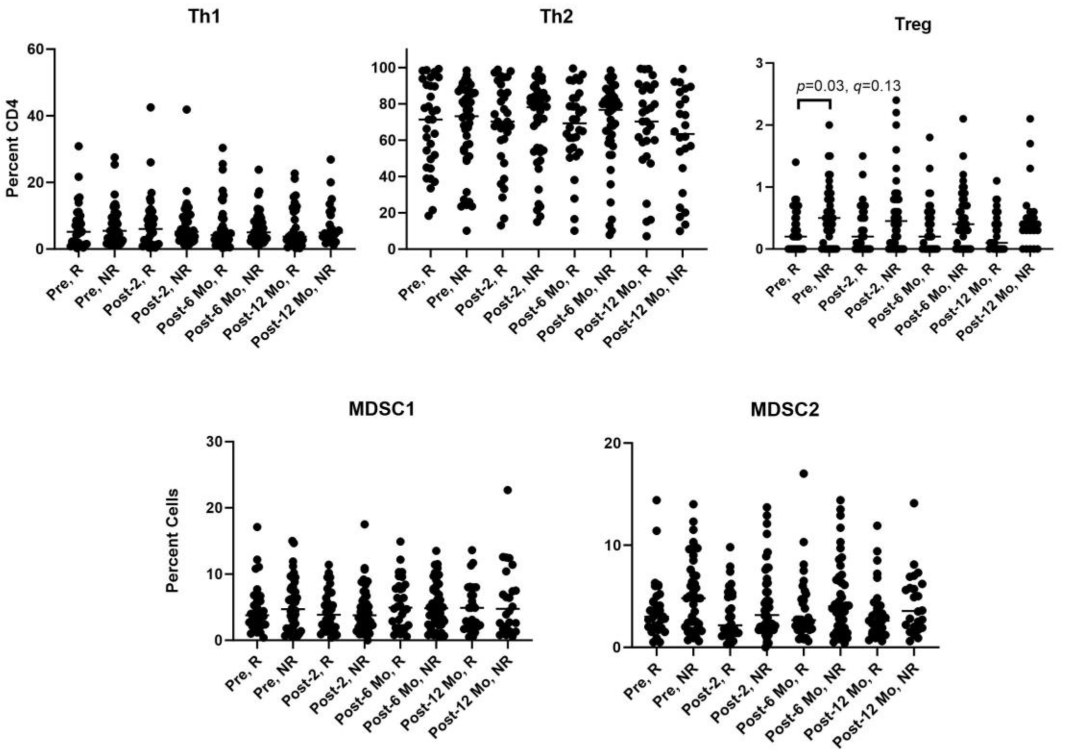
Th1
Th2
Treg
Percent CD4
60
40
20
0
100
80
60
40
20
0
3
2
1
0
p=0.03, q=0.13
Pre, R
Pre, NR
Post-2, R
Post-2, NR
Post-6 Mo, R
Post-6 Mo, NR
Post-12 Mo, R
Post-12 Mo, NR
MDSC1
MDSC2
Percent Cells
30
20
10
0
20
10
0

**E**

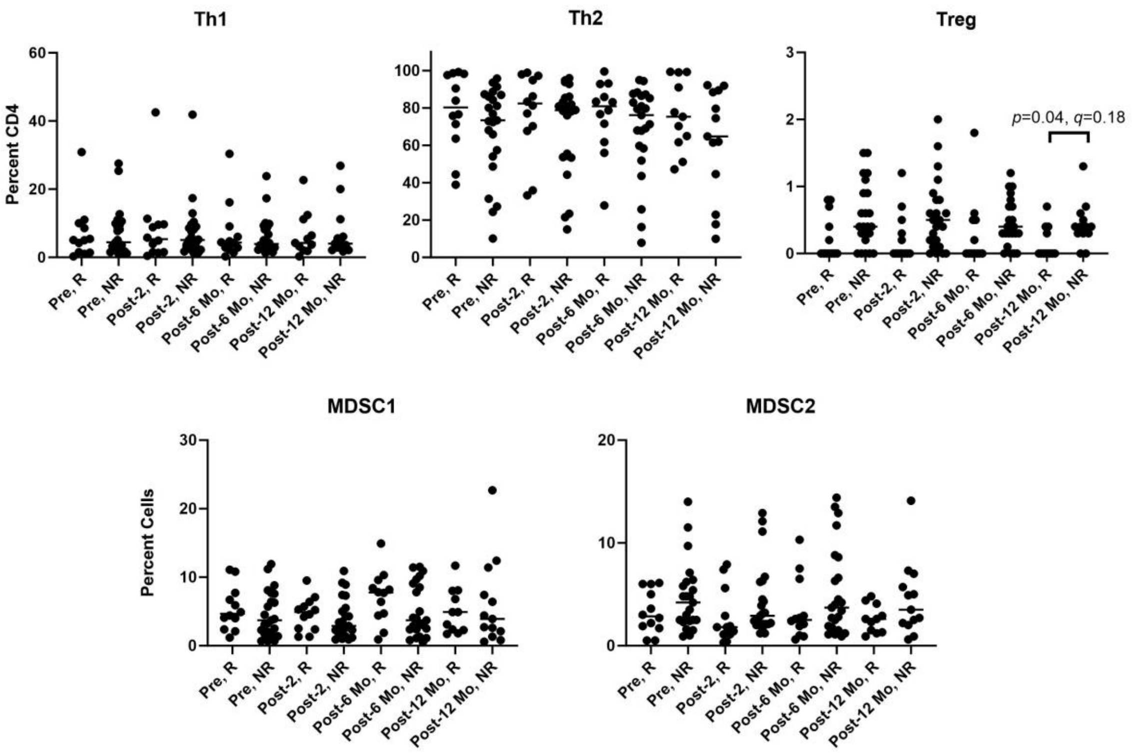
Th1
Th2
Treg
Percent CD4
60
40
20
0
100
80
60
40
20
0
3
2
1
0
p=0.04, q=0.18
Pre, R
Pre, NR
Post-2, R
Post-2, NR
Post-6 Mo, R
Post-6 Mo, NR
Post-12 Mo, R
Post-12 Mo, NR
MDSC1
MDSC2
Percent Cells
30
20
10
0
20
10
0

**F**

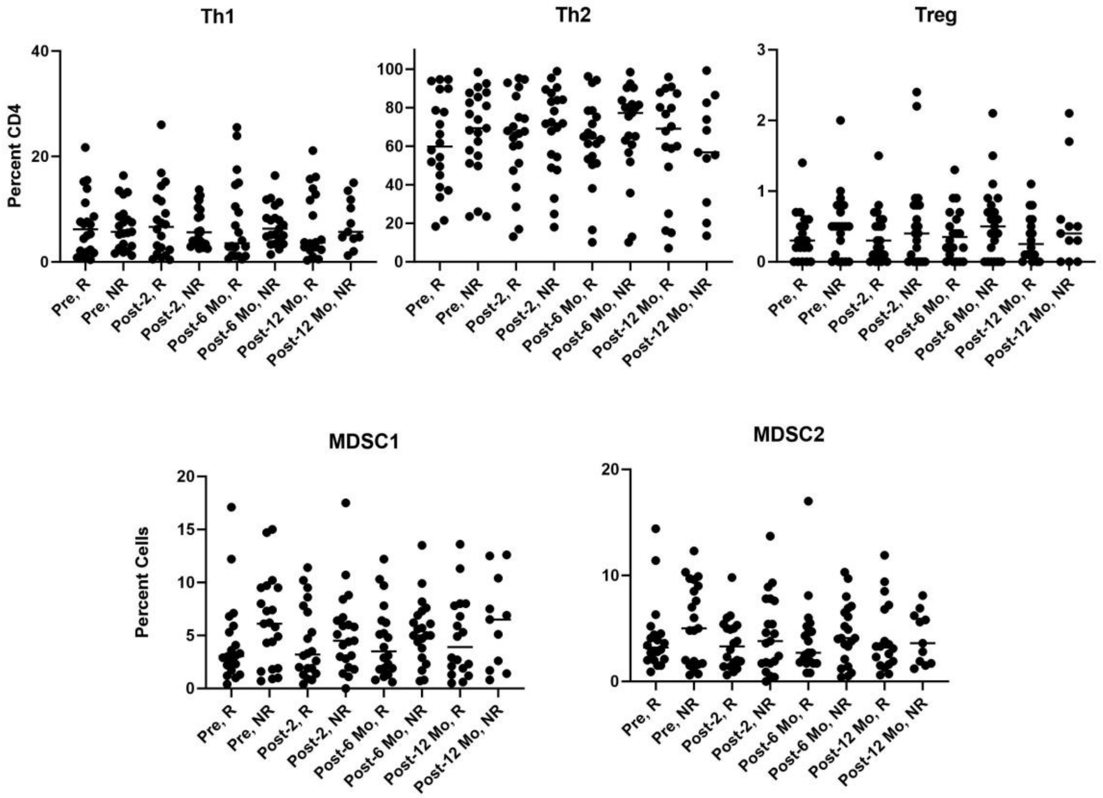

Th1
Th2
Treg
Percent CD4
MDSC1
MDSC2
Percent Cells
Pre, R
Pre, NR
Post-2, R
Post-2, NR
Post-6 Mo, R
Post-6 Mo, NR
Post-12 Mo, R
Post-12 Mo, NR

**G**

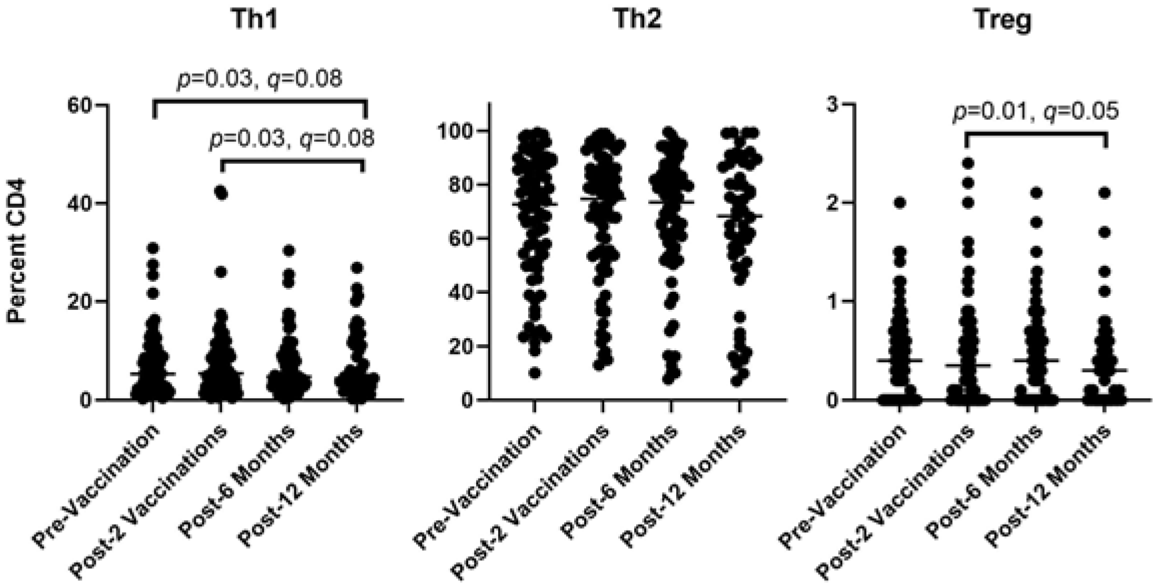

Th1
Th2
Treg
p=0.03, q=0.08
p=0.03, q=0.08
p=0.01, q=0.05
Percent CD4
Pre-Vaccination
Post-2 Vaccinations
Post-6 Months
Post-12 Months

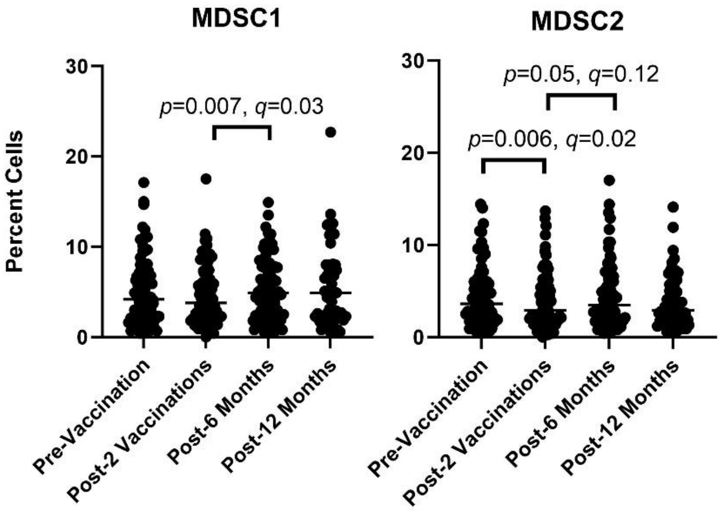

MDSC1
MDSC2
p=0.007, q=0.03
p=0.05, q=0.12
p=0.006, q=0.02
Percent Cells
Pre-Vaccination
Post-2 Vaccinations
Post-6 Months
Post-12 Months

H

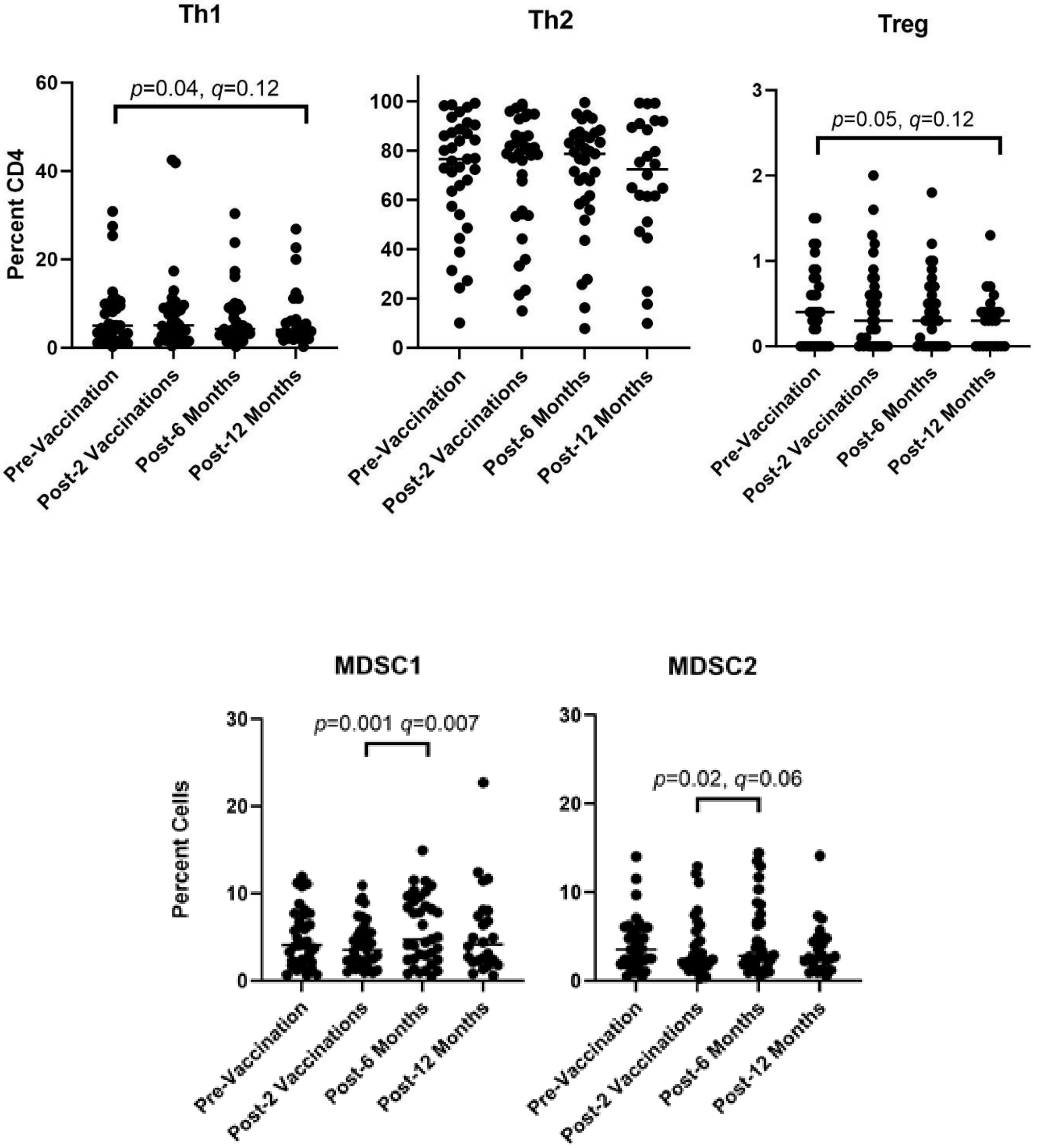
Th1
Th2
Treg
Percent CD4
p=0.04, q=0.12
p=0.05, q=0.12
Pre-Vaccination
Post-2 Vaccinations
Post-6 Months
Post-12 Months
MDSC1
MDSC2
Percent Cells
p=0.001 q=0.007
p=0.02, q=0.06

I

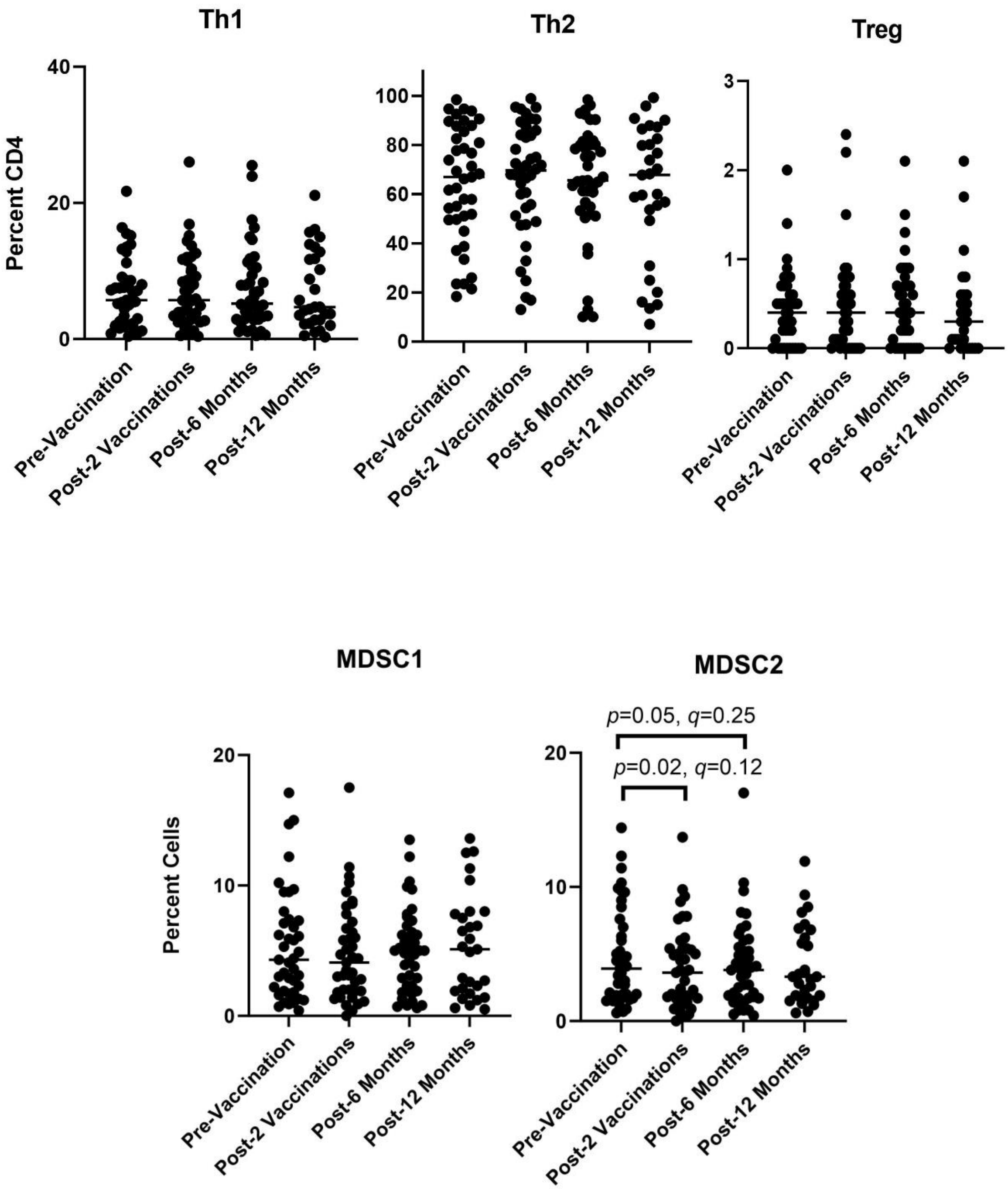

Th1
Th2
Treg
Percent CD4
40
20
0
100
80
60
40
20
0
3
2
1
0
Pre-Vaccination
Post-2 Vaccinations
Post-6 Months
Post-12 Months
MDSC1
MDSC2
Percent Cells
20
10
0
p=0.05, q=0.25
p=0.02, q=0.12

,

**Figure S5. Differential abundance of taxa.**

ALDEx2 (ANOVA-like differential expression, version 2) was utilized to examine differential abundance of individual taxa. Panel A shows effect plots comparing taxa between histological responders and non-responders (stringent) at baseline for patients who completed the 6-month visit in both, PepCan, or *Candida* groups. No individual taxa were associated with histological response. Panel B shows differentially abundant taxa in both groups comparing microbiota at 6 month or 12 month to the baseline. Panel C shows differentially abundant taxa in the PepCan group comparing microbiota at 6 month or 12 month to the baseline. Panel D shows differentially abundant taxa in the *Candida* group comparing microbiota at 6 month or 12 month to the baseline. Panel E shows differentially abundant taxa in histological responders (stringent) comparing microbiota at 6 month or 12 month to the baseline. Panel F shows differentially abundant taxa in the histological non-responders comparing microbiota at 6 month or 12 month to the baseline. The red dots indicate significantly differentially abundant taxa using Benjamini-Hochberg corrected *p*-value of Wilcoxon test.

**A**

**Both** **PepCan** ***Candida***

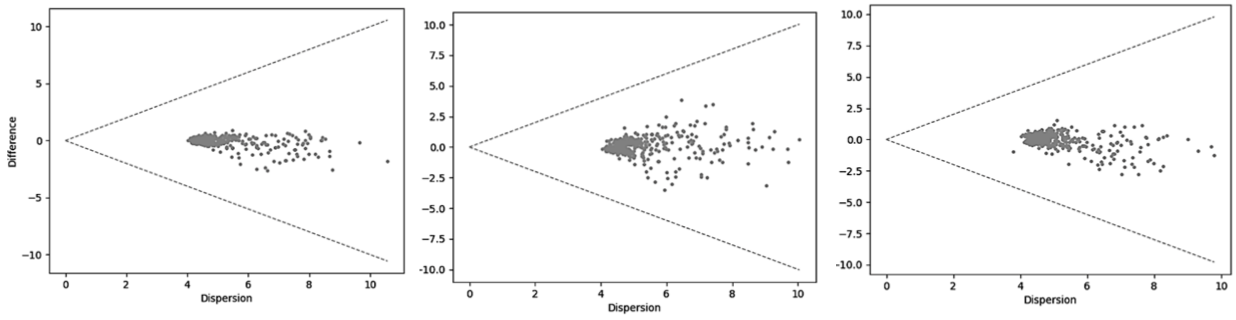

## B

##### Both

###### 6-Month vs Baseline

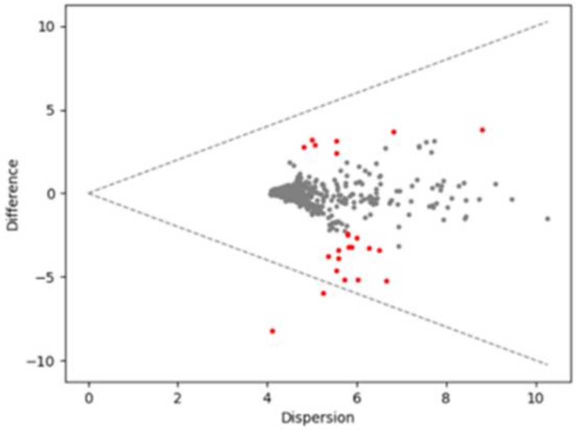

###### 12-Month vs Baseline

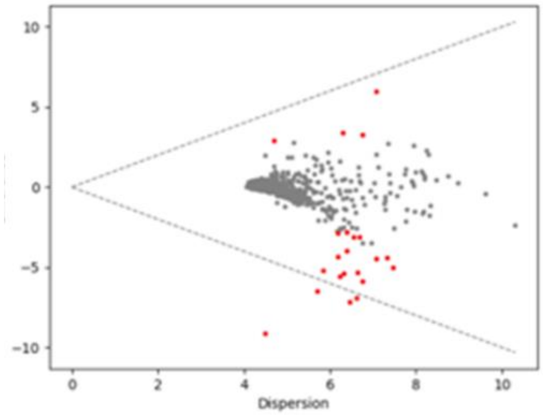

## C

##### PepCan

###### 6-Month vs Baseline

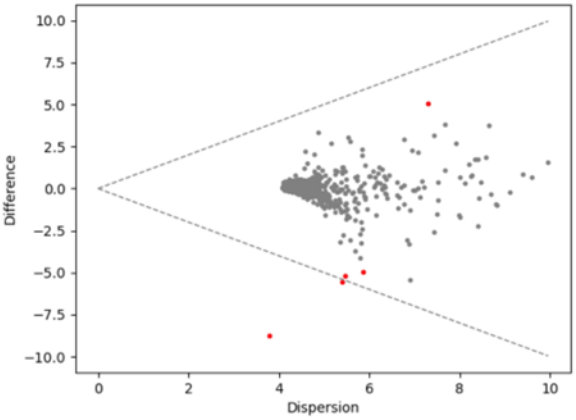

###### 12-Month vs Baseline

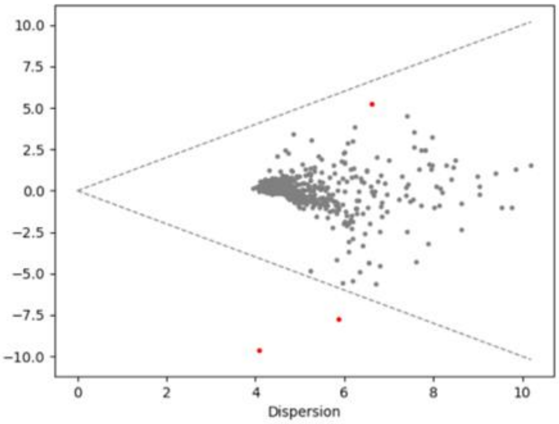

**D**

***Candida***

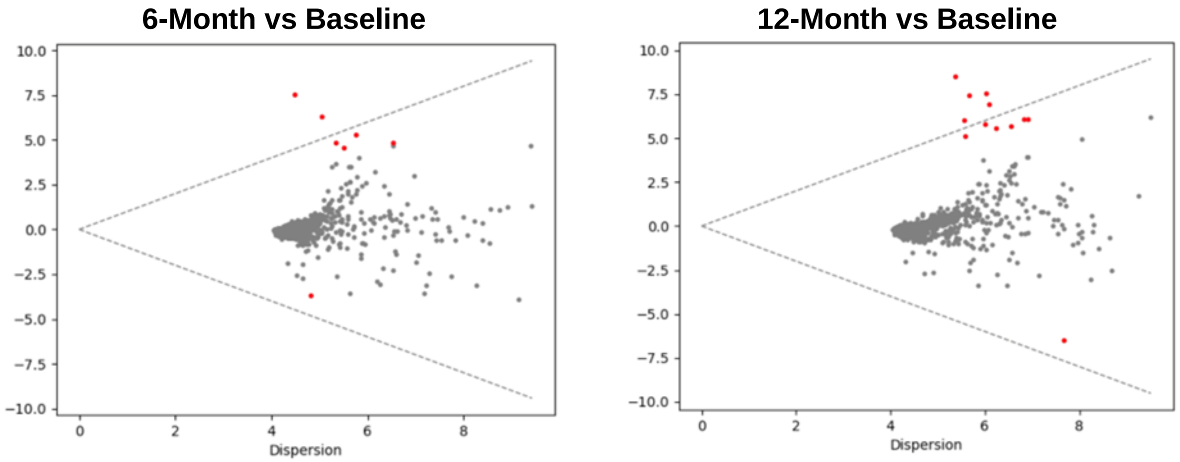

**E**

**Responders**

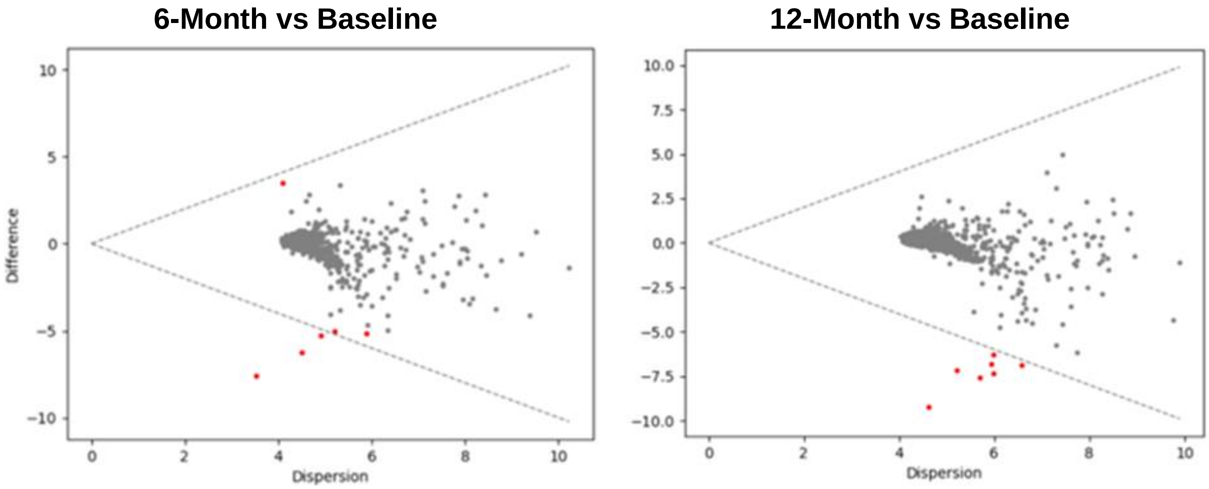

**F**

<table>
<tr><th colspan="2">Non-Responders</th></tr>
<tr><th>6-Month vs Baseline</th><th>12-Month vs Baseline</th></tr>
<tr><td>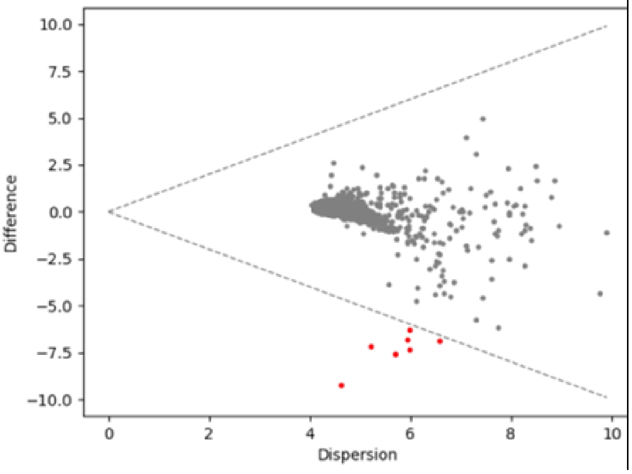
</td><td>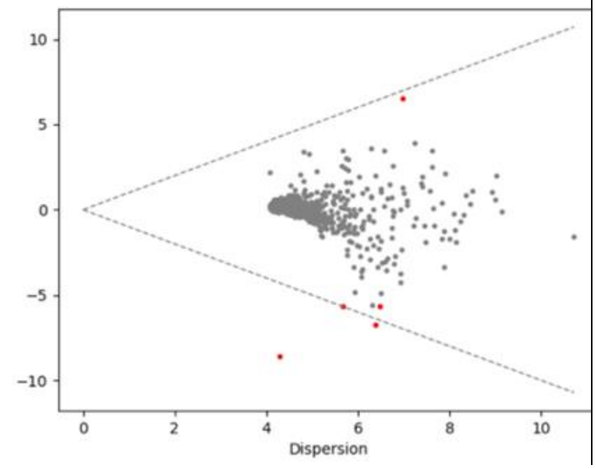
</td></tr>
</table>

**Figure S6. Observed HLA frequencies for patients in comparison to the estimates for the United States population based on racial distributions.**

Panel A shows the A alleles, panel B shows the B alleles, panel C show the C alleles, and panel D shows the DR/DQ alleles. The filled black circles show the frequency patients who received at least one vaccination, the line shows its 95% confidence interval (Pearson-Klopper method), and the red “x” shows estimated frequency in the United States population.

**A**

| HLA Type | % (n) | 95% CI | US Estimate |
|---|---|---|---|
| A02 | 31.82% (49) | 24.55% to 39.80% | 28.65% |
| A01 | 14.29% (22) | 9.17% to 20.83% | 11.96% |
| A24 | 11.04% (17) | 6.56% to 17.09% | 10.02% |
| A03 | 7.79% (12) | 4.09% to 13.22% | 11.46% |
| A68 | 5.19% (8) | 2.27% to 9.98% | 6.24% |
| A29 | 5.19% (8) | 2.27% to 9.98% | 3.59% |
| A26 | 3.90% (6) | 1.44% to 8.29% | 2.55% |
| A23 | 3.90% (6) | 1.44% to 8.29% | 3.16% |
| A32 | 3.25% (5) | 1.06% to 7.41% | 2.77% |
| A31 | 3.25% (5) | 1.06% to 7.41% | 3.12% |
| A25 | 2.60% (4) | 0.71% to 6.52% | 1.44% |
| A66 | 1.95% (3) | 0.40% to 5.59% | 0.65% |
| A33 | 1.95% (3) | 0.40% to 5.59% | 2.48% |
| A11 | 1.95% (3) | 0.40% to 5.59% | 5.24% |
| A30 | 1.30% (2) | 0.16% to 4.61% | 4.00% |
| A74 | 0.65% (1) | 0.02% to 3.56% | 0.72% |

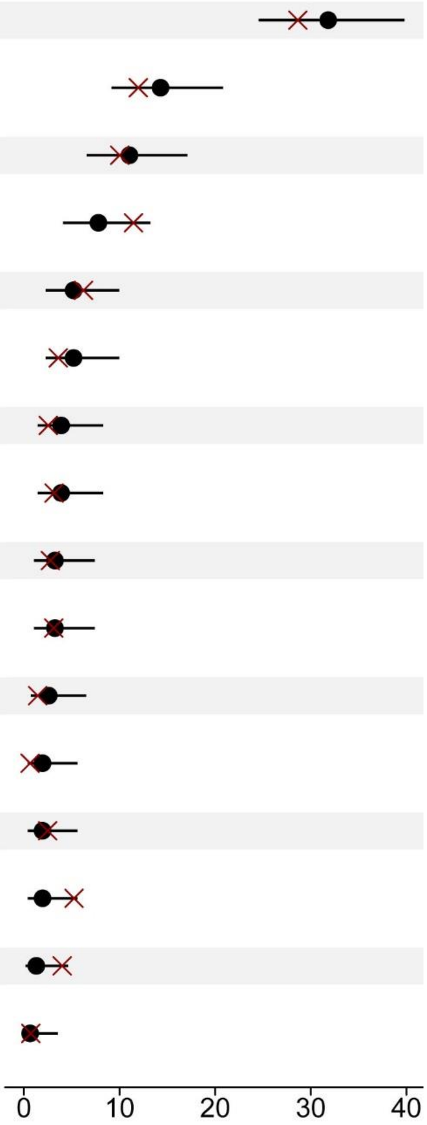

**B**

| HLA Type | % (n) | 95% CI | US Estimate |
|---|---|---|---|
| B44 | 15.38% (24) | 10.11% to 22.02% | 12.42% |
| B07 | 12.82% (20) | 8.01% to 19.10% | 10.88% |
| B35 | 10.90% (17) | 6.48% to 16.87% | 10.55% |
| B15 | 8.33% (13) | 4.51% to 13.83% | 0.10% |
| B08 | 8.33% (13) | 4.51% to 13.83% | 8.31% |
| B40 | 6.41% (10) | 3.12% to 11.47% | 0.24% |
| B51 | 5.13% (8) | 2.24% to 9.85% | 5.42% |
| B49 | 5.13% (8) | 2.24% to 9.85% | 1.77% |
| B39 | 5.13% (8) | 2.24% to 9.85% | 3.52% |
| B14 | 4.49% (7) | 1.82% to 9.03% | 1.00% |
| B57 | 3.21% (5) | 1.05% to 7.32% | 3.21% |
| B45 | 3.21% (5) | 1.05% to 7.32% | 1.44% |
| B18 | 2.56% (4) | 0.70% to 6.43% | 4.47% |
| B27 | 1.92% (3) | 0.40% to 5.52% | 3.50% |
| B50 | 1.28% (2) | 0.16% to 4.55% | 1.26% |
| B37 | 1.28% (2) | 0.16% to 4.55% | 1.17% |
| B13 | 1.28% (2) | 0.16% to 4.55% | 1.74% |
| B81 | 0.64% (1) | 0.02% to 3.52% | 0.18% |
| B58 | 0.64% (1) | 0.02% to 3.52% | 1.78% |
| B55 | 0.64% (1) | 0.02% to 3.52% | 1.12% |
| B41 | 0.64% (1) | 0.02% to 3.52% | 1.08% |
| B38 | 0.64% (1) | 0.02% to 3.52% | 1.54% |

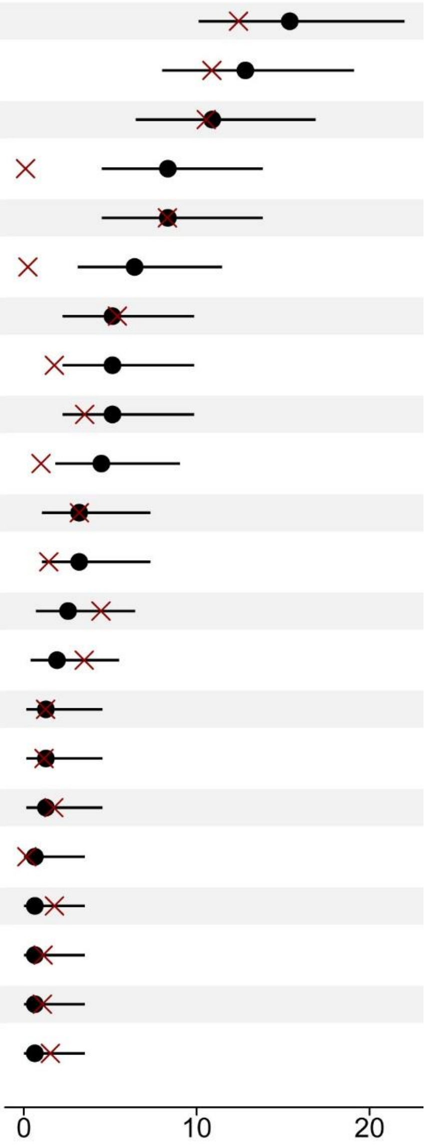

**C**

| HLA Type | % (n) | 95% CI | US Estimate |
|---|---|---|---|
| C07 | 34.21% (52) | 26.72% to 42.33% | 28.57% |
| C03 | 12.50% (19) | 7.70% to 18.83% | 5.48% |
| C06 | 10.53% (16) | 6.14% to 16.53% | 9.41% |
| C04 | 8.55% (13) | 4.63% to 14.18% | 14.08% |
| C05 | 7.24% (11) | 3.67% to 12.58% | 8.20% |
| C16 | 6.58% (10) | 3.20% to 11.77% | 3.17% |
| C08 | 4.61% (7) | 1.87% to 9.26% | 4.54% |
| C01 | 4.61% (7) | 1.87% to 9.26% | 4.19% |
| C15 | 3.29% (5) | 1.08% to 7.51% | 2.64% |
| C12 | 2.63% (4) | 0.72% to 6.60% | 3.50% |
| C02 | 2.63% (4) | 0.72% to 6.60% | 5.34% |
| C17 | 1.32% (2) | 0.16% to 4.67% | 1.48% |
| C14 | 1.32% (2) | 0.16% to 4.67% | 0.69% |

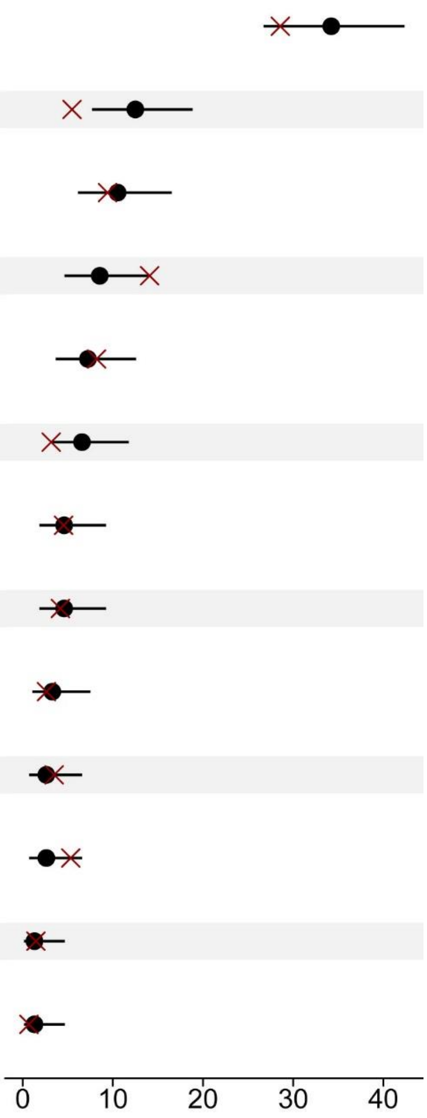

**D**

| HLA Type | % (n) | 95% CI | US Estimate |
|---|---|---|---|
| DR04 | 20.62% (33) | 14.64% to 27.73% | 17.76% |
| DR07 | 15.62% (25) | 10.37% to 22.20% | 11.28% |
| DR15 | 15.00% (24) | 9.85% to 21.49% | 12.77% |
| DR03 | 10.00% (16) | 5.82% to 15.73% | 0.59% |
| DR13 | 8.75% (14) | 4.87% to 14.25% | 11.50% |
| DR11 | 8.12% (13) | 4.40% to 13.49% | 9.59% |
| DR01 | 7.50% (12) | 3.94% to 12.73% | 8.80% |
| DR14 | 5.00% (8) | 2.18% to 9.61% | 3.93% |
| DR08 | 5.00% (8) | 2.18% to 9.61% | 5.49% |
| DR09 | 2.50% (4) | 0.69% to 6.28% | 1.39% |
| DR12 | 1.25% (2) | 0.15% to 4.44% | 1.96% |
| DR16 | 0.62% (1) | 0.02% to 3.43% | 2.23% |
| DQ03 | 38.12% (61) | 30.57% to 46.13% | 3.88% |
| DQ02 | 22.50% (36) | 16.28% to 29.76% | 19.88% |
| DQ06 | 21.25% (34) | 15.19% to 28.41% | 19.48% |
| DQ05 | 10.00% (16) | 5.82% to 15.73% | 13.70% |
| DQ04 | 8.12% (13) | 4.40% to 13.49% | 5.81% |

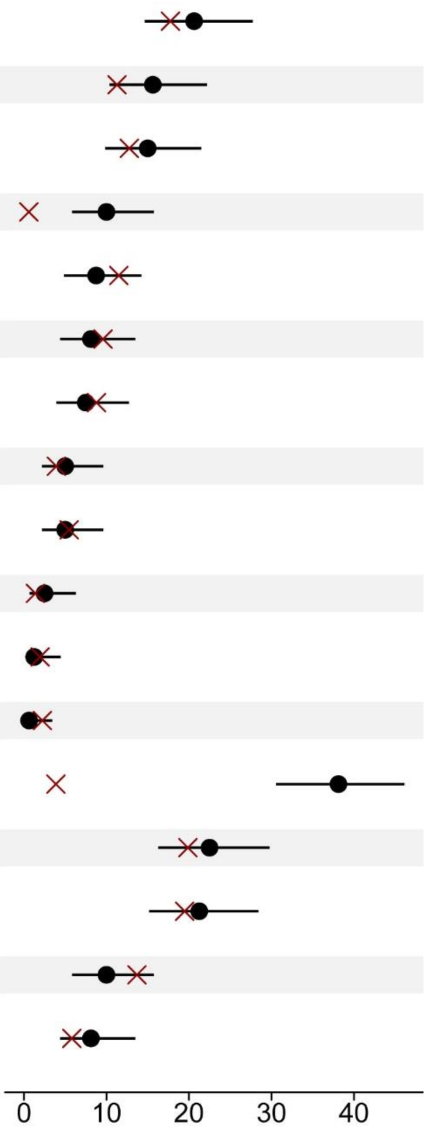
